# Frequency-dependent cognitive effects of Deep Brain Stimulation in Parkinson’s Disease: A Systematic Review and Meta-Analysis

**DOI:** 10.64898/2026.06.17.26355717

**Authors:** Bruna Meira, Paulo Bastos, Vítor Mendes Ferreira, João Albuquerque, Marta Magriço, Raquel Lemos, Raquel Barbosa, Miguel Coelho, Marcelo Mendonça

## Abstract

**Background:** Subthalamic nucleus deep brain stimulation (STN-DBS) improves levodopa-induced motor complications and cardinal motor symptoms of Parkinson’s disease (PD), but stimulation frequency may differentially shape outcomes. This is evident for axial and gait symptoms, which may respond differently to lower-frequency stimulation. Whether frequency-dependent effects extend to cognition remains unclear.

**Objective:** To investigate the cognitive effects of DBS at distinct frequencies in PD.

**Methods:** We conducted a systematic review and meta-analysis (PROSPERO - CRD42024618253). PubMed, Web of Science, and EMBASE were searched for studies assessing cognitive outcomes under different stimulation frequencies. Eight cognitive domains were defined: verbal fluency, cognitive flexibility, executive control, working memory, attention, processing speed, episodic memory, and time processing. Multilevel random-effects meta-analyses were performed, with effect sizes expressed as Hedges’ g.

**Results:** Forty-three studies met the inclusion criteria, the majority (n = 31) involving STN-DBS. Twenty-one STN-DBS studies, including 355 patients, were included in the meta-analysis. Compared with HFS (≥ 130 Hz), lower frequencies (4-80 Hz) were associated with better verbal fluency (g = 0.27) and cognitive flexibility (g = 0.38), with consistent effects across sensitivity and leave-one-out analyses. Accuracy-based executive control measures also favored lower-frequency stimulation. OFF-stimulation comparisons showed a concordant pattern. Evidence for other targets (PPN and NBM) was limited.

**Conclusions:** Lower-frequency STN-DBS was associated with modest benefits in specific cognitive domains compared with HFS. These findings highlight the need for future research to determine how frequency interacts with stimulation location and symptom-specific networks to shape cognitive and cognitive-motor outcomes in PD.

**Highlights:**

- Systematic Review of 43 studies addressing he cognitive effects of DBS at distinct frequencies.
- Low frequency STN-DBS improves verbal fluency and cognitive flexibility vs high-frequency stimulation.
- Low frequency STN-DBS appears to benefit verbal fluency and executive control in comparison to OFF stimulation.
- Effects are robust across multiple sensitivity analyses and are independent from medication state.

## Introduction

Deep brain stimulation (DBS) of the subthalamic nucleus (STN) is a well-established and highly effective treatment for motor symptoms in Parkinson’s disease (PD).^1–3^ High-frequency stimulation (HFS≈130 Hz) remains the clinical standard for improving tremor, rigidity, and bradykinesia; however, gait and other axial symptoms often respond less consistently^4–7^ and may even worsen under HFS settings in some patients.^8–10^ These dissociations suggest that stimulation frequency does not exert uniform effects across clinical domains. HFS improves appendicular motor function while potentially differentially modulating networks involved in gait and axial control.^11^

This circuit-level dissociation is also relevant to cognition.^12,13^ Studies on HFS STN-DBS consistently report subtle but reproducible cognitive changes, most notably reduced verbal fluency.^14–16^ Although often clinically mild, such changes highlight that STN stimulation can influence cognitive circuits and suggest that stimulation may differentially shape appendicular, axial or cognitive outcomes.^13,17–20^

These observations have driven interest in low-frequency stimulation (LFS). Several studies suggest that 60-80 Hz stimulation may be more effective than HFS for improving freezing of gait (FoG), gait speed, stride length, postural control, and other axial functions.^9,21–31^ Yet, the cognitive consequences of LFS remain insufficiently characterized.^32–36^ This gap is clinically relevant as cognitive processes contribute to real-world motor behavior, including gait adaptation, navigation, and response to environmental demands. Although evidence linking stimulation frequency to cognitive outcomes has begun to emerge^37,38^, it has not been systematically synthesized across cognitive domains or stimulation targets. Existing studies also rely predominantly on chronic before-after comparisons. The acute cognitive effects of within-session frequency modulation (the paradigm best positioned to isolate frequency-specific effects from disease progression, medication state, and inter-individual variability) remain entirely unreviewed.

We therefore conducted a systematic review and meta-analysis to compare the effects of DBS frequency on cognition across stimulation targets, using neuropsychological measures.

## Methods

### Literature Search and Study Selection

The systematic review and meta-analysis was conducted in accordance with the Preferred Reporting Items for Systematic Reviews and Meta-Analyses (PRISMA) guidelines^39^ and was registered in PROSPERO CRD42024618253. Three electronic bibliographic databases (PubMed, Web of Science and EMBASE) were searched for studies investigating the effects of low-frequency stimulation on cognitive domains in PD patients with DBS. The following terms were used in database search: (1) “Parkinson’s disease” or “PD” AND (2) “brain stimulation” or “DBS” AND (3) “cogniti*”, “attention”, “memory”, “executive”, “frontal”, “language”, “verbal fluency”, “visuospatial” AND (4) “frequency”, “LFS”, “low-frequency” OR “high-frequency”. The search included all studies indexed in the selected databases, regardless of publication date, and was restricted to publications in English, French, Portuguese, or Spanish. Duplicates were initially removed using Rayyan software from the combined records retrieved from the three databases. Subsequently, titles, abstracts, and full-text articles were independently screened by three reviewers (B.M., J.A., V.M.F.) and assessed for eligibility according to the following inclusion criteria: randomized controlled trials (RCTs), non-randomized controlled trials (N-RCTs), case series, and case reports evaluating the impact of DBS with different frequency parameters (or OFF state) on cognitive functioning in adults with PD. Literature reviews and meta-analyses were screened to identify additional eligible studies that may had been missed by the original search strategy.

The initial database search was conducted in November 2024. To ensure inclusion of the most recent publications relevant to the scope of the review, the search was updated between November 2024 and December 2025 (Figure 1).

**Figure 1:**
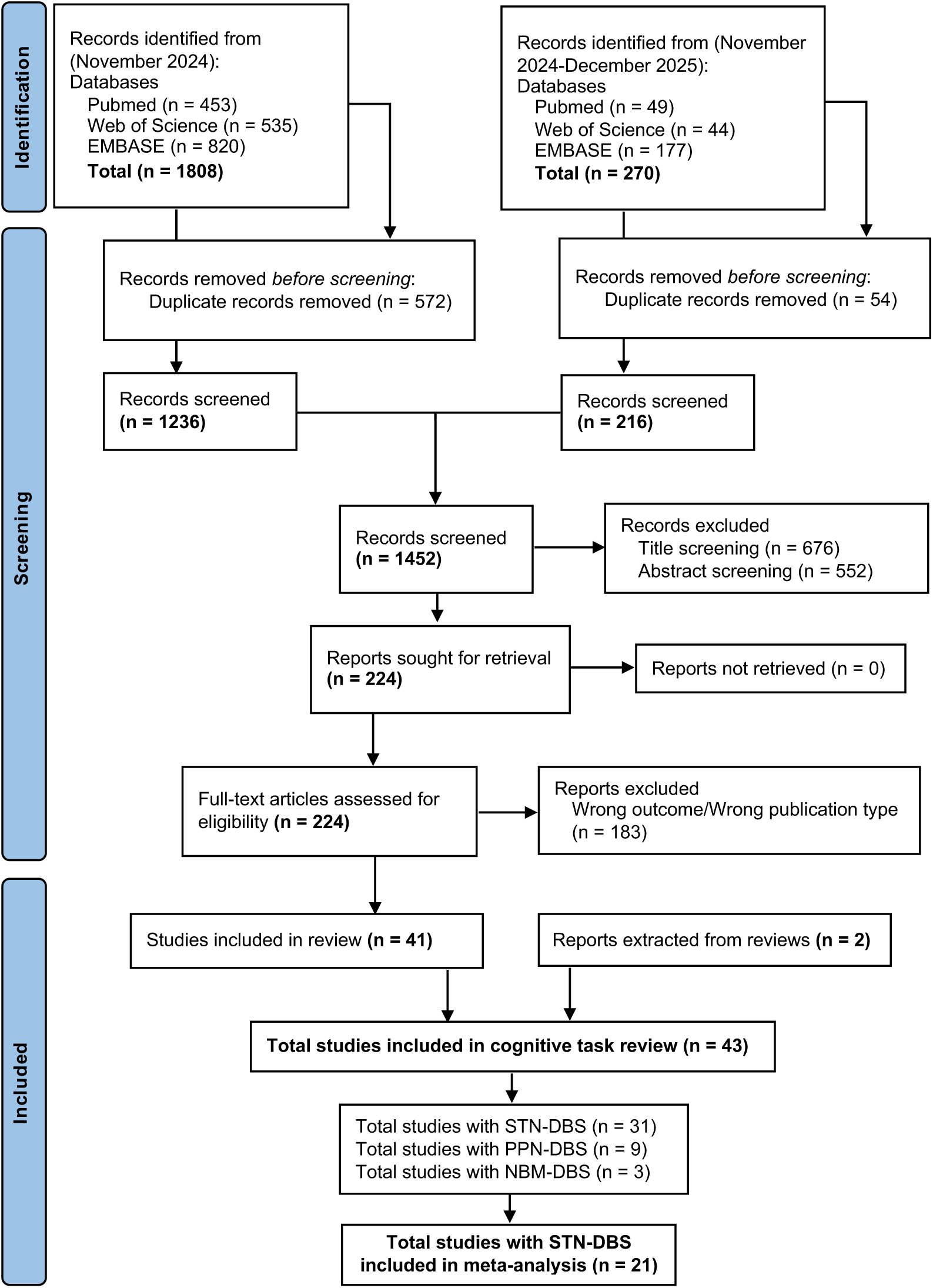
Flow diagram of study selection and inclusion

### Data Extraction

For each paper, information extracted included (1) patient characteristics including sample size, gender, age, education, disease duration, time from DBS, levodopa equivalent daily dosage (LEDD), Unified Parkinson’s Disease Rating Scale (UPDRS/MDS-UPDRS) Part III (2) intervention characteristics, including experimental design, stimulation target, unilateral or bilateral DBS implant, DBS stimulation parameters, medication status, time between assessments and (3) cognitive measures, including tasks, outcomes and scores for each manipulation as mean and standardized mean difference (SMD).

### Study Quality Assessment

The methodological quality of the included studies was assessed using the Cochrane Risk of Bias 2 (RoB-2)^40^ tool for randomized and crossover trials and the ROBINS-I^41^ tool for non-randomized studies.

### Cognitive outcomes definition

Extracted cognitive outcomes were classified according to the process each measure indexes, rather than the task from which it was derived. This distinction matters because different measures from the same paradigm (i.e. accuracy, reaction time, error rate, interference cost) can reflect distinct cognitive mechanisms and should not be collapsed into a single outcome. Outcomes were mapped onto a previously published eight-domain framework^42,43^ comprising verbal fluency, working memory, cognitive flexibility, executive control, attention, processing speed, episodic memory, and time processing.

For each outcome we assigned a primary domain, and when relevant, additional secondary and tertiary domains were recorded to capture less central but meaningful cognitive demands (Supplementary Table 1).

Subdomains were identified for Verbal Fluency: Letter, Semantic, Action and Switching; and Executive Control was further stratified into two subdomains: accuracy-based measures (accuracy or error rate) and time-based measures (reaction time).

### Deep Brain stimulation features

Stimulation targets and frequency were extracted. For STN-DBS, stimulation frequency was classified in very low (VLFS, 4-10 Hz), low (LFS, 60-80 Hz), high (HFS, ≥130 Hz), and OFF stimulation. Frequency grouping was not performed for other targets.

### Meta-analysis

Meta-analysis was only performed for STN-DBS as data for other stimulation targets were too limited for quantitative synthesis. Means and standard deviations (SDs) were extracted directly or derived from reported statistics (e.g., standard error of the mean, 95% confidence intervals, or effect sizes) to ensure consistency across studies. Effect sizes were calculated as standardized mean differences (Hedges’ g) for each cognitive outcome.

To account for the dependency of multiple assessments within the same study and participants, we employed a multilevel random-effects model incorporating both study-level and outcome-level variance components, assuming a within-study correlation of r=0.5. Meta-analyses were performed with the metafor package in R. Sensitivity analyses included recalculating effect sizes across a range of assumed correlations (r=0.0-0.9) and conducting leave-one-study-out analyses.

The principal outcome (key comparison) was the contrast between HFS vs. combined LFS+VLFS for primary cognitive domains. Additional robustness analyses were performed at the subdomain level. Expanded more permissive analyses used secondary and tertiary domains (expanded domain classification analysis). Secondary outcome included HFS vs. LFS and HFS vs. VLFS separately. Exploratory outcomes included OFF vs. HFS, and OFF vs. LFS+VLFS.

Because medication state may influence cognitive performance, we performed an additional moderator analysis to test whether frequency-related cognitive effects differed between MedON and MedOFF conditions. We used robust variance estimation (RVE) to account for multiple dependent effect sizes contributed by the same studies and to obtain corrected standard errors.

Significance level was set at 0.05. To account for multiple testing across the different cognitive domains, Benjamini-Hochberg false discovery rate correction was applied in the primary analysis (HFS vs. LFS+VLFS). Other outcomes are considered secondary/exploratory and are presented without correction for multiple comparisons.

Further details are provided in Supplementary Methods.

## Results

After retrieving 1452 studies from the initial search, 43 studies met the inclusion criteria: 31 studies with STN^37,44–73^, 9 with PPN^74–82^ and 3 NMB^83–85^ (Figure 1).

### Subthalamic nucleus (STN)

Among STN-DBS (Table 1) studies, twenty-one articles were included in the meta-analysis. These 21 STN-DBS studies included a total of 355 patients (mean sample size 16.9±4.7 patients), with a mean age of 62.2±2.9 years and average PD duration of 12.3±3.6 years. The average time between surgical intervention and evaluation was 23.9±20.9 months (Figure 2.A). Risk-of-bias was low or raised only some concerns in the randomized and crossover trials, with the most frequent uncertainties related to carryover effects (inherent to crossover DBS designs). A single non-randomized study was rated as high confounding risk (details on Supplementary Figure 1). Ten studies (129 patients) were excluded from the meta-analysis due to incomparable or insufficient data and were included in a qualitative synthesis only.

**Figure 2:**
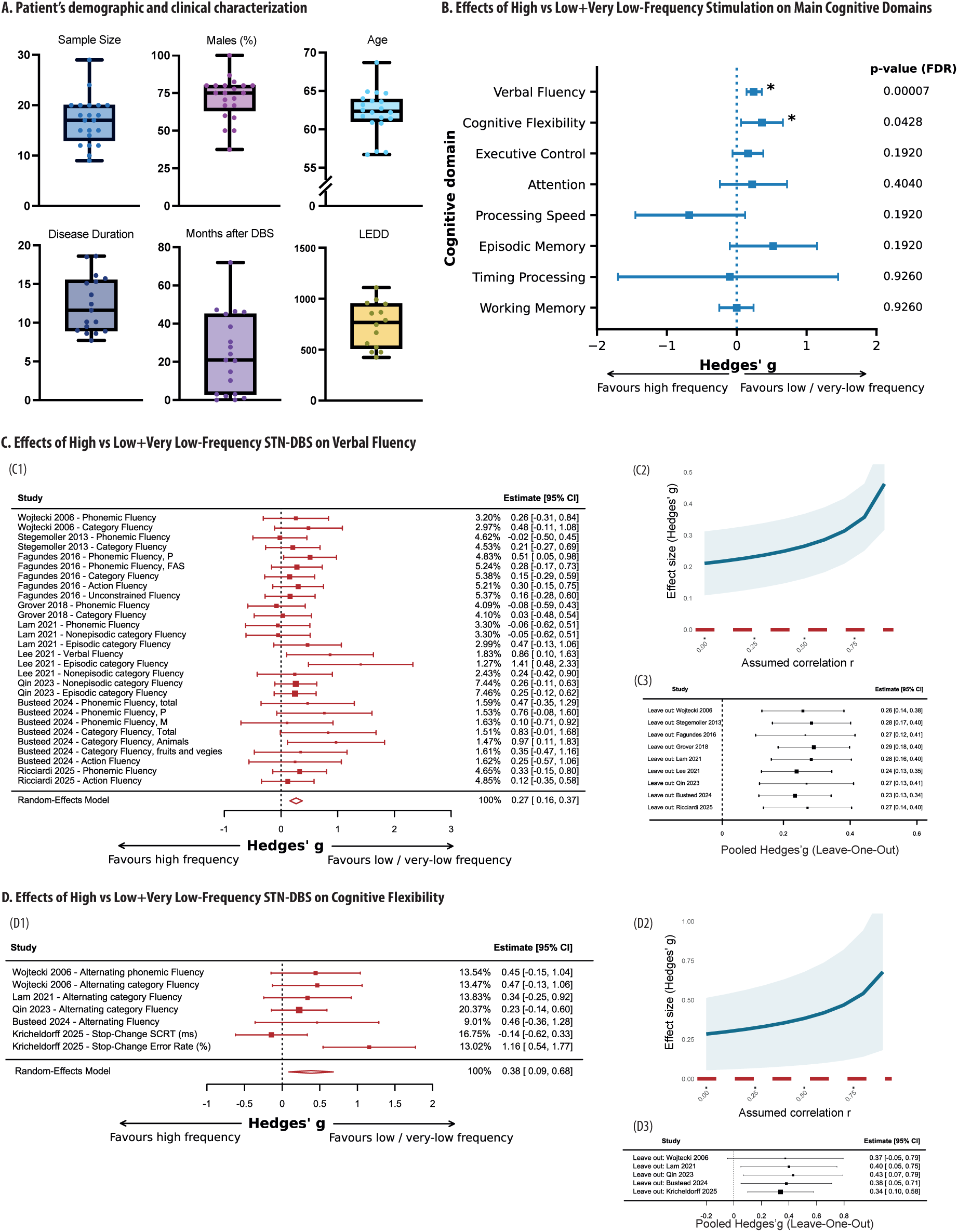
Effects of High-Frequency vs Low+Very Low-Frequency STN-DBS on cognition. **A. Patient’s demographic and clinical characterization**. The 21 STN-DBS studies included in the meta-analysis comprised 355 patients, 251 males (mean age 62.2±2.9 years, mean PD duration 12.3±3.6 years, mean time after surgery 23.9±20.9 months). **B. Effects of High vs. Low+Very Low-Frequency on Main Cognitive Domains.** Comparison of High-frequency stimulation (HFS) vs. Low- and Very low-frequency stimulation (LFS+VLFS) on cognitive outcomes in STN-DBS. LFS+VLFS was associated with modest but statistically significant improvements in verbal fluency and cognitive flexibility compared with HFS. No significant differences were observed across the remaining cognitive domains. p-values corrected using the Benjamini–Hochberg false discovery rate method to account for multiple testing across the different cognitive domains. *P_(FDR)_<0.05 **C. Effects of High vs. Low+Very Low-Frequency STN-DBS on Verbal Fluency.** (C1) Multilevel meta-analysis adjusting for study-level dependence. Forest plot showing Hedges’ g effect sizes for Verbal Fluency comparing HFS vs. LFS+VLFS STN-DBS across studies, estimated using a multilevel random-effects meta-analysis that accounts for dependency among multiple Verbal Fluency outcomes within the same study (effects nested within study). Positive values indicate better Verbal Fluency performance under LFS+VLFS relative to HFS. (C2) Pooled Hedges’ g estimate and 95% confidence interval across a range of assumed within-subject correlations (r=0.0-0.9). (C3) Leave-one-study-out sensitivity analysis of the HFS vs. LFS+VLFS comparison. Each point represents the pooled Hedges’ g estimated from the multilevel random-effects meta-analysis after removing one study at a time. Horizontal error bars indicate 95%CI. The vertical dashed line denotes the pooled effect estimate from the full model including all studies. Positive values indicate better Verbal Fluency performance under LFS+VLFS relative to HFS. **D. Effects of High vs. Low+Very Low-Frequency STN-DBS on Cognitive Flexibility.** (D1) Forest plot showing Hedges’ g effect sizes for Cognitive Flexibility comparing HFS vs. LFS+VLFS STN-DBS across studies. (D2) Pooled Hedges’ g estimate and 95% confidence interval across a range of assumed within-subject correlations (r=0.0-0.9). (D3) Leave-one-study-out sensitivity analysis of the HFS vs. LFS+VLFS comparison.

**Table 1:**
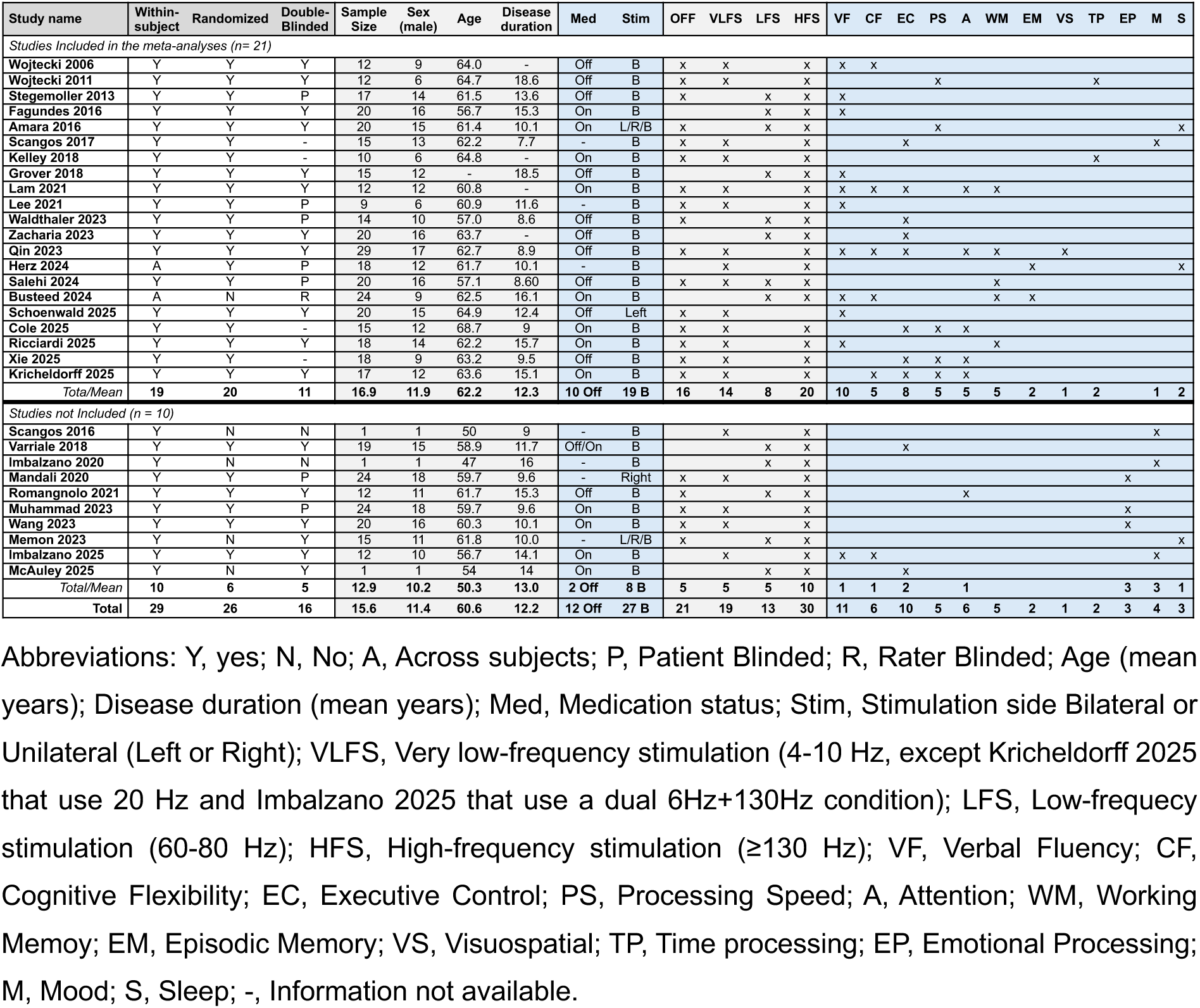
Characteristics of STN-DBS included studies. Abbreviations: Y, yes; N, No; A, Across subjects; P, Patient Blinded; R, Rater Blinded; Age (mean years); Disease duration (mean years); Med, Medication status; Stim, Stimulation side Bilateral or Unilateral (Left or Right); VLFS, Very low-frequency stimulation (4-10 Hz, except Kricheldorff 2025 that use 20 Hz and Imbalzano 2025 that use a dual 6Hz+130Hz condition); LFS, Low-frequecy stimulation (60-80 Hz); HFS, High-frequency stimulation (≥130 Hz); VF, Verbal Fluency; CF, Cognitive Flexibility; EC, Executive Control; PS, Processing Speed; A, Attention; WM, Working Memoy; EM, Episodic Memory; VS, Visuospatial; TP, Time processing; EP, Emotional Processing; M, Mood; S, Sleep; -, Information not available.

### High Frequency vs. Low+Very Low Frequency STN-DBS

#### Main Cognitive Domains

Figure 2.B summarizes the meta-analysis comparing low and very low-frequency (LFS+VLFS) with high-frequency stimulation (HFS) on main cognitive domains. Domain-specific analyses (Figure 2.C1, 3.D1) indicated significantly better performance under LFS+VLFS than HFS for Verbal Fluency (k=28, 9 studies, g=0.27, SE=0.06, 95% CI [0.16, 0.37], p_(FDR)_<0.001) and Cognitive Flexibility (k=7, 5 studies, g=0.38, SE=0.15, 95% CI [0.09, 0.68], p_(FDR)_=0.043).

Sensitivity analyses supported the robustness of these findings. Across the full range of assumed within-study correlations (r=0.0-0.9), pooled effect sizes for these two domains remained positive and statistically significant (Figure 2.C2, D2). Likewise, leave-one-study-out analysis (Figure 2.C3, D3), consistently showed a positive effect favoring LFS+VLFS over HFS on these domains, with omission of individual studies producing no reversal in direction and no extreme shift in magnitude. This suggests that the observed effects were not driven by any single study.

No significant effects were observed in the remaining 6 domains (Figure 2.B, Supplementary Figure 2): Processing Speed (p_(FDR)_=0.192), Attention (p_(FDR)_=0.404), Time Processing (p_(FDR)_=0.926), Executive Control (p_(FDR)_=0.192), Working Memory (p_(FDR)_=0.926) and Episodic Memory (p_(FDR)_=0.192).

#### Robustness analysis

##### Verbal Fluency Subdomains

When verbal fluency (VF) was further subdivided (Supplementary Figure 3), the benefits of LFS+VLFS over HFS remained consistent. Significant effects of comparable effects sizes were observed for Semantic VF (k=13, g=0.30, p=0.001), Letter VF (k=10, g=0.22, p=0.012) and Switching VF (k=5, g=0.34, p=0.005). Action VF showed a non-significant effect in the same direction (k=3, g=0.22, p=0.155).

##### Executive Control Subdomains

Post-hoc stratification of executive control measures into accuracy- and time-based outcomes (Supplementary Figure 4) suggested that the overall null effect in this domain masked a potential differential pattern. In the context of STN-DBS, changes in stimulation frequency are known to affect motor performance and may therefore directly influence response-time measures independently of executive processing. LFS+VLFS was associated with significantly better performance on accuracy-based executive control measures (k=12, g=0.35, p=0.032), whereas a non-significant negative signal was observed for time-based measures (k=8, g=-0.10, p=0.666) depending on motor performance.

#### Expanded domain classification: Primary+Secondary+Tertiary Domains

A broader, more inclusive meta-analysis was conducted by including any cognitive task associated with a given cognitive domain, regardless of whether that domain has been classified as primary, secondary, or tertiary. A better performance of LFS+VLFS when compared with HFS was still observed for Verbal Fluency (k=33, 9 studies, g=0.28, SE=0.05, 95% CI [0.18, 0.38], p<3×10⁻⁸) and Cognitive Flexibility (k=35, 10 studies, g=0.28, SE=0.05, 95% CI [0.19, 0.37], p<1×10⁻⁹). Results remained non-significant across all other domains (Supplementary Figure 5).

#### Study Design and Medication Conditions Variables

We examined whether the pooled effects observed for verbal fluency and cognitive flexibility were disproportionately influenced by study design or medication status. For verbal fluency, most studies used randomized within-subject designs (n=8), with only one non-randomized between-subject study^66^. Six studies were double-blind, while three used single-blind protocols^46,56,66^. All applied bilateral STN stimulation. Medication status varied between OFF (four studies^44,46,52,62^) and ON (four studies^49,55,66,71^) conditions, with one study not reporting it^56^.

For cognitive flexibility, all studies except one^66^ had a randomized within-subject, double-blind design. All assessed bilateral STN stimulation. Medication status varied between OFF (two studies^44,62^) and ON (three studies^55,66,73^) and not reported (one studied^56^).

Using Robust Variance Estimation (RVE) with medication status as a moderator, we examined its influence on verbal fluency and cognitive flexibility outcomes. No significant effect of medication status was observed (p=0.167 and p=0.868, respectively), with effect sizes remaining positive, favoring LFS+VLFS, regardless of ON vs. OFF medication states.

### High Frequency vs. Low Frequency STN-DBS

#### Main Cognitive Domains

Domain-specific analyses (Figure 3.A) revealed a significant improvement in Verbal Fluency (Figure 3.C1) under LFS compared to HFS (k=16, 4 studies, g=0.23, 95% CI [0.02, 0.44], p=0.03). The pooled effect size remained consistently positive and statistically significant across correlation sensitivity analyses and leave-one-study-out iterations (Figure 3.C2-C3). Executive Control (k=6, 2 studies) and Working Memory (k=2, 2 studies) failed to reveal any significant difference (Supplementary Figure 6). Other domains had less than 2 studies: cognitive flexibility, processing speed and episodic memory results are based on single studies (Figure 3.A).

**Figure 3:**
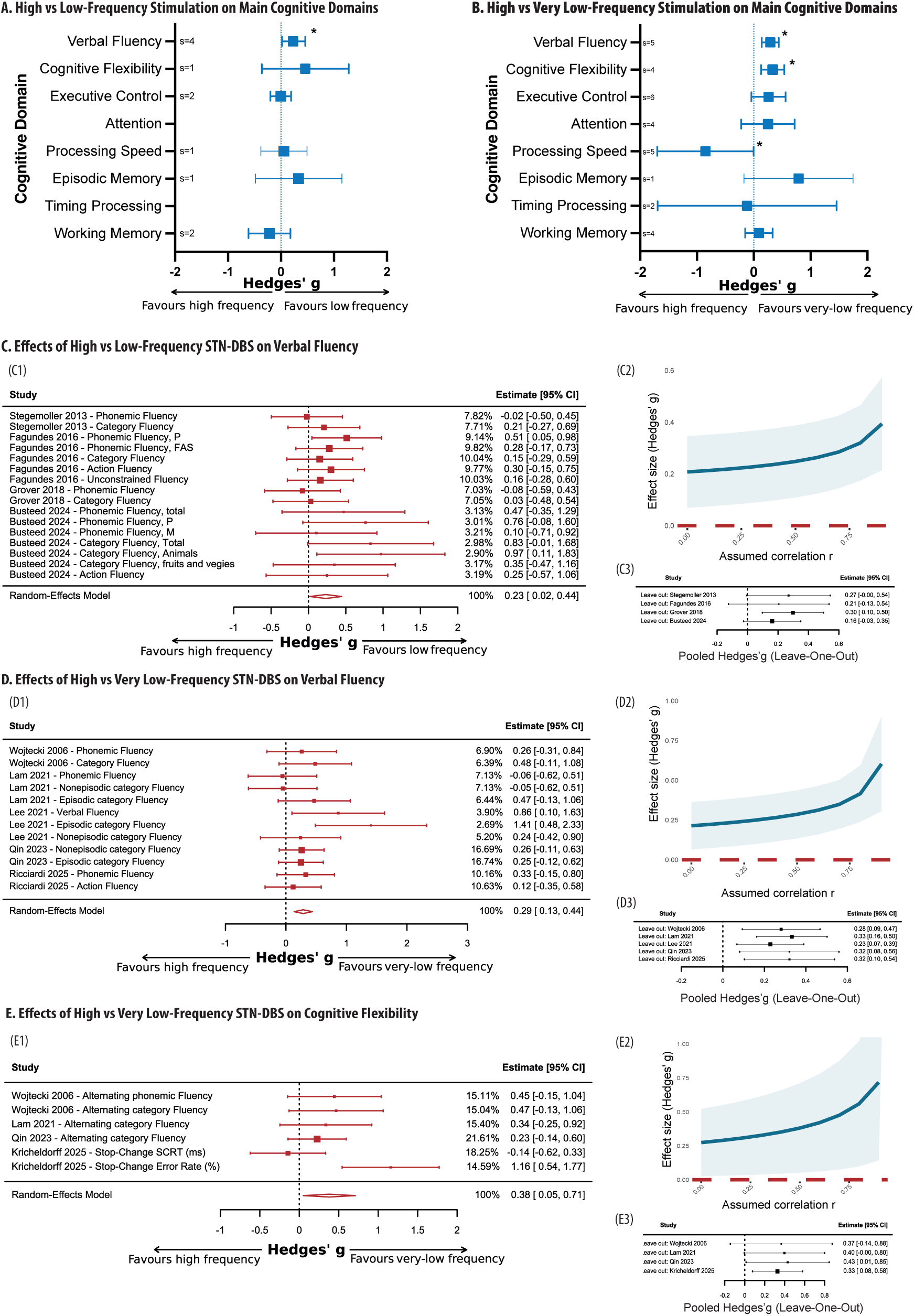
Effects of High-Frequency vs Low or Very Low-Frequency STN-DBS on cognition. **A. Comparison of High-Frequency vs. Low-Frequency on cognitive outcomes.** LFS was associated with modest but statistically significant improvements in verbal fluency compared with HFS. **B. Comparison of High vs. Very Low-Frequency on cognitive outcomes.** Findings were consistent across comparisons of pooled LFS+VLFS vs. HFS, as well as separate analyses of HFS vs. LFS and HFS vs. VLFS, with domain-specific analyses confirming significant benefits of VLFS in verbal fluency and cognitive flexibility. Processing Speed was significantly better with HFS vs. VLFS. No significant differences were observed across the remaining cognitive domains. *P<0.05. In Figure 3.A, cognitive flexibility, processing speed, and episodic memory and in Figure 3.B, episodic memory are based on single studies and therefore do not reflect meta-analytic results (indicated by smaller circles). **C. Effects of High vs. Low-Frequency STN-DBS on Verbal Fluency.** (C1) Forest plot showing Hedges’ g effect sizes for Verbal Fluency comparing HFS vs. LFS STN-DBS across studies. (C2) Pooled Hedges’ g estimate and 95%CI across a range of assumed within-subject correlations (r=0.0-0.9). (C3) Leave-one-study-out sensitivity analysis of the HFS vs. LFS comparison. **D. Effects of High vs. Very Low-Frequency STN-DBS on Verbal Fluency** (D1) Forest plot showing Hedges’ g effect sizes for Verbal Fluency comparing VLFS vs. HFS STN-DBS across studies. (E2) Pooled Hedges’ g estimate and 95%CI across a range of assumed within-subject correlations (r=0.0-0.9). (E3) Leave-one-study-out sensitivity analysis of the VLFS vs. HFS comparison. **E. Effects of High vs. Very Low-Frequency STN-DBS on Cognitive Flexibility.** (E1) Forest plot showing Hedges’ g effect sizes for Cognitive Flexibility comparing HFS vs. VLFS STN-DBS across studies. (E2) Pooled Hedges’ g estimate and 95%CI across a range of assumed within-subject correlations (r=0.0-0.9). (E3) Leave-one-study-out sensitivity analysis of the HFS vs. VLFS comparison. Omission of any single study did not reverse the direction of the effect, nor did it result in extreme changes in effect magnitude, suggesting that no study acted as a statistical outlier or exerted undue influence on the direction of the pooled estimate.

#### Verbal Fluency Subdomains

Verbal fluency subdomain analyses showed positive non-significant effect sizes (g=0.22-0.29), likely reflecting limited statistical power (Supplementary Figure 7).

#### Expanded domain classification: Primary+Secondary+Tertiary Domains

When all cognitive outcomes contributing to a given domain were pooled the results remained broadly consistent with the primary analyses, again indicating significant benefits of LFS for Verbal fluency and Cognitive flexibility (Supplementary Figure 8). Both domains showed similar effect sizes (g=0.24, p=0.031). All remaining domains demonstrated non-significant effects.

### High Frequency vs. Very Low Frequency STN-DBS

#### Main Cognitive Domains

Domain-specific analyses (Figure 3.B) indicated significant improvement under VLFS in Verbal Fluency (k=12, 5 studies, g=0.29, SE=0.08, 95% CI [0.14, 0.44], p<0.001) and Cognitive Flexibility (k=6, 4 studies, g=0.33, SE=0.11, 95% CI [0.13, 0.54], p=0.002). The pooled effect size remained consistently positive and statistically significant across correlation sensitivity analyses and leave-one-study-out iterations (Figures 3.D-E). In contrast, Processing speed was significantly worse under VLFS (k=7, 4 studies, g=-0.85, SE=0.43, 95% CI [-1.70, -0.0002], p<0.05). Other domains failed to present a statistically significant effect. Details on Supplementary Figure 9.

#### Verbal Fluency Subdomains

Across VF subdomains (Supplementary Figure 10), VLFS consistently outperformed HFS (Semantic VF, g=0.31, p=0.001 and Switching VF, g=0.32, p=0.0101). Letter VF showed a positive but non-significant effect size (g=0.20, p=0.214).

#### Executive Control Subdomains

When executive control tasks were stratified into accuracy- and time-based subdomains (Supplementary Figure 11), VLFS revealed a strong positive (beneficial) effect on accuracy measures, whereas a negative non-significant effect was observed for time-based measures (Accuracy, k=8, 6 studies, g=0.46, p=0.030 and Time, k=6, 5 studies, g=-0.12, p=0.722).

#### Expanded domain classification: Primary+Secondary+Tertiary Domains

When all cognitive outcomes contributing to a given domain were pooled, regardless of their hierarchical classification, the results remained broadly consistent with the primary analyses, again indicating significant benefits of VLFS for Verbal Fluency (g=0.30, p=6.7×10⁻⁶) and Cognitive Flexibility (g=0.30, p=1.4×10⁻⁶). All remaining domains showed non-significant effects. Details on Supplementary Figure 12.

Overall, across pooled, subgroups, sensitivity and robustness analyses, lower STN-DBS frequencies were consistently associated with modest improvements in verbal fluency and cognitive flexibility relative to HFS, with similar directional effects across verbal fluency subdomains and accuracy-based executive measures, while other cognitive domains showed no reliable frequency-related effects.

### OFF-Stimulation vs. High Frequency STN-DBS

#### Main Cognitive Domains

Figure 4.A-B presents a graphical summary of the meta-analysis comparing OFF stimulation with LFS+VLFS or HFS on cognitive outcomes in STN-DBS. Domain-specific analyses revealed a significantly worse performance in Verbal Fluency (Figure 4.C1) under HFS vs. OFF-stim (k=12, 5 studies, g=0.16, SE=0.08, 95% CI [0.01, 0.30], p=0.0385) and a non-significant trend for worsening of Cognitive Flexibility (k=6, 4 studies, g=0.22, SE=0.12, 95% CI [-0.02, 0.45], p=0.075). The result was robust across sensitivity analysis (Figure 4.C2, D2). In contrast, Processing Speed (Figure 4.D) showed a better performance with HFS (k=7, 4 studies, g=-0.77, SE=0.37, 95% CI [-1.49, -0.04], p=0.038). No significant results were found in the remaining domains (Supplementary Figure 13). Effect size was comparable across VF subdomains (Supplementary Figure 14).

**Figure 4:**
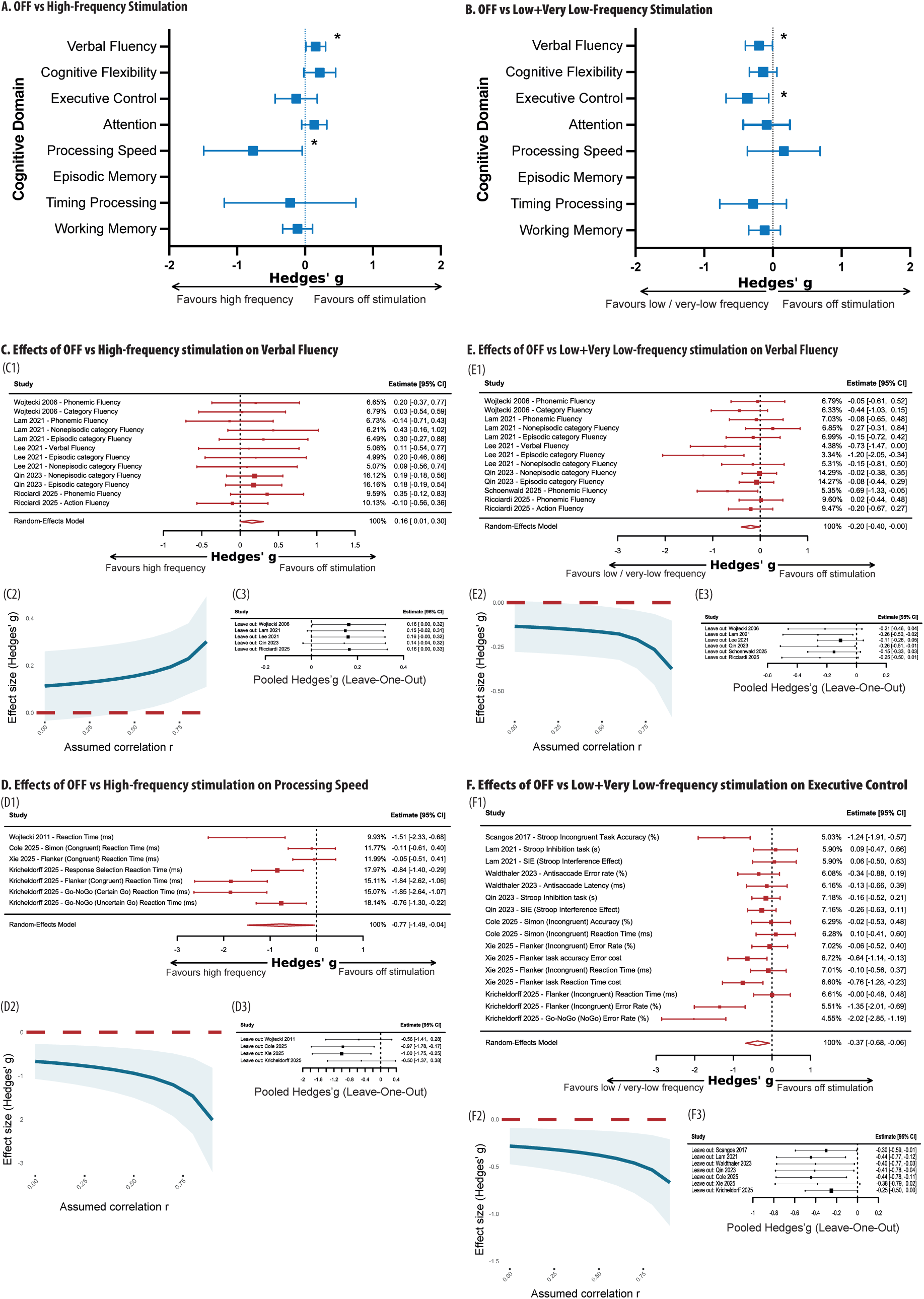
Effects of OFF vs On stimulation STN-DBS on Cognition. **A. Effects of OFF vs. High-Frequency on cognitive outcomes.** HFS was associated with significantly worse verbal fluency and better processing speed. **B. Effects of OFF vs. Low+Very Low-Frequency on cognitive outcomes.** LFS+VLFS were associated with significantly better performance on Verbal Fluency and Executive Control. No significant differences were observed in the remaining cognitive domains. *P<0.05. **C. Effects of OFF vs. High-Frequency STN-DBS on Verbal Fluency.** (C1) Forest plot showing Hedges’ g effect sizes for Verbal Fluency comparing OFF vs. HFS STN-DBS across studies. Positive values indicate better Verbal Fluency performance OFF relative to HFS stimulation. (C2) Pooled Hedges’ g estimate and 95%CI across a range of assumed within-subject correlations (r=0.0-0.9). (C3) Leave-one-study-out sensitivity analysis of the OFF vs. HFS comparison. **D. Effects of OFF vs. High-Frequency STN-DBS on Processing Speed.** (D1) Forest plot showing Hedges’ g effect sizes for Processing Speed comparing OFF vs. HFS STN-DBS across studies. (D2) Pooled Hedges’ g estimate and 95%CI across a range of assumed within-subject correlations (r=0.0-0.9). (D3) Leave-one-study-out sensitivity analysis of the Off vs. HFS comparison. **E. Effects of OFF vs. Low+Very Low-Frequency STN-DBS on Verbal Fluency.** (E1) Forest plot showing Hedges’ g effect sizes for Verbal Fluency comparing OFF vs. LFS+VLFS STN-DBS across studies. (E2) Pooled Hedges’ g estimate and 95%CI across a range of assumed within-subject correlations (r=0.0-0.9). (B3) Leave-one-study-out sensitivity analysis of the OFF vs. LFS+VLFS comparison. **F. Effects of OFF vs. Low+Very Low-Frequency STN-DBS on Executive Control.** (F1) Forest plot showing Hedges’ g effect sizes for Executive Control comparing OFF vs. LFS+VLFS STN-DBS across studies. (F2) Pooled Hedges’ g estimate and 95%CI across a range of assumed within-subject correlations (r=0.0-0.9). (F3) Leave-one-study-out sensitivity analysis of the Off vs. LFS+VLFS comparison.

#### Expanded domain classification: Primary+Secondary+Tertiary Domains

Comparing OFF-stim vs. HFS conditions, across all categories, the results supported a significant deterioration under HFS for Verbal Fluency and Cognitive Flexibility (Supplementary Figure 15). Both domains showed robust positive effect with OFF-stim (Verbal Fluency, g=0.15, SE=0.06, 95% CI [0.02, 0.28], p=0.020 and Cognitive Flexibility, g=0.17, SE=0.06, 95% CI [0.05, 0.29], p=0.0043). Other domains showed non-significant effects.

### OFF-Stimulation vs. Low+Very Low Frequency STN-DBS

#### Main Cognitive Domains

Domain-specific analyses indicated significantly better performance with LFS+VLFS vs. OFF-stim in Verbal Fluency (k=13, 6 studies, g=-0.20, SE=0.10, 95% CI [-0.40, -0.005], p=0.045) and Executive Control (k=16, 7 studies, g=-0.37, SE=0.16, 95% CI [-0.68, -0.06], p=0.0181). These effects were robust across sensitivity analysis (Figure 4.E-F). No other significant results were identified in the remaining domains (Supplementary Figure 16).

Effect size of verbal fluency subdomains were comparable, although not significant, with effect sizes ranging from -0.15 to -0.19 (Supplementary Figure 17). For executive control, subdomain analyses suggested the effect was driven by a better performance with LFS+VLFS relative to OFF-stim for accuracy-based measures (g=-0.54, p=0.057) and not time-based measures (g=-0.14, p=0.195) (Supplementary Figure 18).

#### Expanded domain classification: Primary+Secondary+Tertiary Domains

In the broader, more inclusive analysis, comparing OFF stimulation with LFS+VLFS consistent results were identified (Supplementary Figure 19). Significant improvements were observed for Verbal Fluency (g=-0.21, SE=0.09, 95% CI [-0.40, -0.03], p=0.021), and Cognitive Flexibility (g=-0.18, SE=0.07, 95% CI [-0.33, -0.03], p=0.016) with Executive Control showing a trend toward significance (g=-0.34, SE=0.18, 95% CI [−0.69, 0.02], p=0.064). The remaining domains did not show significant differences.

#### Study Design and Medication Conditions Variables

At last, we examined whether the pooled effects observed for OFF-stim vs. HFS and OFF-stim vs. LFS+VLFS were disproportionately influenced by study design or medication status. Most studies were randomized within-subject snd double-blind, and all but one^67^ applied bilateral STN stimulation.

Using Robust Variance Estimation (RVE) with medication status as a moderator no significant effect of medication status was observed on verbal fluency and processing speed effect size (p=0.955 and p=0.720, respectively) for the OFF-stim vs. HFS comparison. Similarly, for the OFF-stim vs. LFS+VLFS comparison, medication status had no significant effect on verbal fluency and executive control (p=0.356 and p=0.862).

Taken together, analyses suggest that HFS worsens verbal fluency while improving processing speed when compared with OFF, while LFS+VLFS showed improvement of verbal fluency, executive control, and cognitive flexibility when compared with OFF stimulation.

#### STN-DBS studies not included in the meta-analysis

The 10 studies excluded from the meta-analysis provided complementary evidence. Only one study^69^ assessed both cognitive flexibility and verbal fluency, comparing combined simultaneous VLFS+HFS with HFS alone. VLFS+HFS improved verbal fluency but had no significant effect on cognitive flexibility, a pattern partially consistent with our main meta-analytic findings.

Regarding executive control, two studies^37,68^ reported no significant differences on accuracy when comparing LFS to HFS. For attention, electrophysiological evidence suggested that 60 Hz had a milder impact on P300 latency compared to significant increases at 130 Hz, suggesting a relative preservation of attentional circuits.^57^

Research on emotional processing suggests that 10 Hz stimulation may reduce negative bias and increase the subjective pleasantness of imagery.^54,60,61^ Similarly, low-frequency stimulation was associated with improved mood, specifically regarding depressive symptoms^48,53,69^ and hypomania^47,69^. Finally, data on sleep indicate that while low-frequency stimulation may not improve total sleep efficiency^50^, it significantly modulates NREM oscillations^63,64^ and may enhance next-day memory retention^64^.

Overall, these qualitative findings broadly support the meta-analytic results while highlighting additional domain-specific and mechanism-level effects of low-frequency STN-DBS.

### Pedunculopontine nucleus (PPN)

Regarding PPN-DBS a high heterogeneity exists, with a maximum of two studies addressing any single domain, which limits pooled comparisons (Supplementary Table 2).

A standard frequency of 10-25 Hz for PPN-DBS was mostly commonly applied and compared with OFF-stim. For verbal fluency and executive function, stimulation at 25 Hz has been shown to significantly improve phonemic fluency and Trail Making Test-B performance compared with OFF-stim.^74,75^ For reaction time, two studies compared higher and lower frequencies. Only 20-35 Hz significantly improved simple reaction time compared with OFF-stim and 5-10 Hz^78^ and similarly 8-20 Hz showed faster reaction times in alertness tasks compared to OFF-stim and 60-130 Hz.^79^ However, no effects on more complex reaction time paradigms were found.

Regarding working memory, compared with OFF-stim, 25 Hz stimulation significantly reduced reaction times during n-back working memory tasks without changes in accuracy^74^, while 20-40 Hz selectively enhances reward-based learning^81^. One study showed improved delayed recall with 25 Hz compared with OFF^76^, alongside improved sleep efficiency. In contrast, 60-80 Hz stimulation was associated with increased daytime sleepiness compared with 10-25 Hz^77,82^. Lastly, 8-10 Hz showed to increase perceptual thresholds in visual contrast sensitivity relative to OFF and 60 Hz conditions.^80^

### Nucleus basalis of Meynert (NBM)

Only three studies^83–85^ investigated NBM-DBS in PD patients (Supplementary Table 2), but no comparisons across NBM frequencies were performed (only vs. OFF-stim). Frequencies included 15Hz^84,85^, 20Hz^83^ and 60Hz ^84,85^. These studies suggest that the procedure is safe and well-tolerated; however, effects were mostly modest and inconsistent.

A descriptive qualitative synthesis of the included studies, detailing findings, clinical and demographic characteristics, and DBS parameters, is provided in Supplementary Tables 3-5.

## Discussion

The present systematic review and meta-analysis aimed to clarify the cognitive effects of stimulation frequency across multiple domains. Our results suggest that lower STN-DBS frequencies are associated with modest but consistent benefits on verbal fluency, cognitive flexibility and potentially benefits on accuracy-based executive control metrics when compared with HFS (Table 2). These findings are supported by consistent effects across sensitivity analyses, directionally concordant comparisons against OFF stimulation, and apparent independence from medication state. Importantly, these effects were not generalized: working memory, attention, episodic memory, and time processing showed no reliable frequency-related effects, whereas processing-speed performance was better with HFS.

**Table 2:** Summary of meta-analysis results. Summary of meta-analysis results comparing low- and very low-frequency stimulation (LFS+VLFS) vs. high-frequency stimulation (HFS), OFF vs. HFS, and OFF vs. LFS+VLFS across main cognitive domains and subdomains in STN-DBS. LFS and VLFS were associated with modest cognitive benefits compared with both HFS (positive values indicate better performance with LFS+VLFS vs. HFS) and OFF stimulation (negative values indicate better performance with LFS+VLFS vs. OFF-Stim), particularly in verbal fluency, as well as in cognitive flexibility and accuracy-based executive control metrics. Differences in other cognitive domains were non-significant. These findings are consistent with the role of the subthalamic nucleus as a key multifunctional node within fronto-striatal circuits, contributing to executive processes. **\*\*p<0.05; ^$^p=0.05-0.06; others p>0.06**

| Main domain | High vs. Low+Very Low Frequency |  |  | ON stim vs OFF stim |  |
| --- | --- | --- | --- | --- | --- |
|  | High vs. Low+Very Low | High vs. Low | High vs. Very Low | High vs. OFF | Very Low/Low vs. OFF |
| Verbal Fluency | 0.27 [0.16, 0.38]** | 0.23 [0.02, 0.44]** | 0.29 [0.14, 0.44]** | 0.16 [0.01, 0.30]** | -0.20 [-0.40, -0.005]** |
| Semantic VF | 0.30 [0.15, 0.45]** | 0.27 [-0.01, 0.56]<sup>§</sup> | 0.31 [0.12, 0.51]** | 0.20 [0.01, 0.39]** | -0.17 [-0.42, 0.09] |
| Letter VF | 0.22 [0.05, 0.40]** | 0.22 [-0.04, 0.48] | 0.20 [-0.11, 0.50] | 0.17 [-0.14, 0.47] | -0.15 [-0.43, 0.12] |
| Switching VF | 0.34 [0.10, 0.58]** | - | 0.32 [0.08, 0.58]** | 0.13 [-0.11, 0.38] | -0.19 [-0.43, 0.06] |
| Cognitive Flexibility | 0.38 [0.09, 0.68]** | - | 0.33 [0.13, 0.54]** | 0.22 [-0.02, 0.45]<sup>§</sup> | 0.14 [-0.34, 0.06] |
| Executive Control | 0.18 [-0.05, 0.40] | -0.00 [-0.20, 0.19] | 0.26 [-0.04, 0.56] | -0.13 [-0.44, 0.18] | -0.37 [-0.68, -0.06]** |
| Accuracy EC | 0.35 [0.04, 0.65]** | - | 0.46 [0.06, 0.86]** | - | -0.54 [-1.09, 0.02]<sup>§</sup> |
| Time EC | -0.10 [-0.62, 0.42] | - | -0.12 [-0.95, 0.71] | - | -0.014 [-0.36, 0.09] |
| Processing Speed | -0.67 [-1.43, 0.10] | - | -0.85 [-1.70, -0.0002]** | -0.77 [-1.49, -0.04]** | 0.16 [-0.37, 0.69] |
| Attention | 0.25 [-0.22, 0.72] | - | 0.25 [-0.22, 0.72] | 0.14 [-0.05, 0.32] | -0.09 [-0.43, 0.25] |
| Episodic Memory | -0.12 [-1.69, 1.46] | - | - | - | - |
| Working Memory | 0.01 [-0.22, 0.24] | -0.22 [-0.61, 0.18] | 0.09 [-0.15, 0.33] | -0.11 [-0.33, 0.11] | -0.14 [-0.35, 0.11] |
| Timing Processing | -0.12 [-1.69, 1.46] | - | -0.12 [-1.69, 1.46] | -0.22 [-1.19, 0.75] | -0.29 [-0.77, 0.20] |

■ Favours Low + Very-Low Frequency ■ Favours High Frequency ■ Favours Off Stimulation

These results suggest that stimulation frequency may modulate cognitive processes in patients undergoing STN-DBS rather than acting only as a motor programming parameter. The clearest signal concerned verbal fluency, a domain shown to decline after STN-DBS and often considered one of the most reproducible adverse effects of the procedure.^14–16^ Beyond its linguistic component, verbal fluency is a sensitive marker of executive function, requiring self-initiated search, response selection, and monitoring.^42,43,86,87^ Its convergence with effects on cognitive flexibility and accuracy-based executive control therefore points to a frequency-sensitive cognitive-executive profile.^42,43^ This interpretation is further supported by the OFF-stimulation comparisons: HFS was associated with worse verbal fluency relative to OFF-stimulation, whereas LFS+VLFS showed better performance than OFF-stimulation for verbal fluency and executive control. Taken together, this pattern is consistent with the possibility that lower frequencies may mitigate part of the HFS-related verbal fluency cost while preserving or enhancing selected cognitive-executive processes.

These findings may have clinical relevance given the well-established association between executive dysfunction, gait impairment and FoG in PD^34–36^. Thus, the effects observed on verbal fluency, cognitive flexibility, and accuracy-based executive control suggest a potential cognitive-motor bridge between frequency-dependent STN modulation and the reported axial benefits of LFS. This interpretation remains indirect, however as the included studies did not stratify cognitive outcomes by freezing status, axial phenotype, or objective gait measures.

The present findings provide a direction of effect, but not yet a full mechanistic explanation. A key next step is to move beyond frequency as an isolated programming variable and consider how frequency interacts with stimulation location, tissue activation, and the functional topography of the STN. The STN is not a homogeneous motor structure^13,17–19,88–90^: dorsolateral regions are more closely linked to sensorimotor loops, whereas rostral and ventromedial territories are more strongly connected with associative and limbic circuits ^18,19,89,90^. Accordingly, the same stimulation frequency may have different cognitive or motor consequences depending on whether stimulation predominantly engages motor, associative, or overlapping cognitive-motor territories. In the case of posture and gait, fibers projecting also from STN to brainstem locomotor regions must also be considered.^11,26,32,91–94^

Only studies that systematically assess and manipulate both the location of the VTA and stimulation frequency will be able to provide more robust and mechanistically grounded findings.

This spatial specificity is further shaped by oscillatory mechanisms. HFS may effectively suppress pathological beta synchronization in sensorimotor circuits^95–99^, while lower frequencies may preserve or interact with theta and alpha dynamics relevant to cognitive control, response selection, and cognitive-motor adaptation^100–112^. Mechanistic studies integrating frequency, anatomy, and oscillatory dynamics are needed.

Evidence for non-STN targets was too limited for firm conclusions but supports the need for target-specific interpretation. The lack of studies involving the globus pallidum internus is an important limitation, that would help in understanding the cognitive-motor integration. Small PPN-DBS studies suggest frequency may influence reaction time, working memory, and gait-related cognitive processes, while NBM-DBS evidence was even more limited. Frequency-dependent cognitive effects should not be assumed to generalize across DBS targets.

Several limitations deserve acknowledgment. Some domain-level estimates were based on few studies and participants. Neuropsychological tasks are not process-pure, and time-based measures may be influenced by motor or speech changes. DBS effects depend on multiple interacting parameters. Although total equivalent energy delivered (TEED) is an important reporting metric, matching TEED across frequency conditions would require changing other parameters and could obscure the specific contribution of frequency. Importantly, most studies kept amplitude and pulse width constant, facilitating interpretation of frequency as the primary manipulation with only 5 studies^37,52,54,57,59^ adjusting parameters according to TEED (and only 2 of those included in the meta-analysis^52,59^). DBS effects should be understood as consequences of changing temporal stimulation pattern rather than simple energy differences. Although LFS and VLFS were combined it must be considered they can involve distinct physiological mechanisms. Finally, most studies lacked sufficient anatomical or electrophysiological detail, and none tested whether cognitive effects varied by FOG status or objective gait performance. These limitations mean that these findings should be considered hypothesis-generating and require replication in larger, blinded, anatomically informed studies. More broadly, these findings raise a provocative but testable possibility: lower-frequency stimulation may do more than mitigate HFS-related cognitive costs; in selected contexts, it may facilitate cognitive-executive processing. This interpretation aligns with non-invasive neuromodulation studies showing that specific modulation of frontoparietal networks can improve verbal fluency^113^ or executive functions^114,115^.

In conclusion, lower-frequency STN-DBS was associated with modest advantages in verbal fluency, cognitive flexibility, and accuracy-based executive control compared with conventional HFS. These findings suggest that stimulation frequency may shape cognitive-executive processes relevant to both neuropsychological performance and cognitive-motor regulation in PD. Rather than supporting a generalized cognitive benefit of LFS, our results point to a selective, frequency-sensitive profile that warrants mechanistic testing. Future studies should combine systematic frequency manipulation with anatomically informed contact localization, electrophysiological recordings, and integrated cognitive and gait assessments, including dual-task and FOG-provoking paradigms. This approach may help define patient-specific stimulation profiles that optimize appendicular motor control while preserving or enhancing cognitive-executive and axial functions.

## Data Availability

This study was based on a systematic review, and all data used were derived from previously published studies and are publicly available in the scientific literature. Extracted data are from published studies and are available in the paper and supplementary data. The data that support the findings of this study are available from the corresponding author, upon reasonable request.

## Data availability

Extracted data are from published studies and are available in the paper and supplementary data. The data that support the findings of this study are available from the corresponding author, upon reasonable request.

## Author contributions

B.M.: conceptualization, project administration, methodology, screening and data extraction, statistical analysis, review and critique of results, drafted the manuscript, approved the final version.

P.B.: statistical analysis, methodology, review and critique of results, approved the final version.

V.M.F.: screening and data extraction, review and critique of results, approved the final version.

J.A.: screening and data extraction, review and critique of results, approved the final version.

M.M.: data extraction, review and critique of results, approved the final version.

R.L.: methodology, definition of cognitive performance measures, review and critique of results, approved the final version.

R.B.: review and critique of results, approved the final version.

M.C.: review and critique of results, approved the final version.

M.D. M.: conceptualization, methodology, supervision project, review and critique of results, drafted the manuscript, approved the final version.

## Supplementary information

The online version contains supplementary material.

Supplementary Methods Description

Supplementary Table 1-5

Supplementary Figures 1-19

