## Supplementary Methods for "Frequency-dependent cognitive effects of Deep Brain Stimulation in Parkinson’s Disease: A Systematic Review and Meta-Analysis"

### META-ANALYSIS METHODS

#### Data Preprocessing and Effect Size Calculation

#### Data Sources and Selection

We collected data from 21 studies reporting cognitive outcomes under different subthalamic nucleus deep brain stimulation (STN-DBS) conditions: OFF, Very Low Frequency (VLFS), Low Frequency (LFS) and High Frequency (HFS) stimulation.

#### Study Quality Assessment

The methodological quality of the included studies was assessed using the Cochrane Risk of Bias 2 (RoB-2) tool for randomized and crossover trials and the ROBINS-I tool for non-randomized studies.

#### Contrast Generation

All pairwise contrasts within each study and cognitive task were generated to create an effect ID for each unique pair of conditions. Each contrast included: condition A vs. B labels, means, standardized deviations (SDs), and sample sizes. Study-level metadata (study design, cognitive domain, cognitive task, medication state, stimulation side) were likewise recorded per contrast.

#### Deep Brain stimulation features definition

Stimulation targets and frequency were extracted. For STN-DBS, stimulation frequency was classified in very low (VLFS, 4-10 Hz), low (LFS, 60-80 Hz), high (HFS, ≥130 Hz), and OFF stimulation.

#### Cognitive outcomes definition

To enable consistent synthesis across studies and to avoid limitations of grouping findings solely by task design, we classified extracted outcomes (rather than tasks). Different measures derived from the same task (e.g., accuracy, reaction time, error rate, or interference cost) were treated as distinct outcomes indexing different cognitive processes. Outcomes were mapped onto a previously published eight-domain framework comprising verbal fluency, working memory, cognitive flexibility, executive control, attention, processing speed, episodic memory, and temporal processing. Each outcome was assigned a primary domain, and when relevant, additional secondary and tertiary domains were recorded to capture less central but meaningful cognitive demands (see Supplementary Table 1). Main analyses were focused on the primary domain. Secondary and tertiary domains were used for robustness analysis presented as expanded domain classification analysis. Subdomains (i.e. Letter or Semantic fluency) were also identified and used for sensitivity analysis. Additionally, executive control subdomains were further stratified into accuracy-based measures (e.g., accuracy or error rate) and time-based measures (e.g., reaction time).

#### Coding Cognitive Outcomes

Cognitive tasks were coded for whether higher scores reflect better performance (higher_is_better) based on task-specific criteria. Tasks measuring accuracy or correct responses were coded TRUE; tasks measuring time or error rates were coded FALSE.

**Effect Size Computation**

Effect sizes were calculated as Hedges' g for each contrast.

- **Within-subject contrasts**:

$$g = J \cdot \frac{\bar{X}_A - \bar{X}_B}{\sqrt{\text{SD}_A^2 + \text{SD}_B^2 - 2r \cdot \text{SD}_A \cdot \text{SD}_B}}, \text{Var}(g) = \frac{1}{n} + \frac{g^2}{2n}, J = 1 - \frac{3}{4n-1}$$

where $r = 0.5$ is the assumed correlation between repeated measures (but subsequently stress tested for different values of $r$).

- **Between-subject contrasts**:

$$g = J \cdot \frac{\bar{X}_A - \bar{X}_B}{S_{\text{pooled}}}, \text{Var}(g) = \frac{n_A + n_B}{n_A n_B} + \frac{g^2}{2(n_A + n_B)}, S_{\text{pooled}} = \sqrt{\frac{(n_A - 1)\text{SD}_A^2 + (n_B - 1)\text{SD}_B^2}{n_A + n_B - 2}}, J = 1 - \frac{3}{4(n_A + n_B) - 9}$$

Effect sizes were calculated in the direction of cognitive benefit: for tasks where higher values indicate worse performance (e.g., reaction time), the sign of g was flipped so that positive values always indicate better cognition in condition A relative to condition B.

**Meta-Analysis of Low+Very Low Frequency, High Frequency and OFF STN-DBS**

**Data Selection**

For the comparison of Low+Very Low Frequency vs High Frequency, Low Frequency vs High Frequency, Very Low Frequency vs High Frequency, and OFF vs High Frequency and OFF vs Low+Very Low Frequency stimulation, contrasts were extracted from the pre-processed dataset and analysed using two independent meta-analyses. Only rows where the contrast involved Low+Very Low Hz vs High Hz, Low Hz vs High Hz, Very Low Hz vs High Hz, or OFF vs ON (depending on the analysis) were included.

**Multilevel Random-Effects Meta-Analysis**

Several studies contributed multiple cognitive outcomes, resulting in statistical dependence among effect sizes within studies. So, we conducted multilevel meta-analyses using the metafor package in R to synthesize effect sizes while accounting for dependencies among observations. Given that multiple cognitive outcomes were assessed within the same studies, we employed this hierarchical modelling framework that explicitly accounts for this dependency structure: this model more closely accounted for the dependency among effect sizes originating from the same participants by clustering at the study level, with individual effect sizes nested within studies. This accounted for potential dependencies among multiple outcomes from the same cognitive task (when applicable) and addresses the more substantial dependency arising from multiple tests administered to the same participants within a study. This model estimated variance components using restricted maximum likelihood (REML), which provides less biased estimates of variance parameters compared to maximum likelihood, particularly with smaller numbers of clusters.

**Domain-Specific Analyses**

Effect sizes were aggregated within cognitive domains (verbal fluency, cognitive flexibility, executive control, working memory, attention, processing speed, timing processing, and episodic memory). Only domains with ≥2 contrasts were included in the analyses. The same multilevel model structure was applied, and forest plots were generated for each domain. For verbal fluency and executive control, subdomain analyses were conducted, including verbal fluency categories (semantic, phonemic, switching and action) and accuracy- and time-based measures of executive control. When tasks engaged multiple cognitive domains, a hierarchical classification (primary, secondary, and tertiary domains) was applied. This approach increased the number of contrasts per domain and was used as a sensitivity analysis to assess the robustness of the findings.

The principal outcome (key comparison) was the contrast between HFS vs. combined LFS+VLFS for primary cognitive domains. Secondary outcome included HFS vs. LFS and HFS vs. VLFS separately. Exploratory outcomes included OFF vs. HFS, and OFF vs. LFS+VLFS.

Significance level was set at 0.05. To account for multiple testing across the different cognitive domains, Benjamini-Hochberg false discovery rate correction was applied within the primary analysis (key comparison HFS vs. LFS+VLFS). Domains with $q < 0.05$ were considered statistically significant after correction. Secondary outcomes (HFS vs. LFS, HFS vs. VLFS, OFF vs High, OFF vs LFS+VLFS) are considered secondary/exploratory and are presented without correction for multiple comparisons.

**Visualization**

Forest plots were generated for the overall contrasts and for each cognitive domain with at least two effect sizes. The x-axis represented Hedges' g, where positive values indicate better cognitive performance under Low+Very Low Hz stimulation relative to High Hz stimulation settings or better cognitive performance under OFF relative to ON stimulation settings.

**Heterogeneity Measures**

Between-study variance ($\tau^2$) and the $I^2$ statistic were calculated as:

$$I^2 = 100 \cdot \frac{\tau^2}{\tau^2 + \bar{v}}$$

where $\bar{v}$ is the mean of the within-study variances (vi).

**Sensitivity Analysis to Assumed Correlation (r)**

For within-subject contrasts, effect size variance depends on the correlation between repeated measures. To assess sensitivity, we recalculated Hedges' g and its variance for a range of assumed correlations $r = 0.0, 0.1, \ldots, 0.9$ using the above specified Hedges' g calculation formulas. The sensitivity analysis confirmed that results were robust across a plausible range of $r$ values for both the Low+Very Low Hz vs High Hz; Very Low Hz vs High Hz and Low Hz vs High Hz comparisons, as well as OFF vs ON stimulation.

**Leave-One-Study-Out Analysis**

Several studies contributed multiple cognitive outcomes, resulting in statistical dependence among effect sizes within studies. Although this dependency was addressed in the primary analysis using a multilevel random-effects meta-analysis, it remained important to evaluate whether the overall pooled effect was disproportionately influenced by any single study especially given the moderate heterogeneity. Given the relatively small number of contributing studies and the modest magnitude of the pooled effect, we conducted influence diagnostics to assess robustness of the findings to the exclusion of individual studies.

To evaluate the influence of individual studies, we performed a leave-one-study-out sensitivity analysis, whereby the multilevel meta-analysis model was refit repeatedly after excluding all effect sizes from one study at a time. In each iteration, the same model specification was retained, including the multilevel random-effects structure accounting for effect sizes nested within studies. For each refitted model, we extracted the pooled effect size (Hedges' g), its standard error, and the corresponding 95%.

**Robust Variance Estimation (RVE)**

In addition to the multilevel meta-analysis described above, and to appropriately address the dependency arising from multiple effect sizes contributed by the same study, we conducted a Robust Variance Estimation (RVE) meta-analysis with small-sample corrections. This approach allowed medication status (MedON vs. MedOFF) to be examined as a moderator within a single model, rather than stratifying the dataset into separate subgroup analyses. By retaining all available effect sizes simultaneously, RVE preserves statistical power, which is particularly relevant given the limited number of studies and comparisons available. Furthermore, this method accounts for the correlation between non-independent effect sizes within studies and applies small-sample adjustments to the variance estimates and degrees of freedom, thereby improving the accuracy of statistical inference under sparse data conditions. The moderator coefficient was interpreted to determine whether medication status significantly influenced the pooled treatment effect.
