## Supplementary Table for "Frequency-dependent cognitive effects of Deep Brain Stimulation in Parkinson’s Disease: A Systematic Review and Meta-Analysis"

**Supplementary Table 1:** Stratification of cognitive domains of the STN-DBS studies included in the meta-analysis

| Study name | Cog Task | Main_Domain | Main_Subdomain | Secondary_Domain | Tertiary_Domain | Metric |
|---|---|---|---|---|---|---|
| **Wojtecki 2006** | Phonemic Fluency | Verbal Fluency | VF (letter) | Cognitive Flexibility | | Total correct |
| | Category Fluency | Verbal Fluency | VF (semantic) | Cognitive Flexibility | | Total correct |
| | Alternating phonemic Fluency | Cognitive Flexibility | VF (switching) | Verbal Fluency | | Switching total correct |
| | Alternating category Fluency | Cognitive Flexibility | VF (switching) | Verbal Fluency | | Switching total correct |
| **Wojtecki 2011** | Time reproduction task (15 s) | Timing Processing | | | | Performance |
| | Time production task (15 s) | Timing Processing | | | | Performance |
| | Time discrimination task (correct judgements) | Timing Processing | | | | Performance |
| | Tapping unpaced task (ms) | Timing Processing | | | | Performance |
| | Reaction Time (ms) | Processing Speed | | Attention (bottom up) | | RT |
| **Stegemoller 2013** | Phonemic Fluency | Verbal Fluency | VF (letter) | Cognitive Flexibility | | Total correct |
| | Category Fluency | Verbal Fluency | VF (semantic) | Cognitive Flexibility | | Total correct |
| **Fagundes 2016** | Phonemic Fluency, P | Verbal Fluency | VF (letter) | Cognitive Flexibility | | Total correct |
| | Phonemic Fluency, FAS | Verbal Fluency | VF (letter) | Cognitive Flexibility | | Total correct |
| | Category Fluency | Verbal Fluency | VF (semantic) | Cognitive Flexibility | | Total correct |
| | Action Fluency | Verbal Fluency | VF (action) | Cognitive Flexibility | | Total correct |
| | Unconstrained Fluency | Verbal Fluency | VF (unconstrained) | Cognitive Flexibility | | Total correct |
| **Amara 2016** | Psychomotor vigilance task | Processing Speed | | Attention (bottom up) | | RT |
| **Scangos 2017** | Stroop Incongruent Task Accuracy (%) | Executive Control | | | | Accuracy |
| **Kelley 2018** | Time production task (3 s) | Timing Processing | | | | Performance |
| | Time production task (12 s) | Timing Processing | | | | Performance |
| **Grover 2018** | Phonemic Fluency | Verbal Fluency | VF (letter) | Cognitive Flexibility | | Total correct |
| | Category Fluency | Verbal Fluency | VF (semantic) | Cognitive Flexibility | | Total correct |
| **Lam 2021** | Phonemic Fluency | Verbal Fluency | VF (letter) | Cognitive Flexibility | | Total correct |
| | Nonepisodic Category Fluency | Verbal Fluency | VF (semantic) | Cognitive Flexibility | | Total correct |
| | Episodic Category Fluency | Verbal Fluency | VF (semantic) | Cognitive Flexibility | | Total correct |
| | Alternating Category Fluency | Cognitive Flexibility | VF (switching) | Verbal Fluency | | Switching total correct |
| | Stroop Word Reading (s) | Attention | | Processing Speed | | Time |
| | Stroop Color Naming (s) | Attention | | Processing Speed | | Time |
| | Stroop Inhibition task (s) | Executive Control | | Attention (top down) | | Time |
| | SIE (Stroop Interference Effect) | Executive Control | | Attention (top down) | | Interference cost |
| | Random Number Generation (Evan's RNG scores) | Working Memory | | Executive Control | Attention (top down) | Score |
| **Lee 2021** | Verbal Fluency | Verbal Fluency | VF (unspecified) | Cognitive Flexibility | | Total correct |
| | Episodic Fluency | Verbal Fluency | VF (semantic) | Cognitive Flexibility | | Total correct |
| | Nonepisodic Fluency | Verbal Fluency | VF (semantic) | Cognitive Flexibility | | Total correct |
| **Waldthaler 2023** | Antisaccade Error rate (%) | Executive Control | | Attention (top down) | | Error rate |
| | Antisaccade Latency (ms) | Executive Control | | Attention (top down) | | Latency |
| **Zacharia 2023** | Antisaccade Error rate (%) | Executive Control | | Attention (top down) | | Error rate |
| | Stroop Inhibition task (s) | Executive Control | | Attention (top down) | | Time |
| | Stroop effect 2 | Executive Control | | Attention (top down) | | Interference cost |
| | Stroop effect 3 | Executive Control | | Attention (top down) | | Interference cost |
| **Qin 2023** | Nonepisodic Category Fluency | Verbal Fluency | VF (semantic) | Cognitive Flexibility | | Total correct |
| | Episodic Category Fluency | Verbal Fluency | VF (semantic) | Cognitive Flexibility | | Total correct |
| | Alternating Category Fluency | Cognitive Flexibility | VF (switching) | Verbal Fluency | | Switching total correct |
| | Stroop Word Reading (s) | Attention | | Processing Speed | | Time |
| | Stroop Color Naming (s) | Attention | | Processing Speed | | Time |
| | Stroop Inhibition task (s) | Executive Control | | Attention (top down) | | Time |

| | | | | | | |
|---|---|---|---|---|---|---|
| | SIE (Stroop Interference Effect) | Executive Control | | Attention (top down) | | Interference cost |
| | Forward Digital Span | Attention | | | | Span length |
| | SDMT (Symbol Digital Switch Test) | Attention | | Working Memory | | Total correct |
| | Backward Digital Span | Working Memory | | | | Span length |
| **Herz 2024** | Delay Recall | Verbal Episodic Memory | | | | Total correct |
| **Salehi 2024** | Sternberg working memory task | Working Memory | | | | Total correct |
| **Busteed 2024** | Phonemic Fluency, total | Verbal Fluency | VF (letter) | Cognitive Flexibility | | Total correct |
| | Phonemic Fluency, P | Verbal Fluency | VF(letter) | Cognitive Flexibility | | Total correct |
| | Phonemic Fluency, M | Verbal Fluency | VF (letter) | Cognitive Flexibility | | Total correct |
| | Category Fluency, Total | Verbal Fluency | VF (semantic) | Cognitive Flexibility | | Total correct |
| | Category Fluency, Animals | Verbal Fluency | VF (semantic) | Cognitive Flexibility | | Total correct |
| | Category Fluency, fruits and vegies | Verbal Fluency | VF (semantic) | Cognitive Flexibility | | Total correct |
| | Action Fluency | Verbal Fluency | VF (action) | Cognitive Flexibility | | Total correct |
| | Alternating Fluency | Cognitive Flexibility | VF (switching) | Verbal Fluency | | Switching total correct |
| | LetterNumber Sequence | Working Memory | | | | Total correct |
| | Delay Recall | Verbal Episodic Memory | | | | Total correct |
| **Schoenwald 2025** | Phonemic Fluency | Verbal Fluency | VF (letter) | Cognitive Flexibility | | Total correct |
| **Cole 2025** | Simon (Congruent) Accuracy (%) | Attention | | Processing Speed | | Accuracy |
| | Simon (Incongruent) Accuracy (%) | Executive Control | | Attention (top down) | | Accuracy |
| | Simon (Congruent) Reaction Time (ms) | Processing Speed | | Attention | | RT |
| | Simon (Incongruent) Reaction Time (ms) | Executive Control | | Attention (top down) | | RT |
| **Ricciardi 2025** | Phonemic Fluency | Verbal Fluency | VF (letter) | Cognitive Flexibility | | Total correct |
| | Action Fluency | Verbal Fluency | VF (action) | Cognitive Flexibility | | Total correct |
| | N-back task Correct (%) | Working Memory | | | | Accuracy |
| **Xie 2025** | Flanker (Congruent) Error Rate (%) | Attention | | Processing Speed | | Error rate |
| | Flanker (Incongruent) Error Rate (%) | Executive Control | | Attention (top down) | | Error rate |
| | Flanker Error Rate Cost | Executive Control | | Attention (top down) | | Error rate cost |
| | Flanker (Congruent) Reaction Time (ms) | Processing Speed | | Attention | | RT |
| | Flanker (Incongruent) Reaction Time (ms) | Executive Control | | Attention (top down) | | RT |
| | Flanker task reaction time cost | Executive Control | | Attention (top down) | | RT cost |
| **Kricheldorff 2025** | Response Selection Reaction Time (ms) | Processing Speed | | Attention | | RT |
| | Response Selection Error Rate (%) | Attention | | Processing Speed | | Error rate |
| | Flanker (Congruent) Reaction Time (ms) | Processing Speed | | Attention | | RT |
| | Flanker (Congruent) Error Rate (%) | Attention | | Processing Speed | | Error rate |
| | Flanker (Incongruent) Reaction Time (ms) | Executive Control | | Attention (top down) | | RT |
| | Flanker (Incongruent) Error Rate (%) | Executive Control | | Attention (top down) | | Error rate |
| | Go-NoGo (Certain Go) Reaction Time (ms) | Processing Speed | | Attention | | RT |
| | Go-NoGo (Certain Go) Error Rate (%) | Attention | | Processing Speed | | Error rate |
| | Go-NoGo (Uncertain Go) Reaction Time (ms) | Processing Speed | | Executive Control | Working Memory | RT |
| | Go-NoGo (Uncertain Go) Error Rate (%) | Attention | | Executive Control | Working Memory | Error rate |
| | Go-NoGo (NoGo) Error Rate (%) | Executive Control | | Attention (top down) | Working Memory | Error rate |
| | Stop-Change SCRT (ms) | Cognitive Flexibility | | Processing Speed | | RT |
| | Stop-Change Error Rate (%) | Cognitive Flexibility | | Attention | | Error rate |

**Supplementary Table 2:** Characteristics of PPN-DBS and NBM-DBS studies

| Study name | Study design | Sample Size | Sex (male) | Age | Disease duration | Med | Stim | OFF | VLFS | LFS | LHFS | HFS | VF | EF | RT | A | WM | EM | VS | S |
|---|---|---|---|---|---|---|---|---|---|---|---|---|---|---|---|---|---|---|---|---|
| **PNN** | | | | | | | | | | | | | | | | | | | | |
| **Costa 2010** | Within-subject | 5 | 18 | 60.6 | 10.5 | Off | B | x | | x | | | | | | | x | | | |
| **Arnulf 2010** | Within-subject | 2 | - | 62.5 | 22.5 | Off | B | x | | x | x | | | | | | | | | x |
| **Alessandro, 2010 / Ceravolo 2011** | Within-subject | 6 | - | 62.8 | 11.8 | On | B | x | | x | | | x | x | | | x | x | | x |
| **Nosko 2014** | Within-subject | 9 | 7 | 64 | 22.3 | Off/On | B | | | x | x | | | | | | | | | |
| **Thevathasan 2014** | Within-subject | 11 | - | 64.5 | 11.6 | Off | U / B | | | x | x | | | | x | | | | | x |
| **Fischer 2015** | Within-subject | 8 | 3 | 66.4 | 8.9 | On | B | x | x | x | | | | | x | | | | | |
| **Strumpf 2016** | Within-subject | 5 | - | 62.8 | - | - | U / B | x | x | | x | | | | | | | | x | |
| **Skvortsova 2021** | Within-subject | 3 | 1 | 60.0 | - | Off | B | x | | x | | | | | | | x | | | |
| **NBM** | | | | | | | | | | | | | | | | | | | | |
| **Gratwicke 2017** | Within-subject | 6 | 6 | 65.2 | 12.7 | - | B | x | | x | | | x | | x | x | x | x | | |
| **Nombela 2019** | Case report | 1 | 1 F | 68 | 7 | On | B | x | | x | | | x | x | | | x | x | x | |
| **Sasikumar 2021** | Within-subject | 6 | 5 | 65.7 | 11.5 | On | B | x | | | x | x | x | | | x | | | | |

PPN Pedunculopontine nucleus; NBM Nucleus Basalis of Meynert; Age (mean years); Disease duration (mean years); Med Medication status, Stim Stimulation side Bilateral or Unilateral; VLFS, Very low-frequecy stimulation (5-10 Hz); LFS, Low-frequecy stimulation (10-25 Hz); LHFS, Low-high frequecy stimulation (60-80 Hz); HFS, High-frequency stimulation (≥130 Hz); VF Verbal Fluency; EF Executive Function; RT Reaction Time; A Attetion; WM Working Memoy; EM Episodic Memory; VS Visuospatial; S Sleep.
