## Supplementary Table for "Frequency-dependent cognitive effects of Deep Brain Stimulation in Parkinson’s Disease: A Systematic Review and Meta-Analysis"

**Supplementary Table 3:** Summary of Low vs High Frequency DBS studies on cognitive domains

| Autor, date | Type Study and n | Target DBS / Med condition | Cog Domain/Other domain | Frequency of stimulation | Main findings | Comments |
|---|---|---|---|---|---|---|
| **STN** | | | | | | |
| **Verbal Fluency** | | | | | | |
| **Wojtecki, September 2006** | Randomized, 12 PD | Bilateral STN<br>Med off | Verbal Fluency (phonemic, category and switching) | OFF, 10Hz, ≥130Hz, constant contact, amplitude and pulse (15 min washout) | Overall VF was significantly better at 10 Hz (48.3 words) compared with 130 Hz and showed a nonsignificant trend toward worsening at 130 Hz (42.3 words) compared with off (43.8 words). | Phonemic VF: 10 Hz 10.3 ±3.2 words vs 130 Hz 9.1 ±4.9 vs OFF 10.1 ±4.3;<br>Switching phonemic VF: 10 Hz 9.3 ±3.3 vs 130 Hz 7.6 ±3.8 vs OFF 7.8 ±3.8;<br>Category VF 10 Hz 16.7 ±3.9 vs 130 Hz 14.8 ±3.4 vs OFF 14.9 ±3.8<br>Switching category VF 10 Hz 12.1 ±3.0 vs 130 Hz 10.8 ±1.7 vs OFF 11.1 ±2.2 |
| **Stegemöller, October 2013** | Randomized, 17 PD (8 tremor and 9 non tremor dominant) | Bilateral STN<br>Med off | Verbal Fluency (phonemic and semantic VF)<br>Motor, gait and balance assessment | OFF, 60Hz, 130Hz, constant amplitude and pulse (10 min washout) | No differences between groups or stimulation conditions were found for verbal fluency (combined mean: phonemic VF 60Hz 17.0 ±9.1 vs 130Hz 17.2 ±9.6; semantic VF 60Hz 11.4 ±4.7 vs 130 Hz 10,4 ±4,5). | 130Hz significantly reduced UPDRS tremor score in the tremor dominant group; while no other motor scores significantly differed. No differences were found for gait and balance measures. |
| **Fagundes, October 2016** | Randomized, 20 PD | Bilateral STN<br>Med ON | Verbal Fluency (phonemic, semantic, action and unconstrained) | 60Hz, 130Hz (60 min washout) | Positive impact on phonemic and action VF. Phonemic VF, letter P: 60 Hz 16.5 ±8.1 vs 130 Hz 12.5 ±7.0; Phonemic VF, letter FAS: 60 Hz 26,8 ±15.0 vs 130 Hz 23.8 ±12.5, p=0.042. Action VF: 60 Hz 9.9 ±5.5 vs 130 Hz 7.6 ±8.7, p=0.004. | Semantic VF, animals: 60 Hz 13.7 ±5.4 vs 130 Hz 12.9 ±4.8, p=0.290. Unconstrained VF: 60 Hz 29.6 ±13.1 vs 130 Hz 27.6 ±11.6, p=0,292 Phonemic FAS was negatively correlated with age ($r$=−0.0473; p=0.041). Phonemic P was negatively correlated with UPDRS-II ($r$=−0.686; $p$=0.002). |
| **Grover, December 2018** | Randomized, 15 PD | Bilateral STN<br>Med off | Verbal Fluency (phonemic and semantic VF),<br>Speech intelligibility (SI), perceptual speech characteristics | 60Hz, 80Hz, 110Hz, 130Hz or 200Hz; adjusted voltage according TEED (20 min washout) | Phonemic VF cluster switching score improved with LFS (60Hz 26.1 ±9.8, 80Hz 22.3 ±7.6, 110Hz 21.9 ±8.7, 130Hz 19.1 ±9.7, 200Hz 17.4 ±8.2, p=0.005). Semantic VF tends to improve with LFS (60Hz 11.4 ±6.2, 80Hz 12.3 ±4.4, 110Hz 14.1 ±6.3, 130Hz 11.2 ±5.9, 200Hz 11.4 ±5.1, p=0.069).<br>SI and, in particular, articulation, respiration, phonation and prosody improved with LFS (all p < 0.05). | No significant difference on the number of words in the phonemic VF (60Hz 33.4 ±14.2, 80Hz 30.3 ±13.1, 110Hz 33.0 ±15.1, 130Hz 34.7 ±15.5, 200Hz 28.1 ±13.1, p=0.171) or in Semantic switching score VF (60Hz 4.5 ±3.5, 80Hz 6.3 ±3.9, 110Hz 8.0 ±6.0, 130Hz 5.3 ±3.9, 200Hz 6.1 ±3.8, p=0.146). There was a negative correlation between perceptual characteristics of speech and duration of chronic stimulation (all p < 0.05). |
| **Lee, April 2021** | Randomized, prospective, 9 PD | Left STN<br>Med NA | Verbal Fluency (letter, episodic category, nonepisodic category) | OFF, dorsal and ventral 130Hz, dorsal and ventral 4-8Hz (stimulation with peak theta recorded at baseline), 2.5v, 100 µs (5 min washout) | Left dorsal theta stimulation improves overall verbal fuency compared to no stimulation and dorsal or ventral gamma stimulation but not compared to ventral theta stimulation. Episodic verbal fuency was improved with dorsal theta stimulation compared to all other conditions. | No significant differences in non-episodic category or letter verbal fluency. |

| | | | | | | |
|---|---|---|---|---|---|---|
| **Busteed, August 2024** | Non-randomized, prospective, 38 PD | Bilateral STN Med ON | Verbal Fluency (phonemic, semantic, switching and action)<br><br>Parkinson's Disease—Cognitive Rating Scale (PD-CRS) and Frontal Systems Behavior Scale (FrSBe) | No DBS (n=14), LFS 60-90Hz (n=10), HFS 130-180Hz (n=14) | Animals fluency: LFS group (20.0 ±3.5 words) better than HFS group (15.9 ±4.5, p=0.031). Letter fluency: HFS group performed worse (23.5 ±6.9 words) than the noDBS (31.6 ±7.7, p=0.018), but not than LFS (27.1 ±8.1, p=0.485). LFS and noDBS performed similarly (p=0.321).<br>LFS group showed larger cluster sizes in the letter P (p=0.006) and semantic fluency fruits and vegetables conditions (p=0.019), resulting in better overall performance compared to HFS group (p=0.012). | Comparison of neuropsychological scores pre- and post-surgical intervention revealed significant declines in letter (p=0.005), semantic (p=0.005), and action fluencies (p=0.033) for the HFS group. The LFS group revealed no significant decline.<br><br>All groups performed similarly on the PD-CRS (p=0.437) and the FrSBe (p=0.746). PD-CRS total = noDBS 95.1 ±7.3, LFS 98.4 ±11.4, HFS 93.4 ±11.3; FrSBe total noDBS 64,9 ±19.5, LFS 72.1 ±20.7, HFS 65.1 ±14.5. |
| **Schoenwald, March 2025** | Randomized, 20 PD | Left STN Med off | Verbal Fluency (phonemic VF) | OFF, 6Hz circular (3 mA), 6Hz directional (2 mA), dorsal contact with directional leads, 60 µs (20 min washout) | Best directional stimulation improved VF performance signicantly compared with other conditions: best-dDBS versus oDBS (t(69.5)= 3.34, p=0.001), versus DBS-off (t(69.4)= 4.034, p=0.011), versus second-best-dDBS (t(69)= 3.385, p= 0.01), and versus worst-dDBS (t(69.3)= 5.939, p= 0.001). | Neither contact orientation nor TEDD had a signicant main effect on VF performance. Voxel-wise analysis revealed a non-statistically significant cluster centered on the dorsolateral border associative STN associated with better-than-average VF improvement. |
| **Imbalzano, April 2025** | Randomized, 12 PD (1 month analysis), 14 PD (1 hour analysis) | Bilateral STN Med ON | Verbal Fluency (phonemic, category episodic, category non-episodic and switching) | Standard dorsal 100-180Hz vs dual stim 6 Hz most ventral contact + dorsal 100-180Hz, 60 µs, 2-3 mA (1 hour and 1 month) | Non-episodic (mean 7.4 words ±2.7 versus 5.4 ±2,5, p=0.038) and episodic VF (12.25 ±4.7 versus 9.50 ±2,43, p=0.030) improved after 1 month. | At 1 hour, no signicant effects on any VF task.<br><br>At 1 month, no effect on phonemic and switching fuency. Motor and non-motor (MDS-UPDRS, BDI, QUIP-RS) outcomes were unaffected. |
| **Ricciardi, May 2025** | Randomized, 18 PD | Bilateral STN Med ON | Verbal Fluency (phonemic and action) | OFF, 10Hz, 130Hz, dual 130 + 10 Hz (130Hz on the contact used for chronic treatment + 10Hz at on the contact below it). Constant amplitude and pulse (30 min washout) | Phonemic VF improved more with the 130 + 10 Hz than 130 Hz condition (p= 0.006); no difference between 130 + 10 Hz and 10 Hz (p= 0.2) and between 130 Hz and 10 Hz (p= 0.6).<br>No changes on action VF. No signicant correlation between change score in VF under 130 Hz or under 130 + 10 Hz and age, disease duration, LEDD, MoCA, gait, fatigue or stress levels.<br>Positive correlation between VF performance across all frequencies and baseline VF (ρ= 0.40, P= 0.004) indicating that poorer VF at baseline is related to greater improvement with stimulation. | Composite Motor Score (items 3.4, 3.10, 3.17), gait velocity (meters walked/60 seconds), dual-task gait-VF, 2-back letter task, and visual analogue fatigue and stress scores. Worse motor score at 10 Hz than 130 Hz (p= 0.002) and 130 + 10 Hz (p= 0.01). No significant effect of the stimulation conditions on gait velocity (p= 0.2) or on the dual task cost (VF task) for gait velocity (p= 0.8). No effect on 2-back task or fatigue and stress levels.<br>No signicant correlations between the VTAs associated with motor, associative, and limbic regions of left, right, and both hemispheres and VF scores. |
| **Executive functions** | | | | | | |
| **Scangos, October 2017** | Randomized, 15 PD | Bilateral STN Med NA | Executive function (computerized Stroop task); NPI, BDI | OFF, 5Hz, 130Hz, deepest ventral contact, pulse 90-120 µs, amplitude mean 1.34v ±0.65 (15 min | 5 Hz improved incongruent trial accuracy vs OFF (rmANOVA: frequency effect F(2,18) = 5.09, p=0.018). 130 Hz had no effect on accuracy (p=0.937). | Improvements in Stroop incongruent accuracy and accuracy interference did not significantly habituate after two days of continuous 5 Hz stimulation (rmANOVA, accuracy: F(1,9)=1.90, p=0.201; interference: |

| | | | | | | |
|---|---|---|---|---|---|---|
| | | | | washout); then 48 hours with 5Hz stimulation | 5 Hz signicantly improved (p=0.002) and 130 Hz trended toward worsening (p=0.055) stroop accuracy interference compared to no stimulation.<br>The post-confict adjustment score ((iC - cC)+(cI - iI), where iC=congruent trial preceded by an incongruent trial, cI=incongruent trial preceded by a congruent trial, etc.) examine the adaptation of behavior across trials. 130 Hz signicantly worsened conflict adaptation (rmANOVA: frequency effect (F(2,32) = 10.15, p<0.001). No significant effect with 5 Hz (p=0.424). | F(1,9)=2.23, P=0.170). Changes in task accuracy were not due to improvements in motor agility.<br><br>After 48 hours with 5 Hz, there was a significant improvement in BDI (p=0.043) and no patient showed motor worsening (UPDRS-III). |
| **Varriale, August 2018** | Randomized, 19 PD (16 with FOG - FOG-Q item 3 2.3 ±1.3) vs 20 HC | Bilateral STN Med ON | Modified Stroop paradigma (single gait initiation and cognitive interference task - initiating gait "Go" or not "No GO" depending on the colour or the shape of a visual cue)<br>Number of errors (incorrect initiation of gait for a ''No Go" trial or an absence of gait initiation for a ''Go" trial) | 80Hz, 130Hz, same contacts with adjusted TEED (30 min washout), compared with ON/OFF medication before surgery | Simple gait task (n=19 PD): Significant gait initiation improvement with dopaminergic treatment, 80 Hz and 130Hz. Compared with 130 Hz, 80 Hz showed a significantly longer step length and shorter double stance duration.<br><br>Cognitive interference task (n=9 PD): with 130 Hz all gait initiation parameters were significantly degraded, the anticipatory postural adjusments (APAs) and double-stance phase durations were significantly higher, and the antero-posterior and medio-lateral CoP displacements and step length were significantly lower with 130 Hz versus 80 Hz. Cognitive interference task induced no major changes with 80 Hz.<br><br>Number of errors: No significant difference in error rates was found across all groups. | The subject's reaction time was significantly higher during cognitive interference compared to the simple gait task (F = 232.73, p<0.001). No significant effect of group-treatment condition.<br><br>During the cognitive interference, compared to the simple gait task, with 130 Hz, the APAs duration significantly increased (F = 7.38, p<0.001), and both the antero-posterior (F = 12.09, p<0.001) and mediolateral (F = 18.36, p<0.001) centre of foot pressure (CoP) displacements significantly decreased.<br>During the cognitive interference, compared to the simple gait task, with 80 Hz and 130 Hz, step length was significantly lower (F = 16.12, p<0.001) and double stance duration was significantly longer (F = 5.58, p=0.018). No major changes on the APAs duration with 80 Hz, compared to simple gait task. |
| **Lam, January 2021** | Randomized, 12 PD | Bilateral STN Med ON | Executive function (verbal fuency - letter, episodic category, nonepisodic category, and category switching, color-word interference and random number generation task) | OFF, 10Hz, 130-135Hz (5 min washout) | Episodic category VF scores were 16% higher with 10 Hz vs 130-135 Hz (OFF 17,2 ±5,4 vs 10 Hz 18,0 ±5,1 vs 130−135 Hz 15,5 ±5,1). No difference compared to Off. There were no signicant diferences between conditions for letter, nonepisodic category or switching scores. | Color-word interference: no signicant differences across conditions in color processing, word processing, color-word interference or color-word switching.<br>No signicant differences in any indices of random number generation including Evan's RNG. |
| **Waldthaler, January 2023** | Randomized, 14 PD | Bilateral STN Med off | Response inhibition (antisaccade error rate)<br>EEG recording | OFF, 60Hz, 130Hz, constant contact, amplitude and pulse (10 min washout) | 60 Hz induced a slight but significant reduction of directional errors compared with OFF and 130 Hz. % of correct trials: OFF 51.4%, 60 Hz 59.1%, 130 Hz 54.4%. 60 Hz increased the probability of a correct antisaccade compared with 130 Hz (OR | Both frequencies decreased the onset latency of correct antisaccades, while increasing the latency of directional errors (not significant diferences). |

| | | | | | | |
|---|---|---|---|---|---|---|
| | | | | | =1.44, 95%CI [1.23, 1.69], p<0.001) and OFF (OR=1.54, 95%CI [1.31, 1.81], p<0.001).<br><br>60 Hz was associated with an increase in preparatory theta power over a midfrontal region compared with OFF but not 130 Hz. | Both frequencies were associated with a stronger midfrontal beta desynchronization during the mental preparation for correct antisaccades compared with OFF.<br><br>Higher midfrontal theta activity was associated with longer antisaccade latency in OFF; that was reversed by 130 Hz. |
| **Zacharia, March 2023** | Randomized, 21 PD vs 16 HC | Bilateral STN Med off | Prosaccades (latency and gain) and antisaccades (latency of correct and incorrect antisaccades, error rate and gain of the correct antisaccades);<br>Stroop task | 80Hz, 130Hz, adjusted voltage according TEED (20 hours washout) | With 80 Hz, patients perform less well in an antisaccade task.<br><br>Antisaccadic error rate was higher in PD patients (p= 0.0113), and more on 80 Hz compared to 130 Hz (p= 0.001).<br><br>The number of errors during the Stroop inhibition/switching subtest remained unchanged. | Prosaccades - No signicant differences in neither latency (p= 0.1191), nor gain (p= 0.8396).<br><br>Stroop task - Inhibition task (s) 130 Hz 68.8 ±23.2 vs 80 Hz 71.4 ±21.8; Inhibition task (number of errors) 130 Hz 1.9 ±3.8 vs 80 Hz 1.9 ±3.9; Switching task (s) 130 Hz 94.2 ±41.0 vs 80 Hz 87.3 ±39.3. |
| **Qin, August 2023** | Randomized, 29 PD PIGD patients | Bilateral STN Med Off | Executive functions (Stroop Color-Word Test (SCWT), Verbal fluency, Symbol Digital Switch, Digital Span, and Judgment of Line Orientation test) | OFF, 5Hz, 130Hz; constant pulse and amplitude, contact closest to motor STN selected with Lead-DBS (22 hours washout) | Compared to 130 Hz, 5 Hz significantly decreased the completion time of SCWT-C (5 Hz 82.2 ±24.8, 130 Hz 91.7 ±27.3, p<0.001) and Stroop interference effect (5 Hz 43.3 ±18.3; 130 Hz 54.2 ±21.9, p<0.001). Compared to OFF, 5 Hz tends towards decreased Stroop interference effect (5Hz 43.3 ±18.3, OFF 48.7 ±26.2, p=0.08). | There was no difference among stimulation conditions in accuracy of SCWT-A, SCWT-B, and SCWT-C and time of SCWT-A and SCWT-B.<br><br>No significant differences among stimulation states were found for other cognitive tests. |
| **Cole, May 2025** | Randomized, 15 PD | Bilateral STN Med ON | Decision threshold (Simon task accuracy and reaction time (RT), congruent versus incongruent trials) | OFF, 4Hz, 120–160Hz, constant contact, amplitude and pulse (45 min washout) | Accuracy was overall high (84.6 %). No main effect of DBS condition.<br><br>RT = effect of DBS condition (F(2,70) =8.4, p<0.001) but no interaction between DBS condition and trial type on RT (F(2,70)=0.1, p=0.89). RTs were significantly faster under ~130Hz vs 4 Hz (4 Hz > ~130 Hz; t=4.1, p=0.003).<br><br>Decision threshold (using computational diffusion decision modeling analyses) was significantly higher under 4 Hz vs ~130 Hz (p=0.02). | No main effect of DBS condition (F(2,70)=0.1, p=0.89) or interaction of trial type with DBS condition on accuracy (F(2,70)=0.07, p=0.93).<br><br>RTs were significantly slower on incongruent versus congruent trials (F(1,70) =124.2, p<0.001.<br><br>MDS-UPDRS III better at ~130 Hz (20.9 ±13.1) versus 4 Hz (27.1 ±15.2, p=0.004) versus OFF 35.1 ±19.4, p=0.00007). |
| **Xie, July 2025** | Randomized, 18 PD with FOG (FOG-Q 16.17 ±5.40; FOG-Q item 3 3.06 ±0.87) | Bilateral STN Med off | Conflit resolution (conventional Flanker task), fMRI | OFF, 5Hz, 130Hz, constant amplitude and pulse (60 min washout) | 5 Hz significantly reduced the conflict effect (reaction time cost and error cost) in PD-FOG patients performing the CFT.<br><br>5 Hz significantly activated frontal and parietal regions and deep cerebelar nuclei, | 130 Hz had no behavioral effect; but, only 130 Hz significantly improved MDS-UPDRS III.<br><br>130 Hz significantly activated bilateral thalamus, GPi, and deep cerebellar nuclei, and decrease activation in frontal and |

| | | | | | | |
|---|---|---|---|---|---|---|
| | | | | | including the bilateral medial SFG, bilateral MFG, left triangular IFG, orbitofrontal cortex, bilateral angular gyrus, and cerebellar posterior lobe. | temporal regions (medial SFG extending to pre-SMA/dACC, bilateral IFG, and MTG). |
| **Kricheldorff, December 2025** | Randomized, 17 PD | Bilateral STN Med On | Conflit resolution 4 paradigmas: response selection task (response execution), flanker task (conflict monitoring), Go-NoGo task (automatic inhibition) and stop-change task (controlled inhibition). | OFF, 20Hz, 130Hz, constant amplitude and pulse (20 min washout) | 20 Hz improves automatic inhibition, marked by increased accuracy<br><br>Flanker task: slower at 20 Hz vs 130 Hz [m = 77.0 ms, 95%CI (58.3 ms, 96.0 ms)], but not OFF compared to 20 Hz; fewer errors at 20 Hz vs 130 Hz [m = −3.7%, 95% CI (−5.4%, −1.9%)] and vs OFF [m = −3.4%, 95% CI (−5.2%, −1.6%)].<br>Go-NoGo task: slower at 20 Hz vs 130 Hz on uncertain Go trials [m = 105.2 ms, 95%CI (46.7 ms, 164.2 ms)] and vs OFF [m = 136.8 ms, 95%CI (75.6 ms, 198.0 ms)]; fewer errors on uncertain Go trials at 20 Hz vs 130 Hz [m = −6.8%, 95% CI (−10.1%, −3.6%)] and vs OFF [m = −6.5%, 95% CI (−9.8%, −3.3%)]; fewer errors on NoGo trials at 20 Hz vs 130 Hz [m = −10.3%, 95% CI (−14.5%, −6.0%)] and vs OFF [m = −10.6%, 95% CI (−15.0%, −6.3%)]. | Response selection task: faster at 130 Hz vs 20 Hz [m = −33.6 ms, 95%CI (−50.2, −16.7)], but not OFF compared to 20 Hz; fewer errors at 20 Hz vs 130 Hz [m = −6.0%, 95%CI (−8.1%, −3.9%)], but not OFF compared to 20 Hz.<br><br>Stop-change task: no differences |
| **Memory** | | | | | | |
| **Salehi, January 2024** | Randomized, 20 PD | Bilateral STN Med off | Working memory (modified, computerized Sternberg task - participants were instructed to maintain a sequence of five digits and accurately reproduce it in the correct order after arithmetic distractor task, block of 24 trials) | OFF, 6Hz, 15Hz, 70Hz, 130Hz, constant contact, amplitude and pulse (10 min washout) | 6 Hz enhances WM performance.<br>6 Hz stimulation increased WM performance compared to 15 Hz (p<0.01) and 70 Hz (p<0.01), with 130 Hz approaching significance (p=0.054).<br><br>Patients with lower baseline performance showed greater improvement as measured by WM off stimulation (interaction effect frequency*off WM: F1,1853=2.9, p=0.03) and the UPDRS I (F1,19=6.5, p=0.02). | Negative pearson correlation between WM off stimulation and WM changes with 6 Hz (R=-0.71, p<0.001). Positive correlation between WM off and MoCA score (R=0.67, p<0.01).<br><br>Based on normative connectome data, effects of 6 Hz on WM performance correlated with connectivity of the VTA to the right DLPFC centered in the right middle frontal gyrus.<br><br>No motor performance diferences between Off, 6 Hz and 15 Hz but worsen motor function compared to 70 Hz and 130 Hz. |
| **Herz, November 2024** | Randomized, 18 PD (11 tested 2 to 4 days after DBS surgery, 7 chronically | Bilateral STN Med (NA) | Sleep and Memory retention (declarative memory task with learning task conducted in the evening; delayed recall in the morning on the next day)<br>EEG recording | 4Hz (n=9), 130Hz (n=9) during first stage-2 NREM sleep, five 5-min blocks, 1-min OFF interval; rest of the night and memory task 130 Hz; | The number of correctly recalled words from the word pairs was taken as a measure of delayed recall.<br><br>Patients who received 4 Hz DBS showed an increase in recalled words (p=0.027) and higher prefrontal LFO (2–8 Hz) activity in | In the 4 Hz group, a stronger expression of prefrontal LFO was correlated to a better cognitive performance (Spearman correlation ρ=0.711, p=0.037). |

| | | | | | | |
|---|---|---|---|---|---|---|
| | treated with DBS) | | | constant contact, amplitude and pulse | their EEG (four 1-minute OFF-stimulation recordings) compared to 130 Hz (p= 0.039). | |
| **Other cognitive domain and Sleep** | | | | | | |
| **Wojtecki, September 2011** | Randomized, 12 PD vs 12 HC | Bilateral STN Med off | Time Processing (interval timing - four paradigms: time reproduction task and time production task for intervals of 5 and 15 s; time discrimination task with intervals ranging from 800 to 1600ms and tapping task with inter-tap intervals of 800 ms) | OFF, 10Hz, 130-150Hz, constant contact, amplitude and pulse (15 min washout) | PD and HC over-reproduced the 5 s interval and under-reproduced the 15 s interval in both time reproduction and production tasks.<br><br>10 Hz significantly worsened interval timing at the 15 s interval. HC and PD with ≥130Hz showed lowest impairment of time processing.<br><br>Performance in milliseconds timing, as measured by the time discrimination and tapping tasks, did not differ between PD and HC or between stimulation conditions.<br><br>Significantly worse UPDRS motor score with OFF 49±3 and 10 Hz 45±2, vs ≥130 Hz 26±3. | Reproduction task, 10 Hz significantly enhanced the 15 s underreproduction: s mean±SEM 10 Hz 10.4±0.9; vs OFF 12.5±0.8, p<0.05; vs ≥130Hz 13.5±0.6, p<0.05; vs HC 14.8±0.2, p<0.001. Production task, 10 Hz significantly enhanced the 15 s underreproduction: 10 Hz 10.1±0.5; vs OFF 11.1±0.8, p<0.05; vs ≥130Hz 13.2±0.9; p<0.05; vs HC 13.9±0.7, p<0.01.<br><br>Time discrimination task: number of correct judgements intervals did not significantly differ between HC (41±2) and PD (10 Hz 38±1; OFF 35±2; ≥130 Hz 35±2). The mean intertap intervals (in ms) did not differ significantly between HC (773) and PD (10 Hz 744; OFF 805; ≥130 Hz 819). |
| **Amara, June 2016** | Randomized, 20 PD | Unilateral STN (18), Bilateral STN (2)<br>Med ON | Sleep (change in sleep efficiency based on polysomnography) and Reaction Time (Psychomotor vigilance task - PVT) | OFF, 60Hz, ≥130Hz, bipolar stimulation to reduce PSG artifacts (3 days washout) | No difference in sleep efficiency between nights with ≥130 Hz ((median [IQR] 82.1% [72.6–90.1]), 60 Hz (81.2% [56.2–88.8]), or DBS off (82.8% [75.7–87.4]; P= 0.241). Performance on the PVT was not different on the morning following the night with ≥130Hz (RT 3.2 ±0.6) compared to 60 Hz (3.2 ±0.5). | No difference in sleep stage percent, arousals, limb movements, subjective sleep quality, or objective vigilance measures.<br><br>No differences in the UPDRS III, the Stand-Walk-Sit test, or the timed hand/arm movement between two points. |
| **Kelley, November 2018** | Consecutive 10 PD undergoing STN-DBS with intraoperative recordings and EEG<br>+<br>Randomized, 10 PD with DBS and EEG | Bilateral STN Med Off (intraoperative recordings), Med ON (DBS manipulation) | Time Processing - Interval timing task (participants were asked to estimate a time period of 3 or 12 sec; trials started with an instructional cue (a recorded voice stating 'three' or 'twelve') followed 1 s later by an 8-kHz tone ('starting cue') indicating the start of the interval) | (1) Intraoperative recording<br><br>(2) Off, 4Hz, 120-150Hz, constant contact, amplitude and pulse (30 min washout) | (1) During cognitive task, 4 Hz medial prefrontal cortex signals exert top-down control of the STN. Time-frequency analysis revealed a cue-triggered delta/theta 1–4 Hz activity from midfrontal EEG leads; cue-modulated low-frequency activity in the STN and strong 4 Hz coherence between mid-frontal EEG and STN.<br><br>(2) 4 Hz stimulation improved cognitive performance in the interval-timing task. | (1) Patients responded at 2.9 ±0.3 s on 3 s (FI3) trials, and at 8.4 ±1.2 s on 12 s (FI12) trials (p<0.0004).<br><br>(2) 4 Hz STN-DBS increased cue-related delta activity (1–4 Hz) at midfrontal EEG. 4 Hz STN-DBS improved performance on FI12 trials, bringing responses closer to 12 s and to the response times of healthy controls. 120-150 Hz did not change interval timing performance compared to DBS off. |
| **Romangnolo, January 2021** | Randomized, 12 PD | Bilateral STN, Med off | Event-related potentials (ERPs) recordings using a standard oddball auditory paradigm (subjects silently count rate tones differing from others in pitch). Peak amplitude and latency of the | OFF, 60 Hz, 80 Hz, 130 Hz, adjusted amplitude according TEED (20 min washout) | P300 latency over Cz and Pz electrodes significantly increased at 130 Hz compared to OFF. P300 latency was also significantly increased, though to a lesser degree, over Pz electrode with stimulation at 80 Hz. No significant P300 latency modifications were detected at 60 Hz compared to OFF. P300 | The accuracy of "target" stimuli detection did not differ in the four experimental conditions.<br><br>LFS may have a milder impact on neural circuits involved in the generation of P300, a possible neurophysiological marker of cognitive decline in PD. |

| | | | | | | |
|---|---|---|---|---|---|---|
| | | | P300 components at midline positions (Fz, Cz, Pz). | | amplitude did not change significantly for any of the stimulation conditions tested. | |
| **Memon, September 2023** | 15 PD (*post hoc* analysis Amara, 2016) | Unilateral STN (14), Bilateral STN (1) | Sleep (change in sleep spindle density based on polysomnography-derived quantitative electroencephalography (qEEG); other sleep qEEG features) | OFF, 60Hz, ≥130Hz, constant amplitude, bipolar stimulation to reduce PSG artifacts (3 days washout) | Spindle density during N2 sleep was significantly higher in the ≥130 Hz condition compared to 60 Hz.<br>Slow wave amplitude during N2-3 REM sleep was significantly higher with 60 Hz compared to DBS OFF and ≥130 Hz conditions. | No significant differences were observed in the other sleep qEEG features during sleep at different DBS conditions. |
| **McAuley, March 2025** | Case Report, 1 PD | Bilateral STN Med ON | Kinarm tasks: visually-guided reaching (motor), reverse visually-guided reaching (cognitive-motor), spatial span (memory), and object hit (bimanual coordination) | 104 Hz, 149 Hz, 179 Hz, amplitude adjustements, constant contact C1/C9 and pulse 60 µs (60 min washout) | The reverse visually-guided reaching (cognitive-motor function) task revealed significant impairments under 104Hz.<br><br>No difference in the spatial span task (measure of short-term memory). | Gait assessment showed a substantial gait difficulty under 104Hz (more time using support bars) and greater arm swing asymmetry and greater variation in arm swing under 149Hz.<br>Speech impairment ranged from mild (104Hz) to moderate-severe (149Hz). |
| **Mood and Emotional Processing** | | | | | | |
| **Mandali, December 2020** | Randomized, 24 PD (11 PD with 10Hz and 130Hz, 12 PD with 10Hz) | Right STN Med NA | Emotional processing (International Affective Picture System, – rating 0 to 100 – very negative to very positive (valence) or very exciting to not exciting (arousal)). | OFF, time-locked acute stimulation for 1 second at 10 Hz or 130 Hz, constant pulse (90 µs), adjusted voltage according TEED (24 hours washout); LFP and EEG recorded simultaneously. | Great alpha desynchronization in both negative and positive affect relative to neutral timuli.<br><br>10 Hz increased the subjective pleasantness of negative imagery - 10 Hz enhanced subjective positive valence ratings of negative images relative to OFF stimulation with no difference observed for 130 Hz. | 130 Hz decreased subjective arousal ratings (lower excitability) relative to OFF stimulation but no difference compared 10 Hz.<br><br>Higher depression scores were associated with a positive bias at 10 Hz but not 130 Hz stimulation. Greater positive ratings at 10 Hz were associated with more medial contacts, with a trend to more ventral contacts. |
| **Muhammad, July 2023** | Randomized, 24 PD (post hoc analysis Mandali, 2020) | Right STN Med ON | Emotional processing (International Affective Picture System - comparing negative and neutral stimuli); LFP and EEG recordings | OFF, time-locked acute stimulation for 1 second at 10 Hz or 130 Hz (24 hours washout) | 130 Hz showed a decrease in alpha power to negative vs. neutral images. This alpha power decrease wasn't evident in the negative 10 Hz condition. 10 Hz has the capacity to facilitate the synchronization of alpha and enhance alfa power. | 10 Hz condition shows a increase in beta power along with a positive correlation between beta power across the 10 Hz and OFF conditions suggesting physiological and cognitive generalization effects. |
| **Wang, August 2023** | Randomized, 20 PD | Bilateral STN Med ON | Emotional processing (self-report valence and arousal with the International Affective Picture System – rating 0 to 100 – very negative to very positive (valence) or not exciting to very exciting (arousal)) | OFF, 10Hz, 130Hz, monopolar, lowest contact within the STN, constant voltage, 90 µs (10 min washout) | Significant reduced arousal under 130 Hz vs OFF and 10 Hz (for positive, neutral and negative images).<br><br>In depressed patients (BDI>12points, 10 patients), 10 Hz increased the arousal ratings compared to OFF (p=0.028) and 130 Hz (p=0.007).<br><br>In non apathetic patients (AES>7, 8 patients), 10 Hz induced a positive shift in valence rating of negative images (p=0.011); in apathetic patients, 130 Hz led to more | MDS UPDRS-III = OFF 39 ±16; 10 Hz 39 ±15; 130 Hz 28 ±12; No correlation between motor score and arousal ratings except for negative arousal at 10 Hz (Pearson's r=0.485, p=0.03).<br>A lower arousal ratings at 130 Hz (Pearson's r=− 0.456, p=0.043) and at 10 Hz (Pearson's r=− 0.522, p=0.018) were correlated with more anterior contacts.<br>After controlling for neutral images, a more ventral STN stimulation was correlated to a greater positive valence ratings bias at 10 Hz (Negative images: Pearson's r =−0.520, |

| | | | | | | |
|---|---|---|---|---|---|---|
| | | | | | positive valence rating of positive images compared to OFF (p=0.039) and of negative images compared to 10 Hz (p=0.034). | p=0.019) and at 130 Hz (Positive images: r =−0.479, p= 0.033). |
| **Scangos, December 2016** | Case Report, 1 PD | Bilateral STN Med NA | Mood (Clinical judgment, BDI, NPI) | 5 Hz, 130 Hz, 1.1 v (48 hours) | With 130 Hz, patient showed manic symptoms (BDI-II 13, NPI 13, subscore hallucinations 3, agitation/aggression 1, elation 3 and apathy/indifference 6).<br>After 5 Hz, symptoms have resolved. | Baseline BDI-II 8, NPI 5, subscore elation 2 and irritability 3. |
| **Imbalzano, June 2020** | Case report, 1 PD | Bilateral STN Med NA | Mood (Clinical judgment, BDI, HAD) | 60Hz, 80Hz, 130Hz, stepwise increase in voltage | Left STN stimulation resulted in acute mood complications (acute sadness with cry) at low amplitude. Low frequency resulted in higher voltage thresholds for occurrence of mood side effects (the threshold was higher than expected by calculating TEED). | With monopolar stimulation of contact 1 at the threshold values for 130, 80, and 60 Hz, higher scores indicate a depressive/anxiety state (BDI-Sort Form 11/39, HADS anxiety 6/21, HADS depression 6/21). |
| **PPN** | | | | | | |
| **Costa, Jan 2010** | Randomized, 5 PD (PPN) vs 5 PD (no DBS) vs 8 HC | Bilateral STN+PPN Med off | Working memory (n-back paradigm - series of visual patterns or words, n-1 steps) | 25Hz PPN vs OFF PPN (120 min washout); STN off | PPN group: the average accuracy did not change between Off and On condition either in the visual-object (Off 13.7 ±6.5, On 14.2 ±6.4) or the verbal task (Off 13.4 ±6.9, On 13.9 ±5.9). But, the average response times were significantly faster with On vs Off condition in both visual-object (On 783 ±355, Off 983 ±367) and verbal task (On 910 ±311, Off 1171 ±422). PPN stimulation facilitates the information processing speed during WM. | HC: off condition, PD patients tended to achieve slower response times (p=0.07). No difference for the accuracy scores on both n-back task. On condition, no significant difference on the accuracy and the response times.<br><br>No DBS: no significant difference on the accuracy and the response times between DBS and no DBS group. |
| **Arnulf, April 2010** | Randomized, 2 PD (with severe FOG) | Bilateral PPN Med off | Sleep (daytime videopolysomnography) | OFF, left, right, and bilateral, 10-25Hz or 80Hz randomly applied for at least 5 minutes with 3-minute washout periods | Left, right, and bilateral 80Hz consistently caused a marked feeling of sleepiness and induced sleep in both patients.<br>Electrophysiological sleep occurred within 0.6 to 2.5 min in Patient 1 and within 1 to 8 min in Patient 2. | When receiving uni- or bilateral 10-25 Hz, the patients were fully alert and spontaneously active.<br>In Patient 1, the abrupt cessation of the low-frequency PPN stimulation (tested 5 times) consistently induced REM sleep within 0.6 to 1.7 minutes. |
| **Alessandro, September 2010 / Ceravolo September 2011** | Randomized, 6 PD with severe axial signs | Bilateral STN+PPN Med OFF and ON | Cognitive battery (California Verbal Learning test, Long Delay Free Recall and Digit Span test; Trail Making test (TMT) and FAS verbal fluency; object naming test)<br>Epworth Sleepiness Scale (ESS), Parkinson's Disease Sleep Scale (PDSS), Pittsburgh Sleep Quality Index (PSQI)<br>PET | 25Hz PPN vs OFF PPN vs PPN cycling night ON (1-2 weeks each); constant contact, amplitude and pulse; STN Off | Compared to OFF, 25 Hz PPN significantly improved delayed recall (7 ±1.7 to 8.8 ±0.8 in ON, p < 0.05) and executive functions - TMT B-A 180 ±102.3 vs. 124 ±75.3 in ON, p < 0.05) and phonemic verbal fluency FAS (26.3 ±5.3 to 31.7 ±5.2 in ON, p < 0.001).<br><br>PPN-cyclic improveded nocturnal motor restlessness, psychosis and daytime sleepiness. ESS confirmed the reduced daytime sleepiness under both PPN-ON. | PSG (2 patients): 25 Hz PPN promoted a increase from <80% to >90% sleep efficiency, a mild reduction of Stage 1, a increase of Stage 2 and a decrease of awakenings.<br>Compared to OFF, 25 Hz PPN promoted a bilateral increased activity in prefrontal areas (p < 0.001) including frontal inferior gyrus, dorsolateral prefrontal cortex, orbitofrontal cortex, anteriorcingulate, superior frontal gyrus, parietal inferior lobule and supramarginal gyrus; a significant (p < 0.001) increase activity in the left ventral striatum, left subgyral, right insula and right superior |

| | | | | | | |
|---|---|---|---|---|---|---|
| | | | | | | temporal gyrus. PPN stimulation decreased glucose utilization in the left cerebellar anterior lobe (culmen) and right cerebellar posterior lobe (declive) (p < 0.0001). |
| **Thevathasan, November 2010** | Randomized, PD vs HC | Bilateral PPN Med off | Reaction Time (simple, choice and digit vigilance tasks), Speed of Reaction (sum of mean task RT) and Accuracy (omissions and percentage of correct responses) | OFF, 5Hz, 10Hz, 20-35Hz; 2.0-3.7 v and 60 µs (30 min washout) | Only the 20-35 Hz, significantly improved Speed of Reaction compared with off (Z=-2.756, p=0.02).<br>This improvement was significant only for the simple reaction time (not choice or digit vigilance task). | Compared with HC, in OFF condition, PD had significant deficits in Speed (mean±SEM PD 1691.8±189.7 vs HC 1193.1±8.0, p<0.001) and in all 3 tasks reaction times. Accuracy was not different from HC (mean±SEM PD 82.2±5.2 vs HC 90.6±0.1, p=0.56) and did not improve with stimulation. |
| **Nosko, September 2014** | Randomized, 9 PD with FOG (item 14 UPDRS 3 ±1) | Bilateral PPN (7 patients with STN-DBS) Med off and ON | Daytime sleepiness (Epworth Sleepiness Scale) and gait assessment | 10-25Hz, 60-80Hz; constant parameters, med ON/OFF (24 hours washout); STN with chronic parameters | Compared with 60–80 Hz, 10–25 Hz PPNa stimulation led to decreased akinesia, gait dificulties and daytime sleepiness in 7/9 patients. One patient aggravated under 10–25 Hz compared with 60–80 Hz.<br><br>Daytime sleepiness was greater under 60-80 Hz (Epworth Sleepiness Scale LFS 5±4, HFS 7±4). | UPDRS III Off med/Low 24.0 ±13,1, Off/High 29.1 ±13.0, On med/Low 19.6 ±10.8, On/High 22.5 ±10.4.<br>Non-significant difference in FOG duration, although 7/9 patients had less FOG under 10-25 Hz (FOG %duration Off med/Low 20.3±20.7, Off/High 19.2±20.5, On med/Low 14.0±16.6 On/High 16.0±15.4). Two patients could not perform gait test under 60-80 Hz. |
| **Fischer, July 2015** | Randomized, 8 PD with FOG | Bilateral PPN Med ON | 3 standard reaction time (RT) tasks - (a) the alertness task (unwarned and warned condition), (b) the go/no-go task, (c) the divided-attention task | OFF, 8Hz, 20Hz, 60Hz, 130Hz; constant contact, amplitude and pulse (12-24 hours washout) | Compared with OFF, RTs in the unwarned condition of the alertness test were significantly faster at 8 Hz, p=0.02 and 20 Hz, p=0.02. Stimulation did not affect patients' performance in more complex attentional tasks.<br><br>In 3 patients with STN-DBS, comparison off/on STN costimulation showed that patients with on STN became faster at 8Hz and 20 Hz PPN stimulation compared with off PPN. | Unwarned condition: there was a statistical trend for an improvement at 60 Hz (p=0.052). No difference between RT under 130 Hz and OFF.<br>Warned condition: there was a statistical trend for reduced RT under 20 Hz compared with OFF, p=0.07. No other relevant changes.<br><br>No differences in the performance of the go/no-go and the divided attention across conditions. |
| **Strumpf, May 2016** | 4 PD with gait impairment | Bilateral PPN (4), Right PPN (1), + STN (3) Med NA | Visual discrimination (Luminance contrast thresholds to discriminate an orientation-target (Gabor-grating)) | OFF, 8-10Hz, 60Hz, 60 µs, amplitude mean 1,8v ±0,57 (1-4 days washout) STN ON | Threshold increase for the LFS relative to OFF and HFS conditions - LFS showed an average contrast-threshold level (31.5%) that was roughly 10% higher than OFF (23%, p=0.08) and HFS condition (19.7%, p=0.043). | |
| **Skvortsova, March 2021** | Prospective, 3 PD | Bilateral PPN Med off | Reward learning task (choosing between two cues either left or right, and learning the probabilistic associations with monetary reward from the observation of choice outcomes). | OFF, 20-40 Hz | PPN local field potentials (LFPs), recorded while patients performed an instrumental learning task, showed a specific response to reward outcomes in a low-frequency (alpha-beta) band. | PPN DBS selectively improved learning from rewards but not from punishments. |
| **NBM** | | | | | | |

| | | | | | | |
|---|---|---|---|---|---|---|
| **Gratwicke, December 2017** | RCT,<br>6 PD with cognitive decline (MMSE 21-26) | Bilateral NBM<br>Med NA | Cognitive battery (California Verbal Learning Test-II, Wechsler Adult Intelligence Scale-III Digit span, Verbal fluency, Posner covert attention test and simple and choice reaction times); NPI; fMRI | OFF, 20Hz (crossover - 6 weeks ON, 6 weeks OFF, 2 weeks washout) | No improvements were observed in the primary cognitive outcomes or in results of resting state fMRI.<br><br>Neuropsychiatric Inventory improved with NBM DBS (8.5 points [range 4-26]) compared with OFF (12 [8-38]; p = 0.03) and the preoperative baseline (13 [5-25]; p = 0.69). | Both surgery and stimulation were well tolerated in this vulnerable patient group. At baseline, 3 patients reported daily visual hallucinations; 2 of these patients had near-complete cessation of hallucinations with NBM.<br><br>Three patients improved MDS-UPDRS part IV. |
| **Nombela, January 2019** | Case report, 1 PD | Bilateral NBM-GPi<br>Med ON | Cognitive battery (MMSE, Cognitive mini-evaluation, Digit Span WAIS-IV, TMT, Stroop Color-Word Test, Hopkins Memory, MDRS-II, Rey-Osterrieth Complex Figure, Verbal Fluency, Boston Nomination and Line Orientation Test) | GPi bipolar (left 4+5- 4.7mA, 60µs and right 12+13-, 4.0mA, 90µs, 130Hz) with NBM monopolar (left 2- and right 9-, 2.0mA, 60µs, 20Hz); NBM off 2 months vs NBM on 3 months | After 2 months with GPi stimulation, motor score improved but cognitive tests worsened. After 3 months with NBM+GPi, motor improvement remains similar but with some cognitive improvements (non-verbal memory, Rey-Osterrieth Figure - copy time (s) baseline 612, GPi 420, NBM+GPi 370; copy accuracy baseline 25.5, GPi 5, NBM+GPi 27; delay memory accuracy baseline 0, GPi 2, NBM+GPi 9; and VF - Letter P baseline 4, GPi 2, NBM+GPi 5; Letter M baseline 6, GPi 1, NBM+GPi 2). | Other cognitive tests did not improve.<br>MMSE (baseline 27, GPi 25, NBM+GPi 23); MDRS-II (baseline 134, GPi 110, NBM+GPi 111); conceptualization MDRS-II (baseline 39, GPi 33, NBM+GPi 31); category VF (baseline 12, GPi 11, NBM+GPi 8); Judgment Line Orientation (baseline 26, GPi 13, NBM+GPi 12); Direct Digit Span (baseline 6, GPi 5, NBM+GPi 6); Reverse Digit Span (baseline 3, GPi 3, NBM+GPi 2); Strop interference index (baseline 4.46, GPi NA, NBM+GPi 1.15) |
| **Sasikumar, October 2021** | RCT,<br>6 PD with mild cognitive impairment | Bilateral NBM-GPi<br>Med On | Sustained attention task (SAT) similar to the Continuous Performance Test (CPT), wherein participants attend to a continuous stream of two stimuli (Gabor patches) and respond to a pre-specified target; ADAS-Cog-13; Verbal Fluency;<br>PET and MEG | NBM OFF, 60Hz, 130Hz, GPi ON (washout NA)<br>+<br>GPi On/NBM Off vs GPi ON/NBM On 15-60 Hz (crossover - 8 weeks each) | Acute NBM stimulation effects on SAT: 60 Hz NBM signicantly increased accuracy compared to OFF (p=0.003). Performance did not differ between 130 Hz NBM and OFF. Chronic NBM stimulation effects (8 weeks): NBM did not improve ADAS-Cog-13 (p=0.41), letter VF (p=0.41), semantic VF (p=1), and SAT (p=0.70).<br><br>PET: Compared to OFF, NBM ON condition exhibited decreased metabolism in the right opercular part of the inferior frontal gyrus and supramarginal gyrus.<br>MEG: Compared to OFF, NBM ON showed increased delta/theta activity in the left frontal, parietal, and temporal lobe regions and increased beta/low gamma activity in the right occipital and cerebellar regions. | GPi DBS improved dyskinesia and motor fluctuations (p=0.04).<br><br>During a one-year follow-up (open label), 4 participants showed cognitive decline, converting from PD-MCI to PDD. NBM stimulation did not improve visual hallucinations.<br><br>VTA in the left NBM was associated with stable cognition, with additional voxels in the right GPi and GPe. Left NBM activation was functionally connected with medial temporal regions (normative dataset analysis). |

**Supplementary Table 4:** Demographic and clinical features of Parkinson's disease patients

| Autor, date | Sample Size | Sex | Age | Education level (years) | Disease duration (years) | Months since surgery | N patients with FoG / FoG severity | Med | Postoperative Motor score OFF / ON Stim | Postoperative LEDD | MoCA / MMSE / Mattis / FAB |
|---|---|---|---|---|---|---|---|---|---|---|---|
| **STN** | | | | | | | | | | | |
| **Wojtecki 2006** | 12 | 9 M \| 3 F | 64.0 (6.3) | - | - | 27.8 (17.8) | - | Off | UPDRS OFF 43 (13.3), 10Hz 44.9 (11.6), 130Hz 22.1 (7.5) | - | - |
| **Wojtecki 2011** | 12 PD / 12 HC | PD 6 M \| 6 F HC 6 M \| 6 F | PD 64.7 (8.6) HC 65.7 (4.8) | - | 18.6 (5.9) | 46.1 (25.9) | - | Off | UPDRS OFF 49 (3), 10Hz 45 (2), ≥130Hz 26 (3) | 957.5 (468.8) | Mattis 140.6 (2.8) |
| **Stegemoller 2013** | 17 | 14 M \| 3 F | 61.5 (9.5) | - | 13.59 (4.03) | 30.5 (19.9) | 8 Tremor / 9 Non Tremor Dominant | Off | UPDRS OFF 28.4 (10.7), 60Hz 24.5 (8.3), 130Hz 23.1 (5.8) | - | - |
| **Scangos 2016** | 1 | 1 M | 50 | - | 9 | 23 days after DBS | - | - | - | - | - |
| **Fagundes 2016** | 20 | 16 M \| 4 F | 56.65 (10.7) | 10.1 (5.2) | 15.30 (4.71) | 2.1 (1.38) | - | On | UPDRS 60Hz 34.3 (21.2), 130Hz 35.4 (19.2) | 1165.0 (615.1) | MMSE 26.5 (2.5) |
| **Amara 2016** | 20 | 15 M \| 5 F | 61.4 (8.85) | - | 10.1 (4.18) | 14.8 (IQR 6.3-27.8) | - | On | UPDRS OFF stim 34.2 (10.7), 60Hz 27.1 (8.6), 130Hz 28.9 (6.9) | 991.3 (674.7) | - |
| **Scangos 2017** | 15 | 13 M \| 2 F | 62.2 (6.4) | - | 7.7 (3.4) | after DBS surgery | - | - | - | - | - |
| **Varriale 2018** | 19 PD / 20 HC | PD 15 M \| 4 F HC 15 M \| 5 F | PD 58.9 (9.9) HC 62 (4.6) | - | 11.7 (4.1) | 3 | 16 (FOG-Q before DBS 17.5 (12), after 11.3 (8); item 3 FOG-Q before 2.3 (1) after 1.5 (1.3)) | Off / On | UPDRS (after surgery) OFF med 16.2 (8.8) ON med 10.6 (7.3) | 616.6 (386.9) | Mattis Before surgery 139.8 (4.0), After surgery 137.0 (5.6) |
| **Kelley 2018** | 10 | 6 M \| 4 F | 64.8 (7.1) | - | NA | > 3 (NA) | - | On | UPDRS OFF 24.8 (10.9) 120-150Hz 14.0 (9.0) | 857.2 (722.3) | - |
| **Grover 2018** | 15 | 12 M \| 3 F | - | - | 18.5 (3.7) | 72 (42) | - | Off | UPDRS ON med/ON stim 33.7 (9.3) | - | - |
| **Imbalzano 2020** | 1 | 1 M | 47 | - | 16 | after DBS surgery | No FOG | - | - | 600 | - |
| **Mandali 2020** | 24 | 18 M \| 6 F | 59.7 (11.8) | - | 9.6 (3.8) | after DBS surgery | - | - | - | - | MoCA 24.5 (2.75) |
| **Lam 2021** | 12 | 12 M \| 0 F | 60.75 (8.6) | 16.5 (1.5) | NA | 20.98 (14.70) | - | On | UPDRS OFF 25.5 (10.2) 130-135Hz 9.14 (5.40) | - | - |
| **Lee 2021** | 9 | 6 M \| 3 F | 60.9 (5.5) | 15.8 (1.6) | 11.6 (4.9) | 2-4 days after DBS | - | - | - | - | - |
| **Romangnolo 2021** | 12 | 11 M \| 1 F | 61.7 (9.3) | - | 15.3 (3.8) | 36.0 (25.2) | - | Off | UPDRS OFF med/ON stim 30.5 (5.1), ON med/ON stim 17.8 (9.4) | - | MMSE 28.2 (1.1) |
| **Waldthaler 2023** | 14 | 10 M \| 4 F | 57.0 (8.8) | - | 8.6 (3.3) | 10.2 (9.1) | - | Off | MDS-UPDRS OFF 38.9 (9.9), 130Hz 15.7 (8.7) | 476 (307) | MoCA 26.5 (2.3) |

| | | | | | | | | | | | |
|---|---|---|---|---|---|---|---|---|---|---|---|
| **Zacharia 2023** | 20 PD / 16 HC | PD 16 M \| 6 F HC 10 M \| 6 F | PD 63.7 (9.1) HC 66.1 (5.4) | - | - | 46.4 (18.0) | - | Off | UPDRS 80Hz 16.9 (10.5), 130Hz 19.2 (9.1) | 560.81 (298.5) | MMSE PD 28.8 (1.2) HC 28.7 (1.4), FAB PD 15.1 (3.0) HC 17.1 (1.3) |
| **Muhammad 2023** | 24 | 18 M \| 6 F | 59.7 (11.8) | - | 9.6 (3.8) | after DBS surgery | - | On | - | - | MoCA 24.5 (2.8) |
| **Wang 2023** | 20 | 16 M \| 4 F | 60.25 (8.85) | 12.4 (3.6) | 10.1 (4.8) | 11.7 (5.7) | - | On | MDS-UPDRS OFF 39 (16), 10Hz 39 (15), 130Hz 28 (12) | 363.25 (241.0) | - |
| **Qin 2023** | 29 | 17 M \| 12 F | 62.7 (7.4) | 10.9 (3.4) | 8.9 (3.2) | 1 | PIGD, TD/PIGD ratio 0,38 (0,33) | Off | MDS-UPDRS OFF 34.6 (15.6), 5Hz 26.5 (13.1), 130Hz 17.6 (9.3) | 666.5 (444.1) | MMSE 27.9 (1.8) |
| **Memon 2023** | 15 | 11 M \| 4 F | 61.8 (9.5) | - | 10.0 (3.7) | 13.5 (IQR 8.1-23.4) | - | - | UPDRS ON med/OFF stim 31.0 (IQR 28.8-40.3) | 968.9 (673.3) | - |
| **Herz 2024** | 18 (9 LFS, 9 HFS) | 12 M \| 6 F | 61.7 (5.7) | - | 10.1 (4.1) | 11 tested 2-4 days after DBS surgery, 7 chronically with DBS | - | - | MDS-UPDRS 27.3 (18.1) | 866.9 (391.1) | MoCA 26.1 (2.7) |
| **Salehi 2024** | 20 | 16 M \| 4 F | 57.1 (6.8) | - | 8.60 (3.05) | > 3 (NA) | - | Off | UPDRS OFF 39.1 (13.7) HFS ON 15.9 (9.8) | 423.75 (267.5) | MoCA 27.9 (1.5) |
| **Busteed 2024** | 38 (10 LFS, 14 HFS, 14 no DBS) | noDBS 8M \| 6F, LFS 2M \| 8F, HFS 7M \| 7F | noDBS 63.6 (9.0), LFS 62.2 (6.0), HFS 62.7 (5.4) | noDBS 10.7 (3.0), LFS 11.0 (4.1), HFS 10.3 (4.7) | noDBS 11.7 (4.1), LFS 15.5 (7.96), HFS 16.7 (6.4) | LFS 39.6 (30.1), HFS 50.5 (44.4) | - | On | - | noDBS 1464.1 (783.6), LFS 1075,5 (376.9), HFS 1144.0 (438.1) | - |
| **Schoenwald 2025** | 20 | 15 M \| 5 F | 64.9 (8.2) | 14.3 (2.5) | 12.4 (7.1) | 20.6 (21.3) | - | Off | - | 789.00 (554.30) | MoCA 24.8 (2.9), Mattis 140.3 (3.4) |
| **McAuley 2025** | 1 | 1 M | 54 | - | 14 | 48 | - | On | MDS-UPDRS ON/ON 20 | 500 | - |
| **Imbalzano 2025** | 12 | 10 M \| 2 F | 56.7 (8.0) | 11.2 (3.2) | 14.1 (3.7) | 37.3 (15.0) | - | On | MDS-UPDRS HFS 23.5 (12.6), dual 23.0 (8.5) | - | - |
| **Cole 2025** | 15 | 12 M \| 3 F | 68.7 (IQR 7.7) | 14 (IQR 3.5) | 9 (IQR 5.5) | 24.1 (25.1) | - | On | MDS-UPDRS OFF 35.1 (19.4), 4Hz 27.1 (15.2); 130Hz 20.9 (13.1) | 948.9 (689.2) | MoCA 25 (range 4) |
| **Ricciardi 2025** | 18 | 14 M \| 4 F | 62.2 (7.4) | - | 15.7 (5.4) | 47.2 (17.7) | - | On | Composite Motor Score 3.4, 3.10, 3.17 OFF 8.2 (3.5), 130Hz 3.9 (2.3), 10Hz 5.6 (2.6), 130+10hz 4.1 (2.5) | 745.2 (399.5) | MoCA 27 (1.8) |
| **Xie 2025** | 18 | 9 M \| 11 F | 63.2 (7.9) | 11.9 (2.5) | 9.5 (4.8) | NA | 18 (FOG-Q 16.2 (5.4), item 3 FOG-Q 3.1 (0.8)) | Off | MDS-UPDRS OFF 45.3 (18.5), 5Hz 42.9 (17.0), 130Hz 21.5 (9.1) | 476.2 (220.2) | MoCA 23.9 (3.5), MMSE 26.4 (2.4) |
| **Kricheldorff 2025** | 17 | 12 M \| 5 F | 63.6 (7.1) | - | 15.1 (6.1) | 38.4 (27.6) | - | On | UPDRS OFF 35.7 (16.3), 20Hz 33.4 (14.5), 130Hz 24.1 (8.5) | 524.4 (398.6) | MMSE 28.2 (1.6) |

| | | | | | | | | | | | |
|---|---|---|---|---|---|---|---|---|---|---|---|
| **PPN** | | | | | | | | | | | |
| **Costa 2010** | 5 PD PPN / 5 noPPN / HC | 18 M \| 0 F | PPN 60.6 (7.7), noPPN 39.6 (6.5), HC 57.6 (6.2) | PPN 8.8 (4.0), noPPN 8.8 (4.0), HC 11.0 (5.3) | PPN 10.5 (2.4), noPPN 8.2 (3.9) | age at DBS 45.7 (9.0) | - | Off | UPDRS PPN Off 58.5 (17.7), PPN On 41.5 (5.6), noPPN 39.6 (6.5) | range 375–625 | MMSE PPN 27.4 (1.1), noPPN 28.1 (1.3), HC 29.1 (1.3) |
| **Arnulf 2010** | 2 | - | 1. 68<br>2. 57 | - | 1. 16<br>2. 29 | 12 | 2 Severe | Off | - | 1. 900mg;<br>2. 0mg | - |
| **Alessandro, 2010 / Ceravolo 2011** | 6 | - | 62.8 (2.2) | - | 11.8 (3.5) | 12 (NA) | Severe axial, UPDRS item 29 OFF med 3.7 (0.5) ON med 2.3 (0.5) | On | - | - | - |
| **Thevathasan 2010** | 11 PD / HC | - | 64.5 (6.8) | - | 11.6 (5.0) | 12.7 (12.6) | - | Off | UPDRS OFF med 46.5 (14.2), ON med 27.3 (9.5), OFF stim 46.5 (14.2), ON stim 44.9 (14.9) | 1227.3 (510.1) | MMSE 28.5 (1.4) |
| **Nosko 2014** | 9 | 7 M \| 2 F | 64 (5.5) | - | 22.3 (6.4) | 9 (NA) | 9 (UPDRS item 14 OFF = 3 ±1) | Off / On | UPDRS Off/Low 24.0 (13.1); Off/High 29.1 (13.0); On/Low 19.6 (10.8); On/High 22.5 (10.4) | 689.4 (351.0) | - |
| **Fischer 2015** | 8 | 3 M \| 5 F | 66.4 (7.6) | - | 8.88 (3.0) | 15.4 (9.1) | 8 | On | - | 1027.4 (774.0) | Mattis 138.0 (5.2) |
| **Strumpf 2016** | 5 | - | 62.8 (9.5) | - | - | 8.4 (4.8) | Gait impairment | - | - | - | Mattis >131 |
| **Skvortsova 2021** | 3 | 1 M \| 2 F | 60.0 (12.5) | - | - | - | - | Off | UPDRS OFF med/OFF stim ON 18.0 (1.73) med/OFF stim 44.3 (6.0) | 973.3 (371.0) | Mattis 140.0 (1.0) MMSE 25.7 (1.2) |
| **NBM** | | | | | | | | | | | |
| **Gratwicke 2017** | 6 | 6 M \| 0 F | 65.2 (10.7) | - | 12.7 (2.3) | tested after DBS surgery | - | - | - | 646.9 (204.7) | Median (IQR) MMSE baseline 24.5 (4), OFF 21 (5), ON 23 (9) Mattis baseline 113 (25), OFF 117 (27), ON 116 (21) |
| **Nombela 2019** | 1 | 1 F | 68 | - | 7 | 12 | - | On | UPDRS ON med/ON stim 16 | 1000 | MMSE baseline 27, GPi 25, NBM+GPi 23; Mattis baseline 134, GPi 110, NBM+GPi 111 |
| **Sasikumar 2021** | 6 | 5 M \| 1 F | 65.7 (3.9) | - | 11.5 (1.2) | - | - | On | - | 1374.5 (412.2) | Mattis pre DBS 130.2 (5.0), post DBS 123.3 (6.4) |

**Supplementary Table 5:** DBS parameters of Parkinson's disease patients

| Autor, date | n | Target DBS | Electrode contacts | TEED adjustament | Voltage/Amplitude | Pulse | Frequency |
|---|---|---|---|---|---|---|---|
| **Wojtecki 2006** | 12 | STN | Medtronic, model 3389 | No | Left 3.4 (0.7)<br>Right 3.0 (0.6) | Left 70.0 (14.9)<br>Right 67.5 (18.7) | Left 157.1 (20.6)<br>Right 152.5 (19.9) |
| **Wojtecki 2011** | 12 | STN | Medtronic, model 3389 | No | Left 2.9 (0.7)<br>Right 2.9 (0.7) | Left 62.5 (8.7)<br>Right 65.0 (17.3) | 137.5 (15.5) |
| **Stegemoller 2013** | 17 | STN | - | No | - | - | - |
| **Scangos 2016** | 1 | STN | - | - | - | - | - |
| **Fagundes 2016** | 20 | STN | Medtronic, model 3389 | No | Left 3.0 (0.7)<br>Right 3.0 (0.6) | Left 79.5 (17.6)<br>Right 81.0 (17.6) | 124.0 (26.0) |
| **Amara 2016** | 20 | STN | - | No | 4.01 (0.74) | 82.3 (25.1) | 158.4 (14.7) |
| **Scangos 2017** | 15 | STN | Medtronic, model 3389 | No | 1.3 (0.7) | 90-120 | - |
| **Varriale 2018** | 19 | STN | - | YES | Left 2.7 (0.60)<br>Right 2.7 (0.7) | Left 67.9 (13.6)<br>Right 66.3 (12.6) | Left 135.3 (41.0)<br>Right 132.9 (35.5) |
| **Kelley 2018** | 10 | STN | - | No | Left 2.4 (1.0)<br>Right 2.5 (1.1) | Left 80.0 (32.7)<br>Right 82.0 (29.4) | Left 132.0 (7.5)<br>Right 133.5 (6.3) |
| **Grover 2018** | 15 | STN | - | YES | Left 3.4 (1.2)<br>Right 3.3 (0.8) | 60 | 96.7 (24.4) |
| **Imbalzano 2020** | 1 | STN | Medtronic, model 3389 | No | - | - | - |
| **Mandali 2020** | 24 | STN | SceneRay, model 1510 | YES | 1.8 (0.4) mA for 130 Hz<br>6.5 (1.5) mA for 10 Hz | 90 | - |
| **Lam 2021** | 12 | STN | - | - | Left 2.3 (0.4)<br>Right 2.0 (0.3) | Left 68.8 (11.9)<br>Right 69.2 (16.1) | 131 (5) |
| **Lee 2021** | 9 | STN | Medtronic, model 3387 | No | 2,5 | 100 | - |
| **Romangnolo 2021** | 12 | STN | Medtronic, model 3389 | YES | 3.1 (0.9) | 60.5 (0.9) | 121.7 (19.5) |
| **Waldthaler 2023** | 14 | STN | Vercise Cartesia Directional | No | Left 1.8 (0.8)<br>Right 1.6 (0.6) | 60.8 (9.5) | 130.0 (0) |
| **Zacharia 2023** | 20 | STN | Medtronic, model 3389 | YES | Left 2.9 (0.7)<br>Right 3.0 (0.7) | 60 | Left 115.7 (29.1)<br>Right 112.9 (24.7) |
| **Muhammad 2023** | 24 | STN | SceneRay, model 1510 | YES | 1.8 (0.4) mA for 130 Hz<br>6.5 (1.45) mA for 10 Hz | 90 | - |
| **Wang 2023** | 20 | STN | - | No | Left 2.3 (0.6)<br>Right 2.3 (0.6) | 90 | - |
| **Qin 2023** | 29 | STN | Model L301, PINS Medical | No | Left 2.4 (3.2)<br>Rigth 1.7 (0.6) | 60 | - |
| **Memon 2023** | 15 | STN | - | No | 3.9 (0.7) | 77.5 (24.9) | 158.4 (14.8) |
| **Herz 2024** | 18 | STN | 17 Abbott, 1 Medtronic | No | Left 2.5 (1.4)<br>Right 2.1 (0.9) | Left 55.0 (14.3)<br>Right 53.9 (10.4) | - |

| | | | | | | | |
|---|---|---|---|---|---|---|---|
| **Salehi 2024** | 20 | STN | Vercise Cartesia Directional | No | Left 2.6 (1.3)<br>Right 2.3 (1.1) | Left 60 (10)<br>Right 61.5 (6.7) | 123.9 (15.2) |
| **Busteed 2024** | 24 | STN | - | No | LFS Left 3.3 (1.1),<br>Right 3.0 (1.1),<br>HFS Left 2.6 (0.8)<br>Right 2.7 (0.6) | LFS Left 63 (17.0),<br>Right 66 (19.0),<br>HFS Left 68.6 (14.1),<br>Right 68.6 (18.3) | LFS 62.0 (16.4),<br>HFS 146.8 (23.7) |
| **Schoenwal 2025** | 20 | STN | 19 Abbott Infinity,<br>1 Boston Cartesia | YES | Left 1.7 (1.1)<br>Right 1.7 (1.1) | 60 | Left 131.4 (6.9)<br>Right 132.7 (9.2) |
| **McAuley 2025** | 1 | STN | Vercise Cartesia Directional | No | Left 3.1<br>Right 2.9 | 60 | 60-179 |
| **Imbalzano 2025** | 12 | STN | Vercise Cartesia Directional | No | Left 3.0 (0.7)<br>Right 2.9 (0.8) | 60 | 140.0 (25.6) |
| **Cole 2025** | 15 | STN | Medtronic Activa or Percept | No | Left 2.1 (1.1)<br>Right 2.4 (1.2) | Left 70.0 (15.6)<br>Right 72.0 (17.0) | 133.7 (10.1) |
| **Ricciardi 2025** | 18 | STN | Vercise Cartesia X, Boston | No | Left 3.4 (1.2)<br>Right 3.7 (1.4) | Left 54.4 (7.8)<br>Right 51.1 (10.2) | 132.7 (11.5) |
| **Xie 2025** | 18 | STN | Medtronic, model 3389 | No | Left 2.4 (0.8)<br>Right 2.6 (0.8) | Left 73.3 (13.3)<br>Right 73.9 (12.4) | 124.2 (24.3) |
| **Kricheldorff 2025** | 17 | STN | Medtronic, model 3389 | No | Left 2.9 (0.8)<br>Right 2.7 (1.0) | 60 | 164.7 (23.8) |
| **Arnulf 2009** | 2 | PPN | Medtronic, model 3389 | No | Patient 1: Right 2.7v, Left 3v;<br>Patient 2: Right 2.8v, Left 1.5v | 60 | 10-25 |
| **Costa 2010** | 5 | PPN | Medtronic, model 3389 | No | 2 (range 1.8–2.2) | 60 | 25 |
| **Alessandro 2010 / Ceravolo 2011** | 6 | PPN | Medtronic, model 3389 | No | 1.5-2 | 60 | 25 |
| **Thevathasan 2010** | 11 | PPN | Medtronic, model 3387 | No | 2.9 (0.6) | 60 | 26.8 (5.6) |
| **Nosko 2014** | 9 | PPN | - | No | Left 1.6 (0.7)<br>Right 1.6 (0.5) | 60 (1 patient with 450) | Low 17.8 (5.7)<br>High 68.9 (10.5) |
| **Fischer 2015** | 8 | PPN | Medtronic, model 3389 | No | 2.0 (0.7) | 60 | - |
| **Strumpf 2016** | 5 | PPN | Medtronic, model 3389 | - | 1.8 (0.6) | 60 | - |
| **Skvortsova 2021** | 3 | PPN | Medtronic, model 3389 | | Left 1.8 (1.1)<br>Right 2.1 (1.2) | 50.0 (17.3) | 33.3 (11.6) |
| **Gratwicke 2017** | 6 | NBM | Medtronic, model 3389/3387 | No | 2.75 (0.6) | 60 | 20 |
| **Nombela 2019** | 1 | NBM+GPi | Vercise Standard (8 contacts) | No | - | - | - |
| **Sasikumar 2021** | 6 | NBM+GPi | - | No | GPi Left 4.7 (1.6), Right 4.8 (1.6);<br>NBM Left 5.3 (2.7), Right 5.4 (2.7) | GPi 56.7 (5.2); NBM Left 53.3 (12.1), Right 45.8 (23.3) | GPi 126.5 (38.6);<br>NBM 24.2 (17.8) |
