## Supplementary Figure for "Frequency-dependent cognitive effects of Deep Brain Stimulation in Parkinson’s Disease: A Systematic Review and Meta-Analysis"

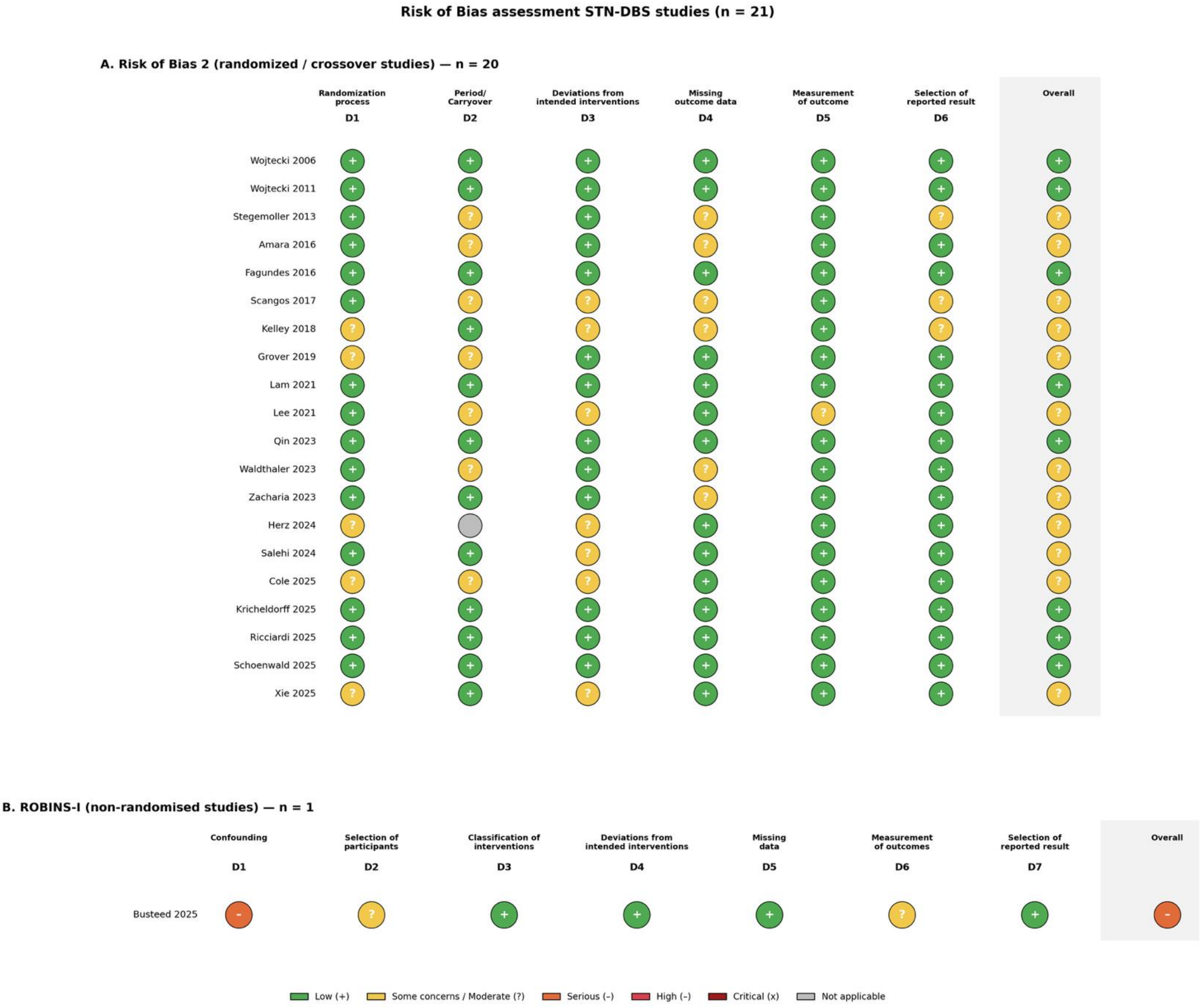


**Supplementary Figure 1. Study Quality Assessment.** The methodological quality of the included studies was assessed using the Cochrane Risk of Bias 2 (RoB-2) tool for randomized and crossover trials and the ROBINS-I tool for non-randomized studies. Risk-of-bias was low or raised only some concerns in the randomized and crossover trials, with the most frequent uncertainties related to carryover effects (inherent to crossover DBS designs). A single non-randomized study was rated as high confounding risk

### Comparison of Low+Very Low-frequency stimulation with High-frequency stimulation

### Low+Very Low-frequency vs High-frequency stimulation
### Main Domains

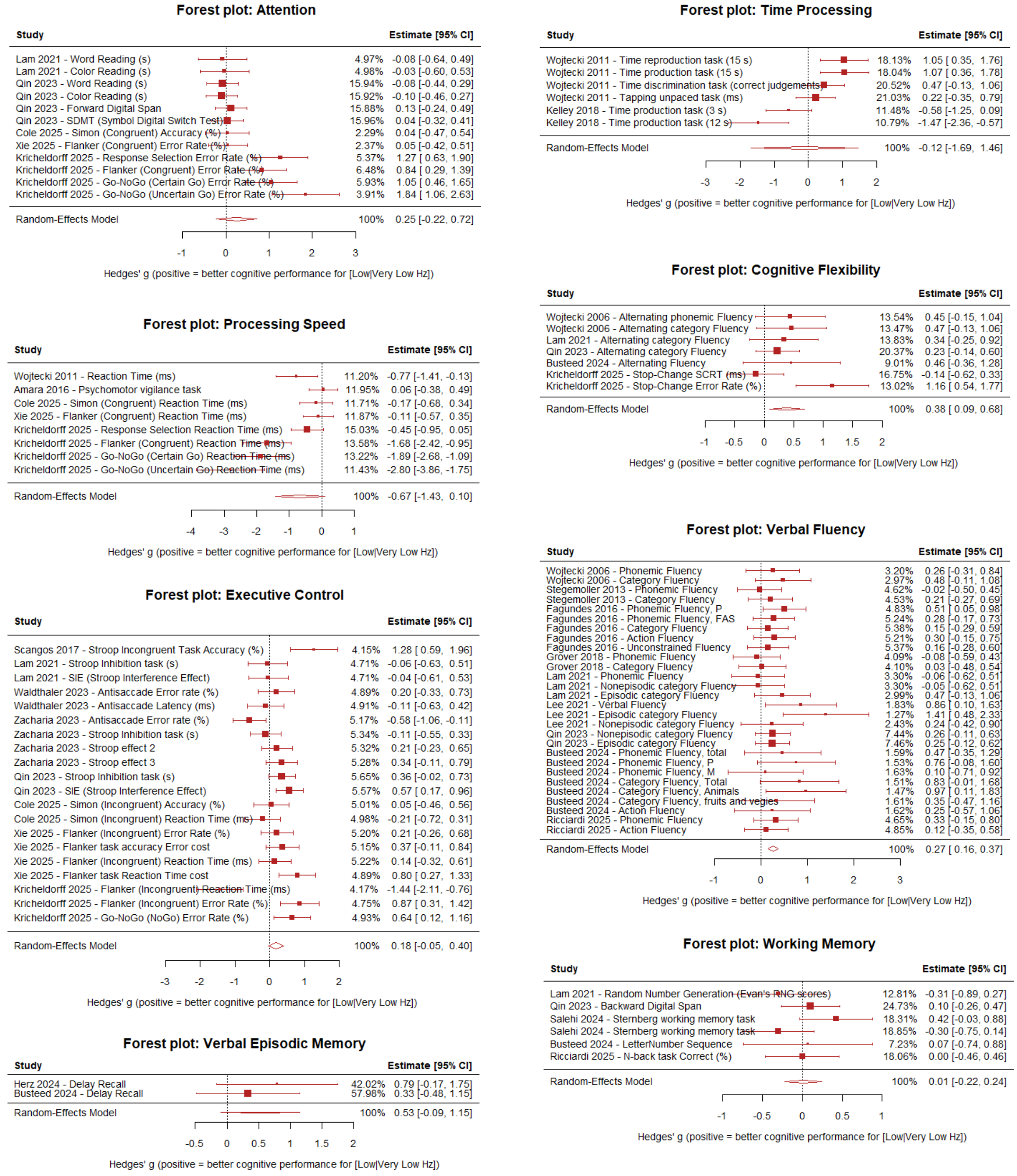


**Supplementary Figure 2. Effects of Low+Very Low- vs High-Frequency STN-DBS on the main cognitive domains.** Multilevel meta-analysis adjusting for study-level dependence. Forest plot showing Hedges' g effect sizes for the main cognitive domains comparing Low+Very Low-frequency versus High-frequency STN-DBS across studies, estimated using a multilevel random-effects meta-analysis that accounts for dependency among multiple cognitive domains measurements within the same study (effects nested within study). Positive values indicate better cognitive domain performance under Low+Very Low-frequency stimulation relative to High-frequency stimulation. Low- and very low-frequency stimulation was associated with modest but statistically significant improvements in verbal fluency and cognitive flexibility compared with High-frequency stimulation. No significant differences were observed across the remaining cognitive domains.

Verbal Fluency (k=28, 9 studies, g=0.27, SE=0.06, 95% CI [0.16, 0.37], p<0.001, p(FDR)<0.001), Cognitive Flexibility (k=7, 5 studies, g=0.38, SE=0.15, 95% CI [0.09, 0.68], p=0.011, p(FDR)=0.043), Processing Speed (k=8, 5 studies, g=-0.67, SE=0.39, 95% CI [-1.43, 0.10], p=0.09, p(FDR)=0.192), Attention (k=12, 5 studies, g=0.25, SE=0.24, 95% CI [-0.22, 0.72], p=0.30, p(FDR)=0.404), Time Processing (k=6, 2 studies, g=-0.12, SE=0.80, 95% CI [-1.69, 1.46], p=0.86, p(FDR)=0.926), Executive Control (k=20, 8 studies, g=0.18, SE=0.12, 95% CI [-0.05, 0.40], p=0.12, p(FDR)=0.192), and Working Memory (k=6, 5 studies, g=0.01, SE=0.12, 95% CI [-0.22, 0.24], p=0.93, p(FDR)=0.926).

### Low+Very Low-frequency vs High-frequency stimulation Verbal Fluency Subdomains

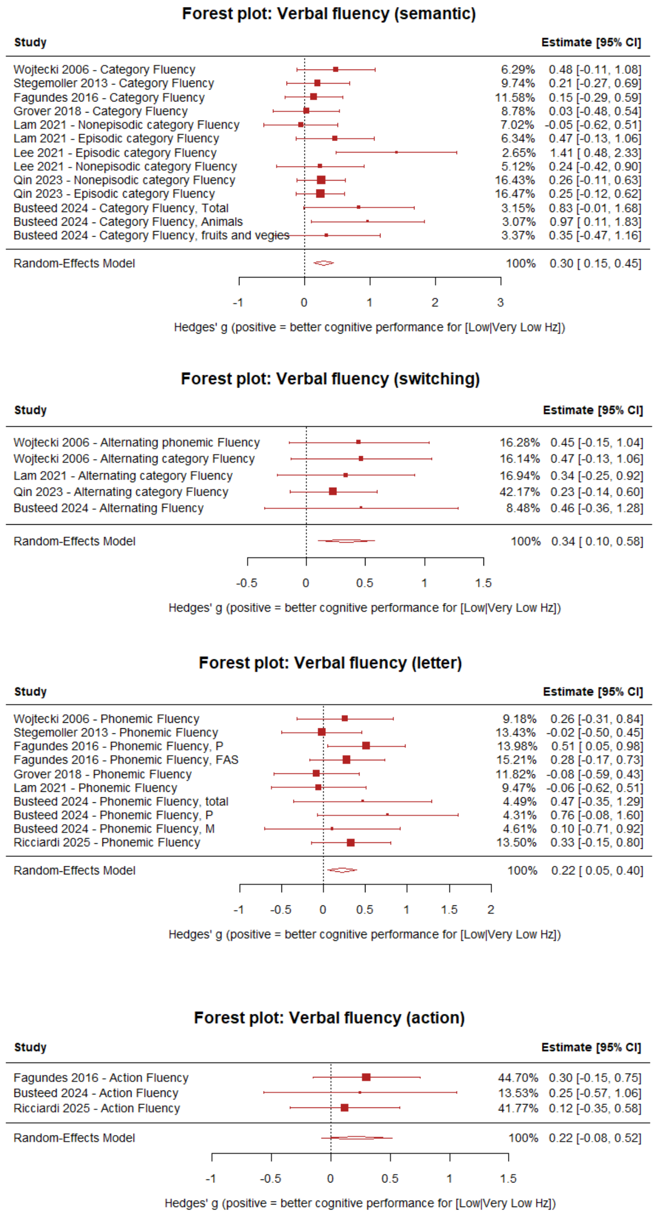


**Supplementary Figure 3. Effects of Low+Very Low- vs High-Frequency STN-DBS on the main Verbal Fluency sub domains.** Multilevel meta-analysis adjusting for study-level dependence. Forest plot showing Hedges' g effect sizes for the verbal fluency subdomains comparing Low+Very Low-frequency versus High-frequency STN-DBS across studies, estimated using a multilevel random-effects meta-analysis that accounts for dependency among multiple cognitive domains measurements within the same study (effects nested within study). Positive values indicate better cognitive performance under Low+Very Low-frequency stimulation relative to High-frequency stimulation. Across verbal fluency subdomains Low+Very Low-frequency consistently outperformed High-frequency.

Significant effects of comparable effects sizes were observed for Semantic VF (k=13, g=0.30, SE=0.08, 95% CI [0.15, 0.45], p=0.001), Letter VF (k=10, g=0.22, SE=0.09, 95% CI [0.05, 0.40], p=0.012) and Switching VF (k=5, g=0.34, SE=0.12, 95% CI [0.10, 0.58], p=0.005). Action VF showed a non-significant effect in the same direction (k=3, g=0.22, SE=0.15, 95% CI [-0.082, 0.52], p=0.155).

### Low+Very Low-frequency vs High-requency stimulation Executive Control Subdomains

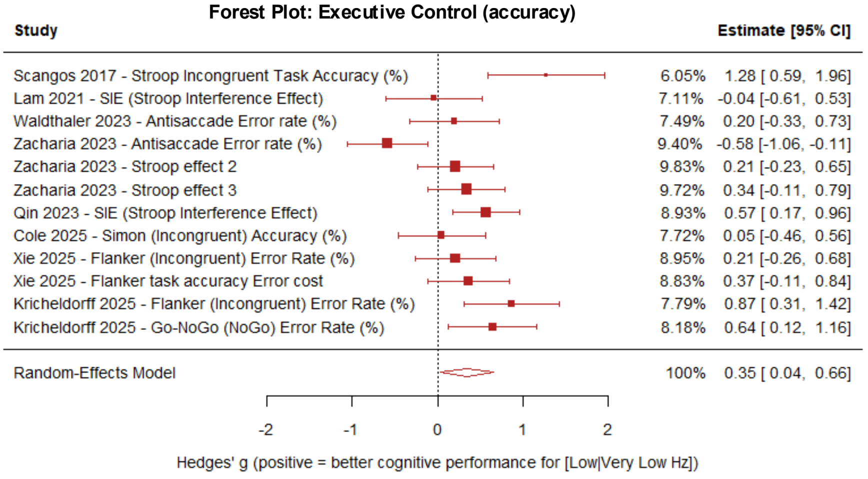


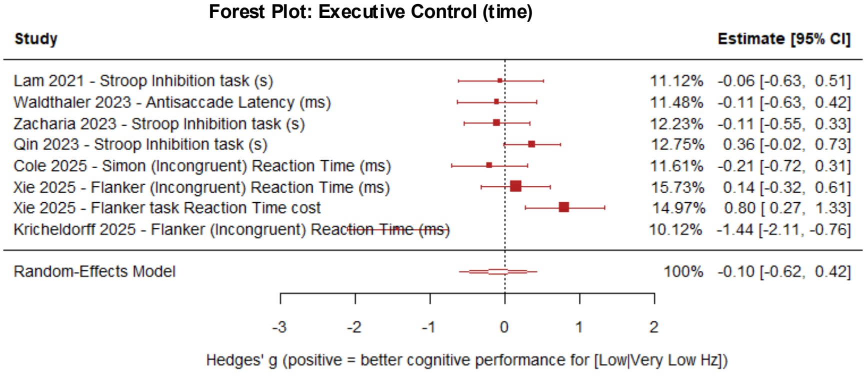


**Supplementary Figure 4. Effects of Low+Very Low- vs High-Frequency STN-DBS on the main Executive Control sub domains.** Multilevel meta-analysis adjusting for study-level dependence. Forest plot showing Hedges' g effect sizes for the Executive Control subdomains (accuracy and time metrics) comparing Low+Very Low-frequency versus High-frequency STN-DBS across studies, estimated using a multilevel random-effects meta-analysis that accounts for dependency among multiple cognitive domains measurements within the same study (effects nested within study). Positive values indicate better cognitive performance under Low+Very Low-frequency stimulation relative to High-frequency stimulation. Comparison of low- and very low-frequency stimulation (LFS+VLFS) versus high-frequency stimulation (HFS) on executive control in STN-DBS.

LFS+VLFS was associated with significantly better performance on accuracy-based executive control measures (k=12, g=0.35, SE=0.14, 95% CI [0.04, 0.65], p=0.032), whereas a non-significant negative signal was observed for time-based measures (k=8, g=-0.10, SE=0.22, 95% CI [-0.62, 0.42], p=0.666) depending on motor performance.

### Low+Very Low-frequency vs High-frequency stimulation
### Main + Secondary + Tertiary Cognitive Domains

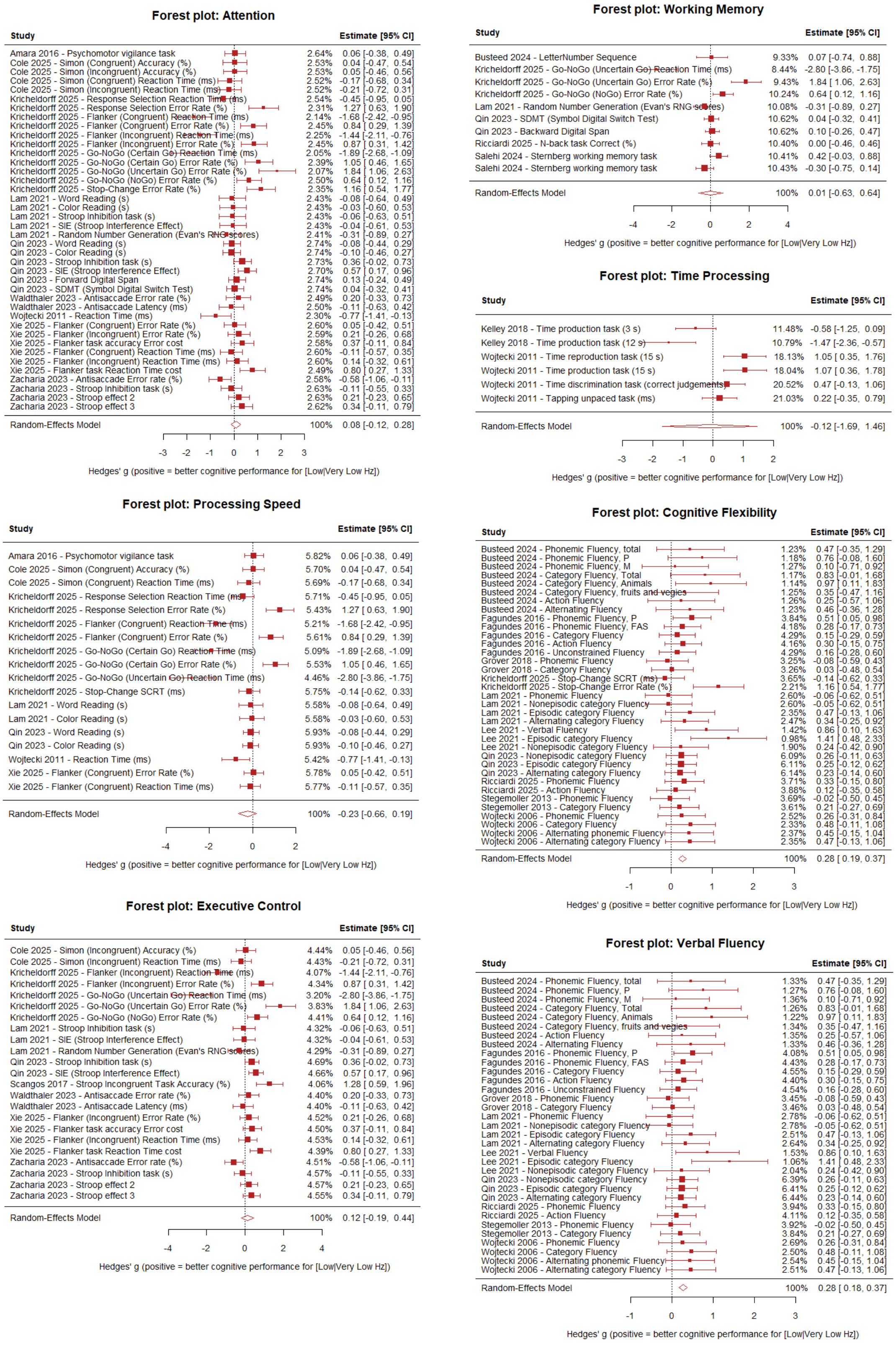


**Supplementary Figure 5. Effects of Low+Very Low- vs High-Frequency STN-DBS on the overall cognitive domains regardless of aggregation/classification level (Main + Secondary + Tertiary).** Multilevel meta-analysis adjusting for study-level dependence. Forest plot showing Hedges' g effect sizes for the overall aggregated cognitive domains comparing Low+Very Low-frequency versus High-frequency STN-DBS across studies, estimated using a multilevel random-effects meta-analysis that accounts for dependency among multiple cognitive domains measurements within the same study (effects nested within study). Positive values indicate better cognitive performance under Low+Very Low-frequency stimulation relative to High-frequency stimulation. Low- and very low-frequency stimulation was associated with modest but statistically significant improvements in verbal fluency and cognitive flexibility compared with High-frequency stimulation. No significant differences were observed across the remaining cognitive domains: Attention (p=0.416), Executive Control (p=0.440), Processing Speed (p=0.274), Time Processing (p=0.886), and Working Memory (p=0.984).

### Comparison of Low-frequency stimulation with High-frequency stimulation

### Low-frequency vs High-frequency stimulation
### Main Domains

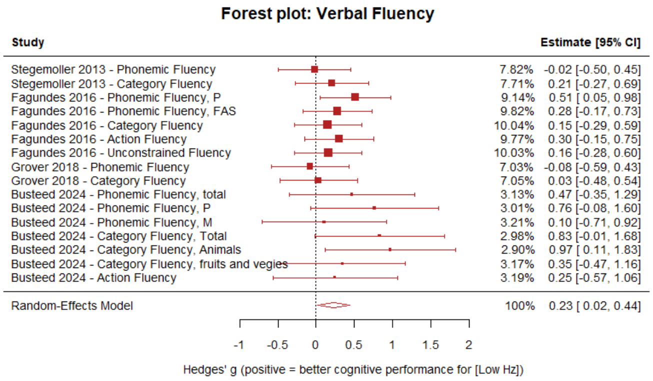


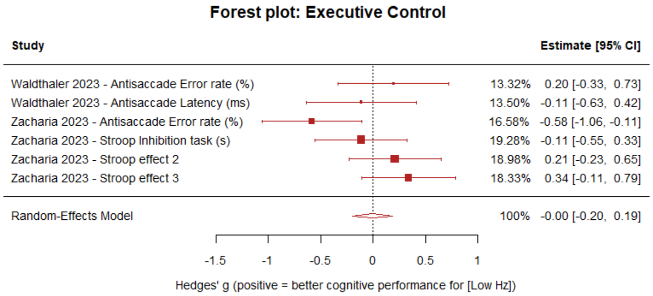


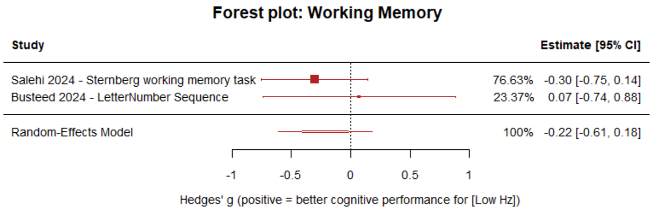


**Supplementary Figure 6. Effects of Low- vs High-Frequency STN-DBS on the main cognitive domains.** Multilevel meta-analysis adjusting for study-level dependence. Forest plot showing Hedges' g effect sizes for the main cognitive domains comparing Low-frequency versus High-frequency STN-DBS across studies, estimated using a multilevel random-effects meta-analysis that accounts for dependency among multiple measures within the same study (effects nested within study). Positive values indicate better performance under Low-frequency stimulation relative to High-frequency stimulation. Low-frequency stimulation was associated with modest but statistically significant improvements in verbal fluency compared with High-frequency stimulation. No significant differences were observed across the remaining cognitive domains.

### Low-frequency vs High-frequency stimulation
### Verbal Fluency Subdomains

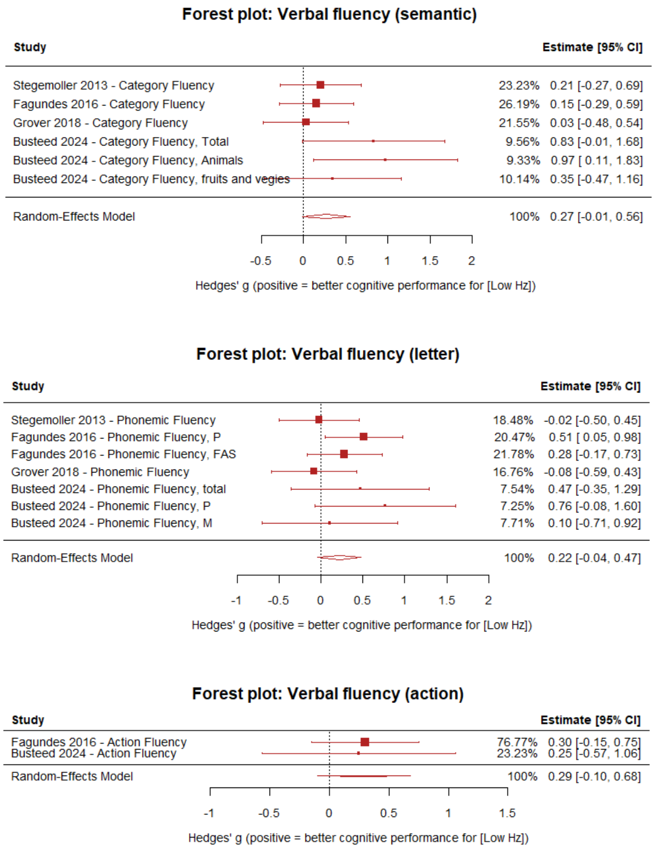


**Supplementary Figure 7. Effects of Low- vs High-Frequency STN-DBS on the main Verbal Fluency sub domains.** Multilevel meta-analysis adjusting for study-level dependence. Forest plot showing Hedges' g effect sizes for the verbal fluency subdomains comparing Low-frequency versus High-frequency STN-DBS across studies, estimated using a multilevel random-effects meta-analysis that accounts for dependency among multiple measures within the same study (effects nested within study). Positive values indicate better performance under Low-frequency stimulation relative to High-frequency stimulation. Across verbal fluency subdomains Low-frequency consistently tend to outperform High-frequency (Semantic VF, k=6, g=0.27, SE=0.14, 95% CI [-0.01, 0.56], p=0.06, Letter VF, k=7, g=0.22, SE=0.13, 95% CI [-0.04, 0.48], p=0.09, Action, k=2, g=0.29, SE=0.20, 95% CI [-0.10, 0.68], p=0.149).

### Low-frequency vs High-frequency stimulation
### Main + Secondary + Tertiary Cognitive Domains

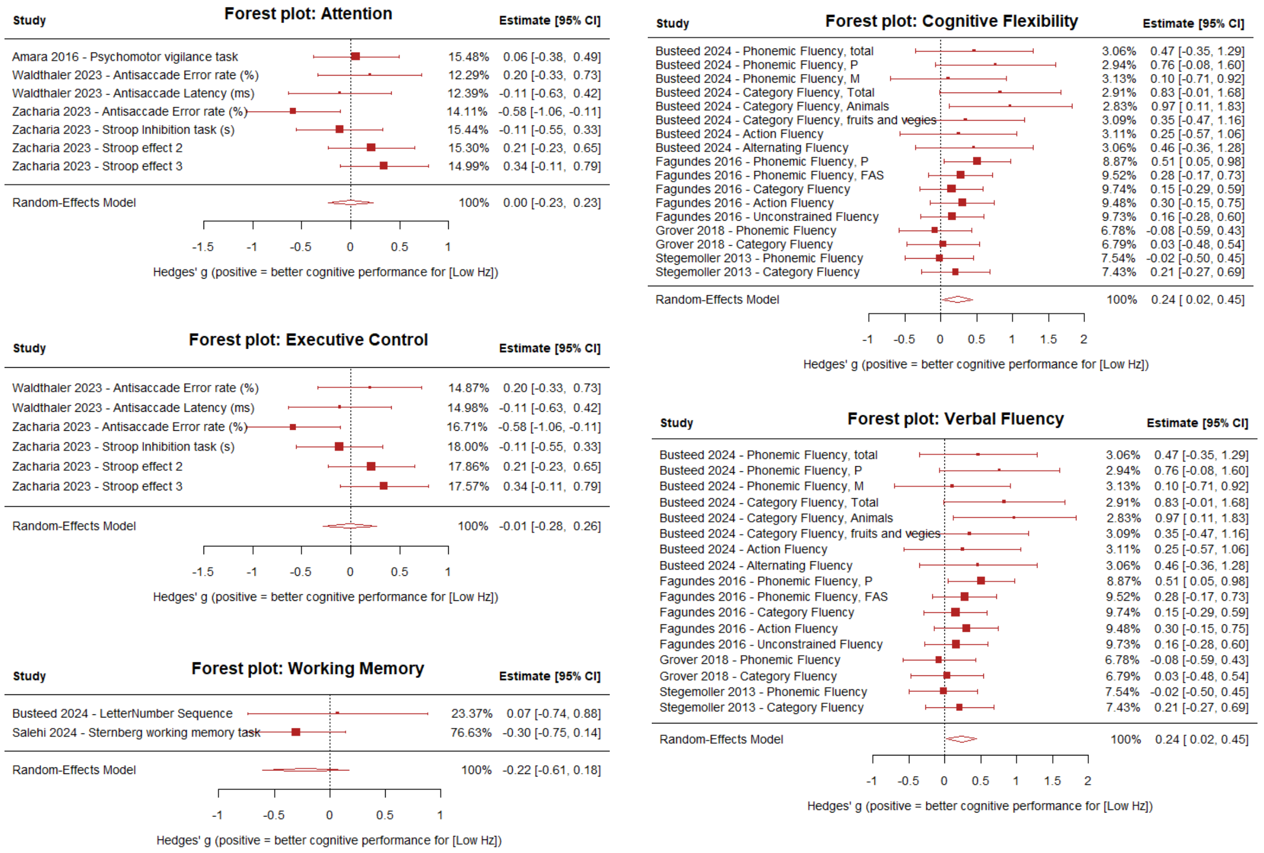


**Supplementary Figure 8. Effects of Low- vs High-Frequency STN-DBS on the overall cognitive domains regardless of aggregation/classification level (Main + Secondary + Tertiary).** Multilevel meta-analysis adjusting for study-level dependence. Forest plot showing Hedges' g effect sizes for the overall aggregated cognitive domains comparing Low-frequency versus High-frequency STN-DBS across studies, estimated using a multilevel random-effects meta-analysis that accounts for dependency among multiple cognitive domains measurements within the same study (effects nested within study). Positive values indicate better cognitive performance under Low-frequency stimulation relative to High-frequency stimulation. Low- -frequency stimulation was associated with modest but statistically significant improvements in verbal fluency and cognitive flexibility compared with High-frequency stimulation (p=0.031). No significant differences were observed across the remaining cognitive domains: Attention (p=0.980), Executive Control (p=0.958), and Working Memory (p=0.281).

### Comparison of Very Low-frequency stimulation with High–frequency stimulation

### Very Low-frequency vs High-frequency stimulation
### Main Domains

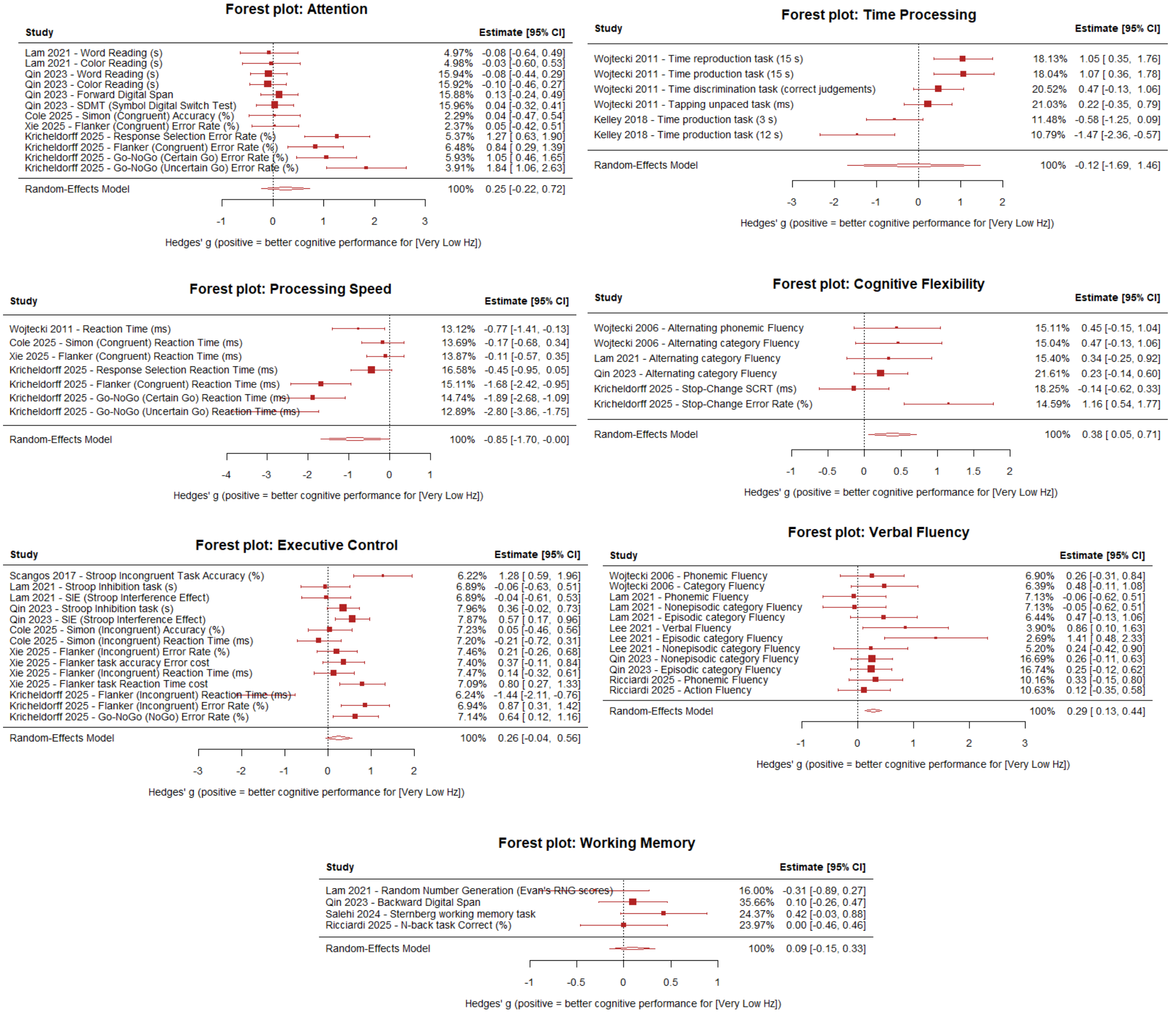


**Supplementary Figure 9. Effects of Very Low- vs High-Frequency STN-DBS on the main cognitive domains.** Multilevel meta-analysis adjusting for study-level dependence. Forest plot showing Hedges' g effect sizes for the main cognitive domains comparing Very Low-frequency versus High-frequency STN-DBS across studies, estimated using a multilevel random-effects meta-analysis that accounts for dependency among multiple cognitive domains measurements within the same study (effects nested within study). Positive values indicate better cognitive domain performance under Very Low-frequency stimulation relative to High-frequency stimulation. Very low-frequency stimulation was associated with modest but statistically significant improvements in verbal fluency and cognitive flexibility compared with High-frequency stimulation. No significant differences were observed across the remaining cognitive domains.

Verbal Fluency (k=12, 5 studies, g=0.29, SE=0.08, 95% CI [0.14, 0.44], p<0.001), Cognitive Flexibility (k=6, 4 studies, g=0.33, SE=0.11, 95% CI [0.13, 0.54], p=0.002), Processing speed (k=7, 4 studies, g=-0.85, SE=0.43, 95% CI [-1.70, -0.0002], p=0.050, Attention (k=12, 5 studies, g=0.25, SE=0.24, 95% CI [-0.22, 0.72], p=0.30), Executive Control (k=14, 6 studies, g=0.26, SE=0.16, 95% CI [-0.04, 0.56], p=0.0925), Time Processing (k=6, 2 studies, g=-0.12, SE=0.80, 95% CI [-1.69, 1.46], p=0.886), Working Memory (k=4, 4 studies, g=0.09, SE=0.12, 95% CI [-0.15, 0.33], p=0.467).

### Very Low-frequency vs High-frequency stimulation
### Verbal Fluency Subdomains

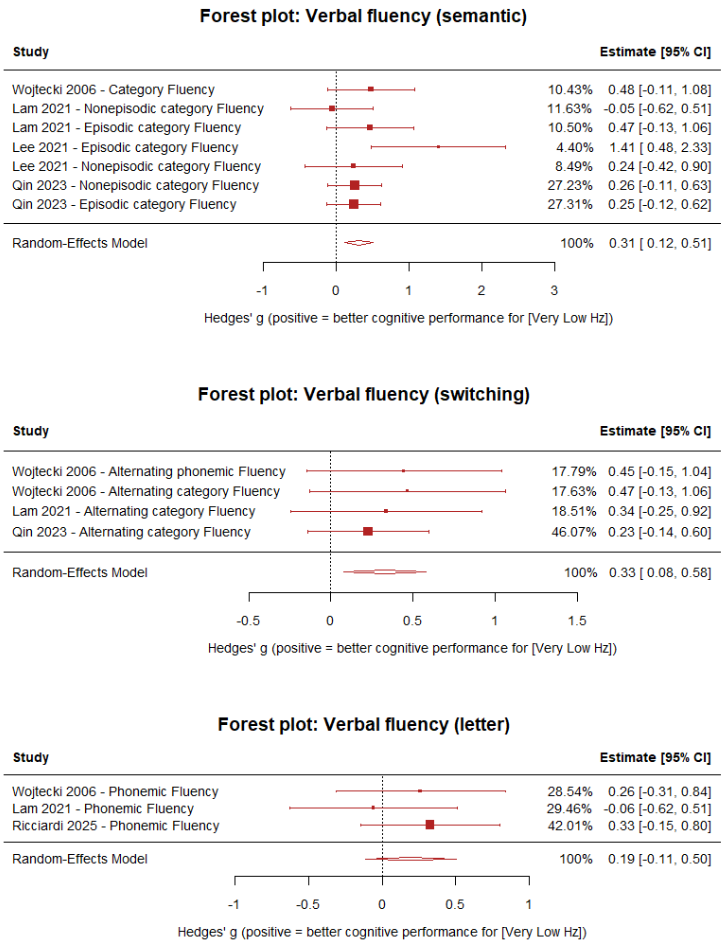


**Supplementary Figure 10. Effects of Very Low- vs High-Frequency STN-DBS on the main Verbal Fluency sub domains.** Multilevel meta-analysis adjusting for study-level dependence. Forest plot showing Hedges' g effect sizes for the verbal fluency subdomains comparing Very Low-frequency versus High-frequency STN-DBS across studies, estimated using a multilevel random-effects meta-analysis that accounts for dependency among multiple measures within the same study (effects nested within study). Positive values indicate better performance under Very Low-frequency stimulation relative to High-frequency stimulation. Across verbal fluency subdomains Very Low-frequency consistently outperformed High-frequency.

Semantic VF, k=7, g=0.31, SE=0.10, 95% CI [0.12, 0.51], p=0.001, Switching VF, k=4, g=0.32, SE=0.13, 95% CI [0.08, 0.58], p=0.0101). Letter VF, k=3, g=0.20, SE=0.16, 95% CI [-0.11, 0.50], p=0.214.

### Very Low-frequency vs High-frequency stimulation Executive Control Subdomains

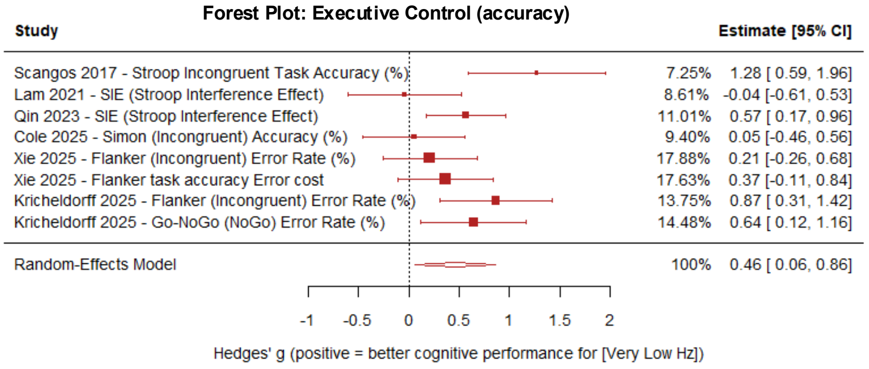


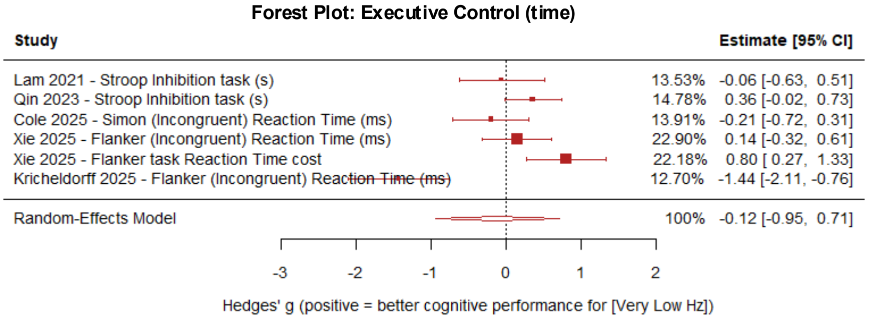


**Supplementary Figure 11. Effects of Low+Very Low- vs High-Frequency STN-DBS on the main Executive Control sub domains.** Multilevel meta-analysis adjusting for study-level dependence. Forest plot showing Hedges' g effect sizes for the Executive Control subdomains (accuracy and time metrics) comparing Low+Very Low-frequency versus High-frequency STN-DBS across studies, estimated using a multilevel random-effects meta-analysis that accounts for dependency among multiple cognitive domains measurements within the same study (effects nested within study). Positive values indicate better cognitive performance under Low+Very Low-frequency stimulation relative to High-frequency stimulation. Very low-frequency stimulation was associated with modest but statistically significant improvements in accuracy-based measures of executive control compared with High-frequency, whereas no significant differences were observed in time-based measures (Accuracy, k=8, 6 studies, g=0.46, SE=0.17, 95% CI [0.06, 0.86], p=0.030 and Time, k=6, 5 studies, g=-0.12, SE=0.32, 95% CI [-0.95, 0.71], p=0.722).

### Very Low-frequency vs High-frequency stimulation Main + Secondary + Tertiary Cognitive Domains

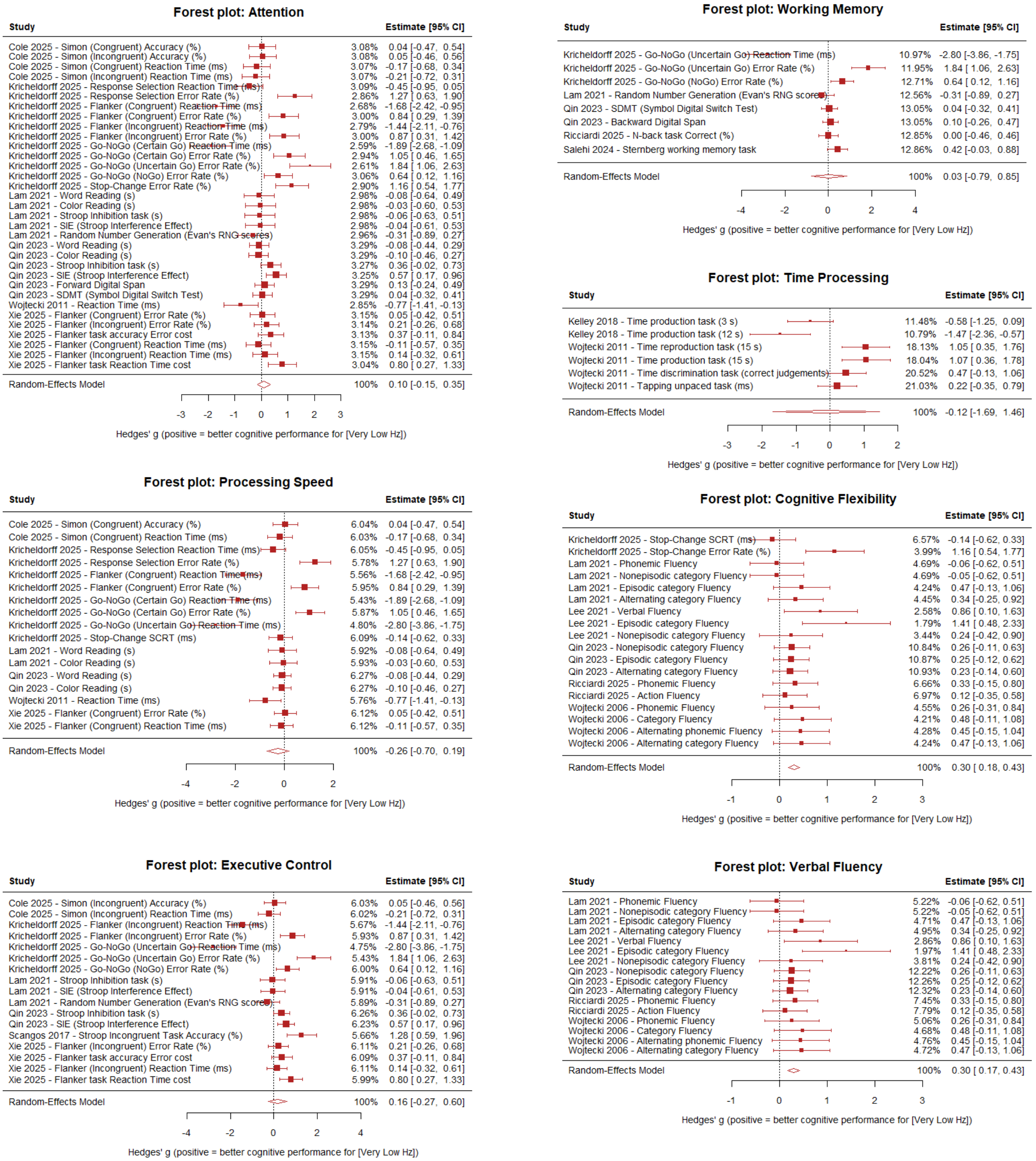


**Supplementary Figure 12. Effects of Very Low- vs High-Frequency STN-DBS on the overall cognitive domains regardless of aggregation/classification level (Main + Secondary + Tertiary).** Multilevel meta-analysis adjusting for study-level dependence. Forest plot showing Hedges' g effect sizes for the overall aggregated cognitive domains comparing Very Low-frequency versus High-frequency STN-DBS across studies, estimated using a multilevel random-effects meta-analysis that accounts for dependency among multiple cognitive domains measurements within the same study (effects nested within study). Positive values indicate better cognitive performance under Very Low-frequency stimulation relative to High-frequency stimulation. Very low-frequency stimulation was associated with modest but statistically significant improvements in verbal fluency and cognitive flexibility compared with High-frequency stimulation (Cognitive Flexibility, g=0.30, SE=0.06, 95% CI [0.18, 0.43], $p=1.4\times10^{-6}$, Verbal Fluency g=0.30, SE=0.07, 95% CI [0.17, 0.43], $p=6.7\times10^{-6}$). No significant differences were observed across the remaining cognitive domains: Attention (p=0.424), Executive Control (p=0.465), Processing Speed (p=0.265), Time Processing (p=0.886), and Working Memory (p=0.945).

#### Comparison of OFF stimulation with High-frequency stimulation

### OFF stimulation vs High-frequency stimulation Main Domains

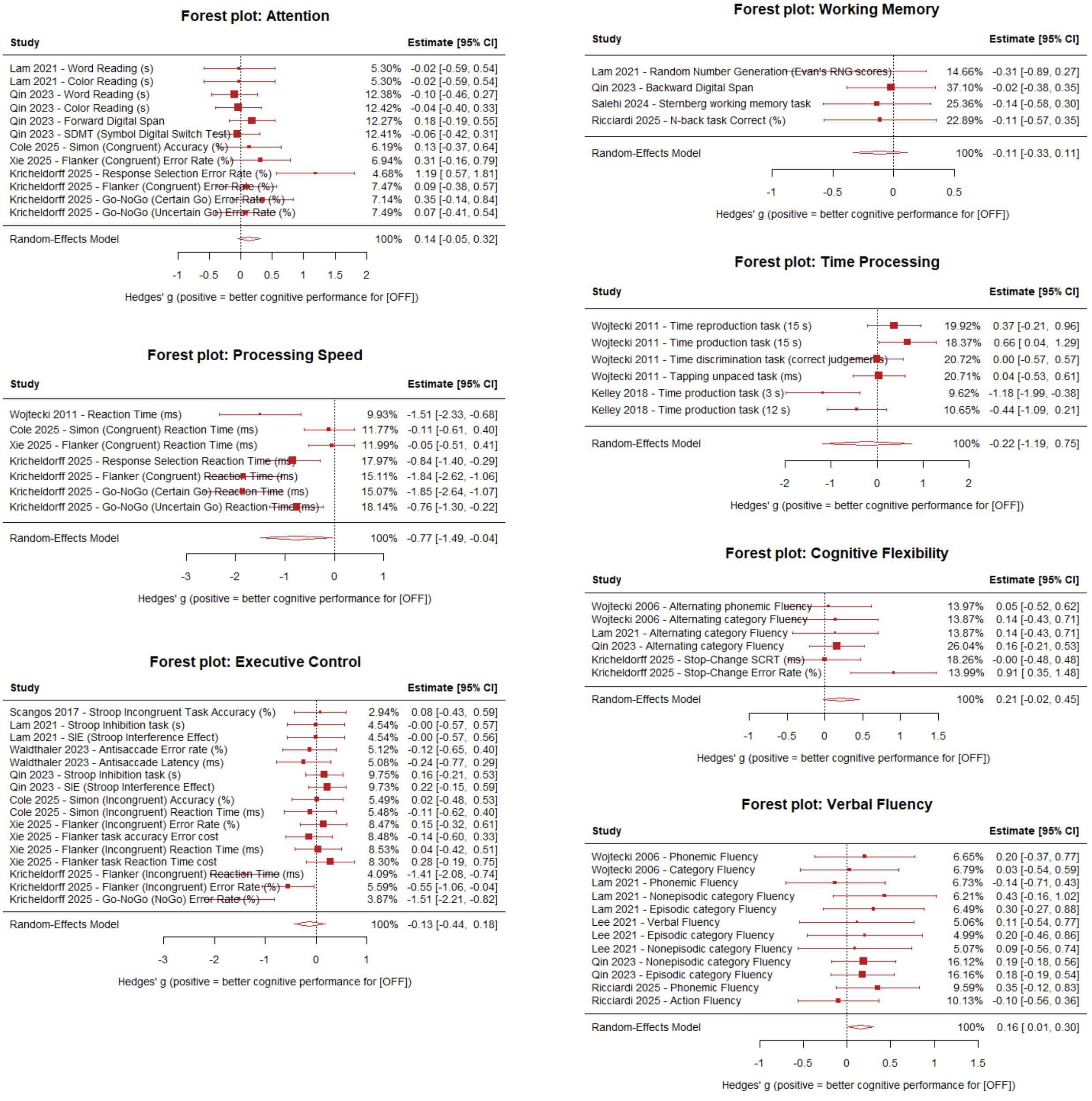


**Supplementary Figure 13. Effects of OFF stimulation vs High-Frequency STN-DBS on the main cognitive domains.** Multilevel meta-analysis adjusting for study-level dependence. Forest plot showing Hedges' g effect sizes for the main cognitive domains comparing OFF vs High-frequency stimulation STN-DBS across studies, estimated using a multilevel random-effects meta-analysis that accounts for dependency among multiple cognitive domains measurements within the same study (effects nested within study). Positive values indicate better cognitive domain performance in OFF relative to High-frequency stimulation.

Verbal Fluency (k=12, 5 studies, g=0.16, SE=0.08, 95% CI [0.01, 0.30], p=0.0385), Cognitive Flexibility (k=6, 4 studies, g=0.22, SE=0.12, 95% CI [-0.02, 0.45], p=0.075), Processing Speed (k=7, 4 studies, g=-0.77, SE=0.37, 95% CI [-1.49, -0.04], p=0.038), Attention (k=12, 5 studies, g=0.14, SE=0.09, 95% CI [-0.05, 0.32], p=0.142), Executive Control (k=16, 7 studies, g=-0.13, SE=0.16, 95% CI [-0.44, 0.18], p=0.424), Time Processing (k=6, 2 studies, g=-0.22, SE=0.49, 95% CI [-1.19, 0.75], p=0.658), and Working Memory (k=4, 4 studies, g=-0.11, SE=0.11, 95% CI [-0.33, 0.11], p=0.324).

### OFF stimulation vs High-frequency stimulation
### Verbal Fluency Subdomains

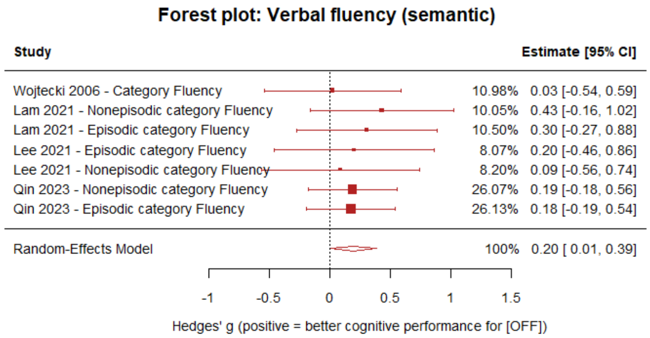


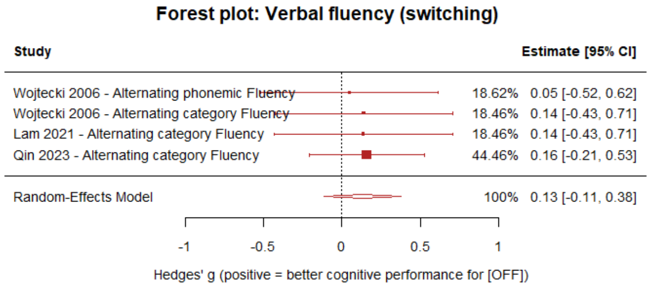


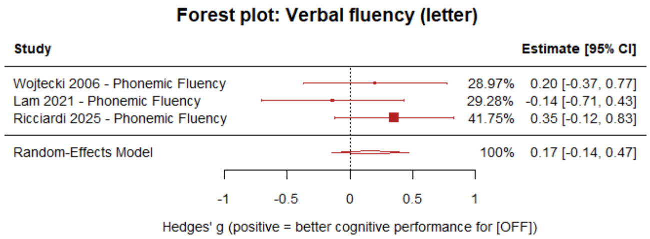


**Supplementary Figure 14. Effects of OFF stimulation vs High-Frequency STN-DBS on the main Verbal Fluency sub domains.** Multilevel meta-analysis adjusting for study-level dependence. Forest plot showing Hedges' g effect sizes for the verbal fluency subdomains comparing OFF vs High-frequency stimulation STN-DBS across studies, estimated using a multilevel random-effects meta-analysis that accounts for dependency among multiple measures within the same study (effects nested within study). Positive values indicate better performance in the OFF state relative to the High-frequency stimulation.

Semantic, k=7, g=0.20, SE=0.10, 95% CI [0.01, 0.39], p=0.0388; Letter, k=3, g=0.17, SE=0.16, 95% CI [-0.14, 0.47], p=0.293; Switching, k=4, g=0.13, SE=0.13, 95% CI [-0.11, 0.38], p=0.289

### OFF stimulation vs High-frequency stimulation
### Main + Secondary + Tertiary Cognitive Domains

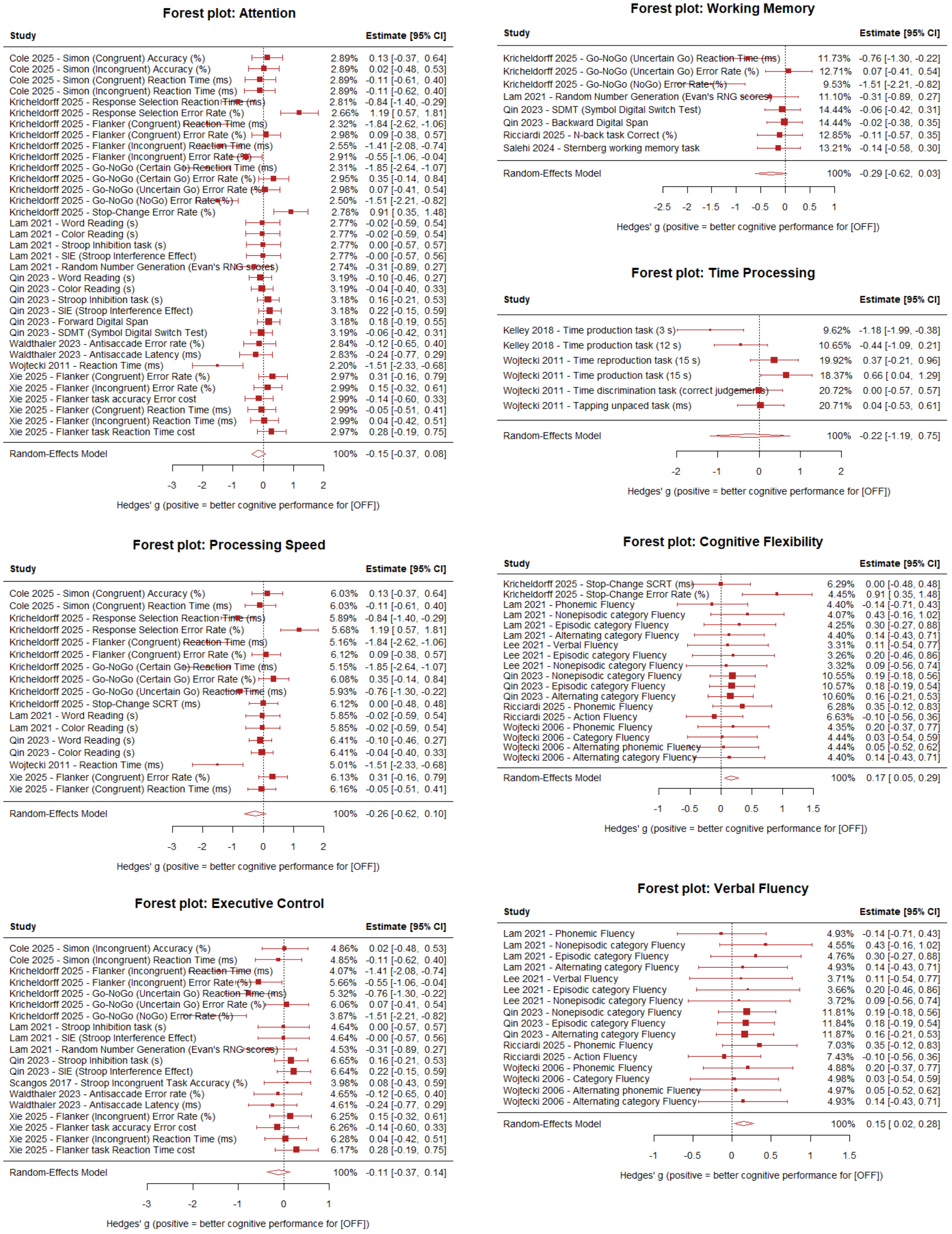


**Supplementary Figure 15. Effects of OFF stimulation vs High-Frequency STN-DBS on the overall cognitive domains regardless of aggregation/classification level (Main + Secondary + Tertiary).** Multilevel meta-analysis adjusting for study-level dependence. Forest plot showing Hedges' g effect sizes for the overall aggregated cognitive domains comparing the OFF vs High-frequency stimulation state across studies, estimated using a multilevel random-effects meta-analysis that accounts for dependency among multiple cognitive domains measurements within the same study (effects nested within study). Positive values indicate better cognitive performance in the OFF state relative to the High-frequency stimulation.

Verbal Fluency (g=0.15, SE=0.06, 95% CI [0.02, 0.28], p=0.020), Cognitive Flexibility (g=0.17, SE=0.06, 95% CI [0.05, 0.29], p=0.0043). Other domains showed non-significant effects: Attention (p=0.196), Executive Control (p=0.375), Processing Speed (p=0.152), Time Processing (p=0.658), and Working Memory (p=0.073).

#### Comparison of OFF stimulation with Low+Very Low-frequency stimulation

### OFF stimulation vs Low+Very Low-frequency stimulation
### Main Domains

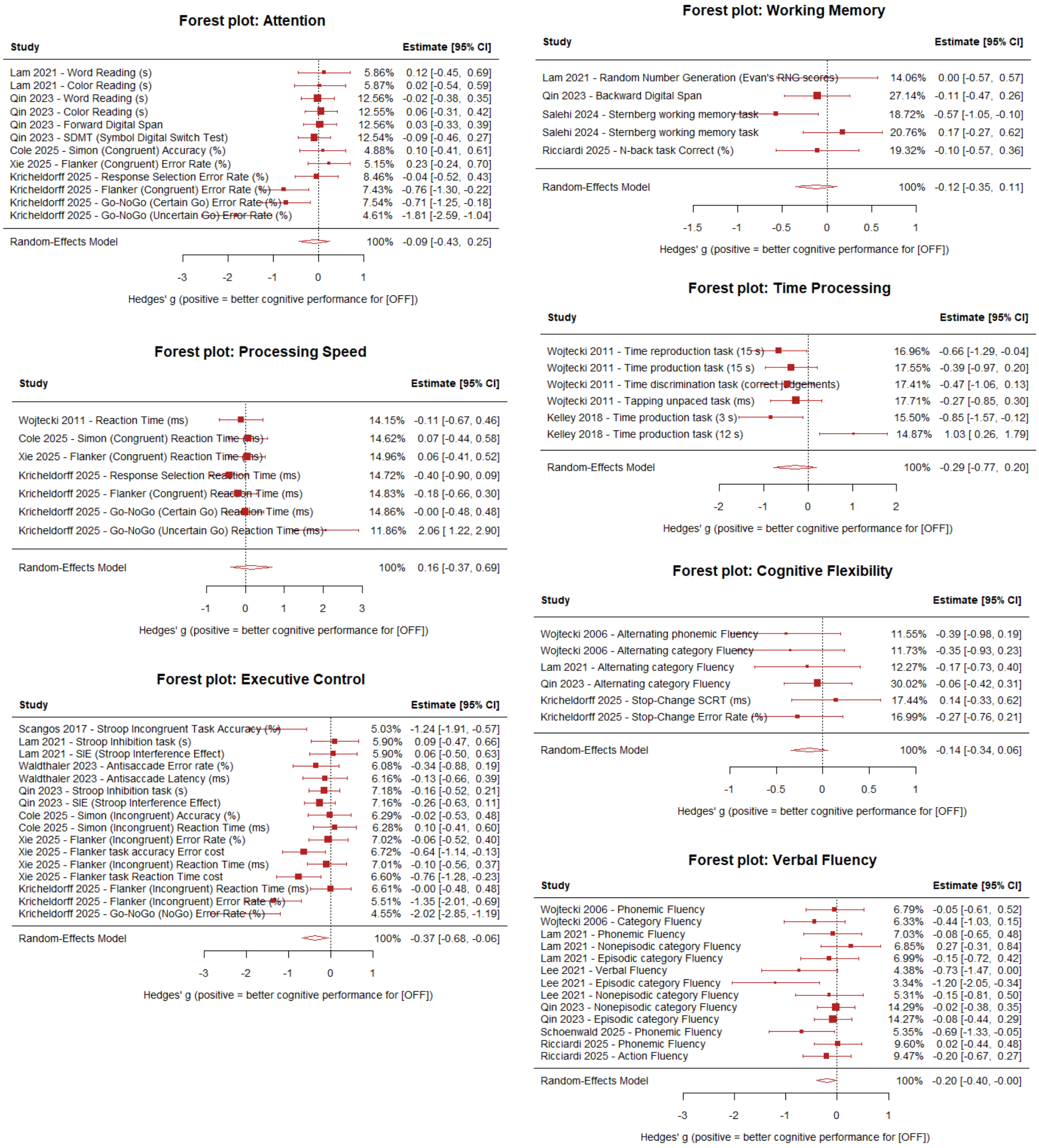


**Supplementary Figure 16. Effects of OFF stimulation vs Low+Very Low-frequency STN-DBS on the main cognitive domains.** Multilevel meta-analysis adjusting for study-level dependence. Forest plot showing Hedges' g effect sizes for the main cognitive domains comparing OFF vs LOW+VERY LOW Hz STN-DBS across studies, estimated using a multilevel random-effects meta-analysis that accounts for dependency among multiple cognitive domains measurements within the same study (effects nested within study). Negative values indicate better cognitive domain performance in LOW+VERY LOW Hz relative to OFF stimulation.

Verbal Fluency (k=13, 6 studies, g=-0.20, SE=0.10, 95% CI [-0.40, -0.005], p=0.0449), Executive Control (k=16, 7 studies, g=-0.37, SE=0.16, 95% CI [-0.68, -0.06], p=0.0181), Attention (k=12, 5 studies, g=-0.09, SE=0.17, 95% CI [-0.43, 0.25], p=0.593), Cognitive Flexibility (k=6, 4 studies, g=-0.14, SE=0.10, 95% CI [-0.34, 0.06], p=0.156), Processing Speed (k=7, 4 studies, g=0.16, SE=0.27, 95% CI [-0.37, 0.69], p=0.552, Time Processing (k=6, 2 studies, Hedges' g=-0.29, SE=0.25, 95% CI [-0.77, 0.20], p=0.243), Working Memory (k=5, 4 studies, g=-0.12, SE=0.12, 95% CI [-0.35, 0.11], p=0.311).

### OFF stimulation vs Low+Very Low-frequency stimulation
### Verbal Fluency Subdomains

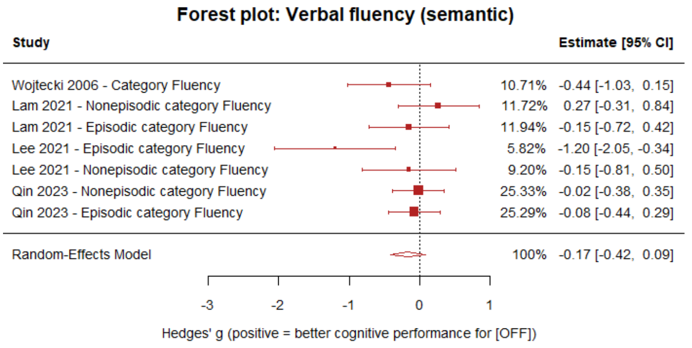


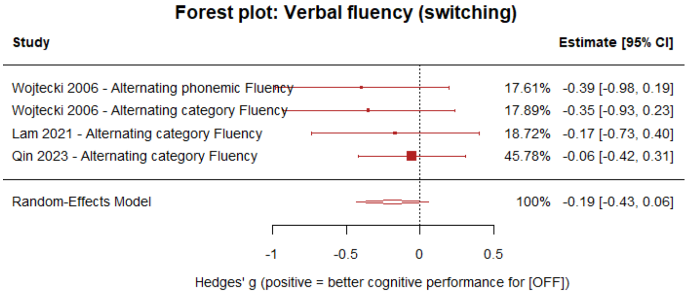


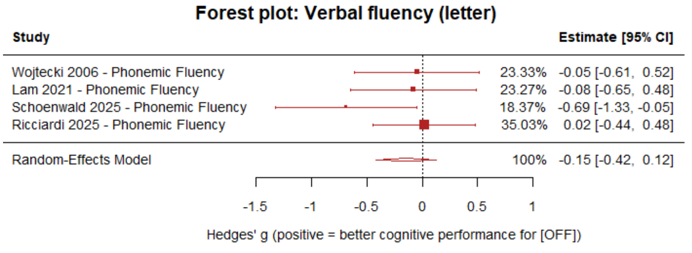


**Supplementary Figure 17. Effects of OFF stimulation vs Low+Very Low-frequency STN-DBS on the main Verbal Fluency sub domains.** Multilevel meta-analysis adjusting for study-level dependence. Forest plot showing Hedges' g effect sizes for the verbal fluency subdomains comparing OFF vs LOW+VERY LOW Hz STN-DBS across studies, estimated using a multilevel random-effects meta-analysis that accounts for dependency among multiple measures within the same study (effects nested within study). Negative values indicate better cognitive domain performance in LOW+VERY LOW Hz relative to OFF stimulation.

Semantic, k=7, g=-0.17, SE=0.13, 95% CI [-0.42, 0.09], p=0.198; Letter, k=4, g=-0.15, SE=0.14, 95% CI [-0.43, 0.12], p=0.279; Switching, k=4, g=-0.19, SE=0.13, 95% CI [-0.43, 0.06], p=0.136.

### OFF stimulation vs Low+Very Low-frequency stimulation Executive Control Subdomains

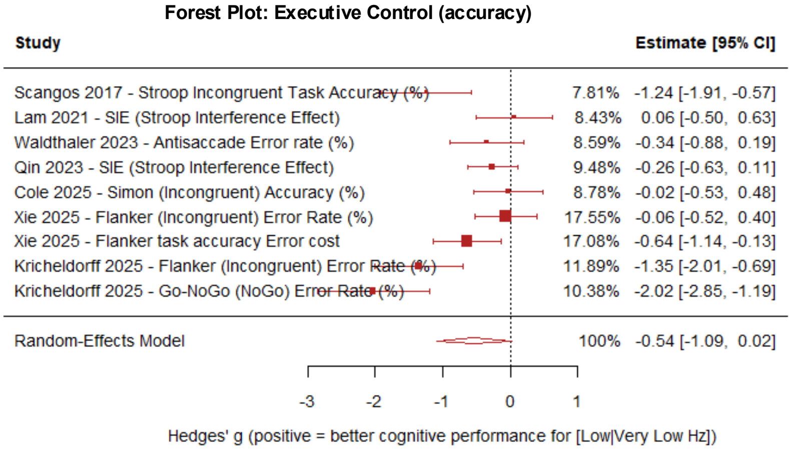


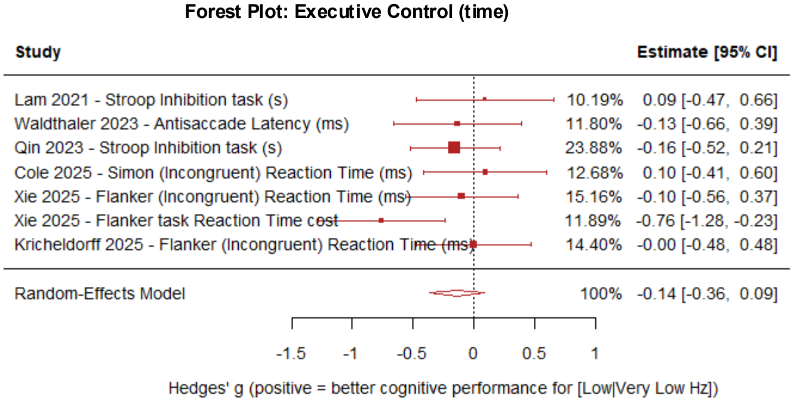


**Supplementary Figure 18. Effects of OFF stimulation vs Low+Very Low-Frequency STN-DBS on the main Executive Control sub domains.** Multilevel meta-analysis adjusting for study-level dependence. Forest plot showing Hedges' g effect sizes for the Executive Control subdomains (accuracy and time metrics) comparing OFF stimulation versus Low+Very Low-frequency STN-DBS across studies, estimated using a multilevel random-effects meta-analysis that accounts for dependency among multiple cognitive domains measurements within the same study (effects nested within study). Negative values indicate better cognitive performance under Low+Very Low-frequency stimulation relative to OFF stimulation. For executive control, subdomain analyses suggested the effect was driven by a better performance with Low+Very Low-frequency stimulation relative to OFF stimulation for accuracy-based measures (g=-0.54, p=0.057) and not time-based measures (g=-0.14, p=0.195)

### OFF stimulation vs Low+Very Low-frequency stimulation
### Main + Secondary + Tertiary Cognitive Domains

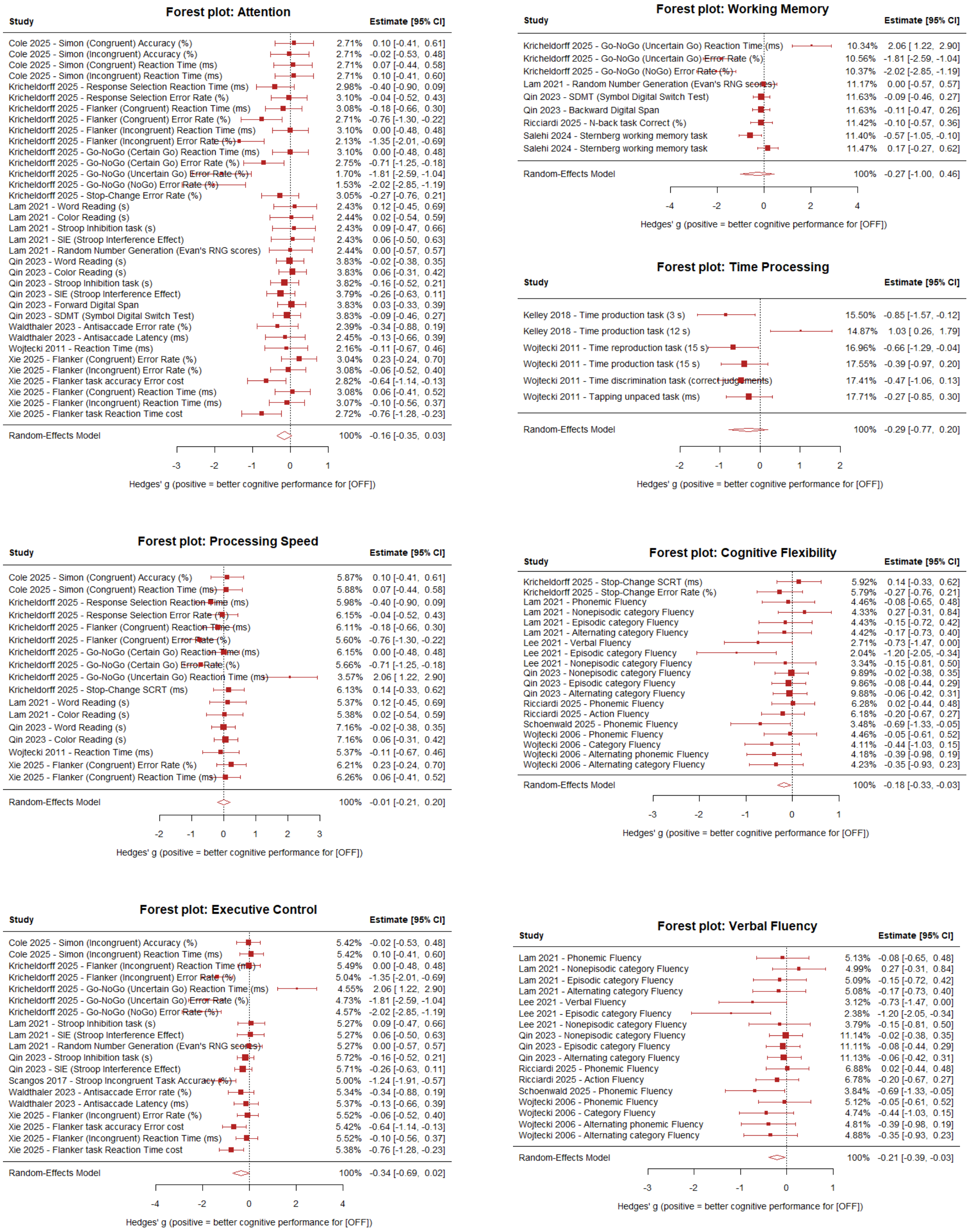


**Supplementary Figure 19. Effects of OFF stimulation vs Low+Very Low-frequency STN-DBS on the overall cognitive domains regardless of aggregation/classification level (Main + Secondary + Tertiary).** Multilevel meta-analysis adjusting for study-level dependence. Forest plot showing Hedges' g effect sizes for the overall aggregated cognitive domains comparing the OFF vs LOW+VERY LOW Hz state across studies, estimated using a multilevel random-effects meta-analysis that accounts for dependency among multiple cognitive domains measurements within the same study (effects nested within study). Negative values indicate better cognitive domain performance in LOW+VERY LOW Hz relative to OFF stimulation.

Verbal Fluency (Hedges' g=-0.21, SE=0.09, 95% CI [-0.40, -0.03], p=0.021), Cognitive Flexibility (g=-0.18, SE=0.07, 95% CI [-0.33, -0.03], p=0.016), Executive Control (g=-0.34, SE=0.18, 95% CI [-0.69, 0.02], p=0.064), Attention (g=-0.16, SE=0.10, 95% CI [-0.35, 0.03], p=0.091), Processing Speed (p=0.951), Time Processing (p=0.243), Working Memory (p=0.471).
